# State-level disparities in cervical cancer prevention and outcomes in the U.S.: A modeling study

**DOI:** 10.1101/2024.06.11.24308795

**Authors:** Fernando Alarid-Escudero, Valeria Gracia, Marina Wolf, Ran Zhao, Caleb W. Easterly, Jane J. Kim, Karen Canfell, Inge M.C.M. de Kok, Ruanne V. Barnabas, Shalini Kulasingam

## Abstract

**Background:** Despite HPV vaccines’ availability for over a decade, coverage across the US varies. While some states have tried to increase HPV vaccination coverage, most model-based analyses focus on national impacts. We evaluated hypothetical changes in HPV vaccination coverage at the national and state levels for California, New York, and Texas using a mathematical model.

**Methods:** We developed a new mathematical model of HPV transmission and cervical cancer, creating US and state-level models, incorporating country- and state-specific vaccination coverage and cervical cancer incidence and mortality. We quantified the national and state-level impact of increasing HPV vaccination coverage to 80% by 2025 or 2030 on cervical cancer outcomes and the time to elimination defined as <4 per 100k women.

**Results:** Increasing vaccination coverage to 80% in Texas over ten years could reduce cervical cancer incidence by 50.9% (95% credible interval [CrI]:46.6-56.1%) by 2100, from 1.58 (CrI:1.19-2.09) to 0.78 (CrI:0.57-1.02) per 100,000 women. Similarly, New York could see a 27.3% (CrI:23.9-31.5%) reduction, from 1.43 (CrI:0.93-2.07) to 1.04 (Crl:0.66-1.53) per 100,000 women, and California a 24.4% (CrI:20.0-30.0%) reduction, from 1.01 (Crl:0.66-1.44) to 0.76 (Crl:0.50-1.09) per 100,000 women. Achieving 80% coverage in five years will provide slightly larger and sooner reductions. If the vaccination coverage levels in 2019 continue, cervical cancer elimination could occur nationally by 2051 (Crl:2034-2064), but state timelines may vary by decades.

**Conclusion:** Targeting an HPV vaccination coverage of 80% by 2030 will disproportionately benefit states with low coverage and higher cervical cancer incidence. Geographically focused analyses can better inform priorities.

## Introduction

Rates of cervical cancer incidence and mortality have decreased dramatically since the introduction of Pap-smear-based screening in the U.S. (1) Persistent infection with high-risk human papillomavirus (HPV) is the cause of most cervical cancers (2); vaccines are now available to prevent infection with HPV. Further decreases in cancer incidence and mortality are predicted over the coming decades, assuming widespread coverage of HPV vaccines and higher screening coverage (3,4). In 2018, the American Cancer Society (ACS) stated a nationwide goal of eliminating cervical cancer as a public health problem. This statement was endorsed by all National Cancer Institute-funded cancer centers nationally. To achieve this goal, a target of 80% HPV vaccination coverage for boys and girls by 2025 was set. This target aligns with the Department of Health and Human Services’ Healthy People 2030 goals, stating that 80% of adolescents aged 13 to 15 years be vaccinated by 2030 (5–7).

HPV vaccines have been approved in the U.S. since 2006, when the FDA first approved them for girls. Later, in 2009, the approval was extended to boys. Despite the availability of these vaccines for over a decade, the coverage across the U.S. is varied.(8) Furthermore, areas with low vaccine coverage face an increased burden of screening, abnormal results, and cancer cases. This variation may potentially impact the long-term reductions in cervical cancer and cervical cancer deaths, limiting efforts to reach elimination (9) as predicted by modeling studies focused on the U.S. as a whole (4). According to the National Immunization Survey (NIS)-Teen, the coverage of HPV up-to-date vaccination varies greatly across states. Mississippi has a coverage of only 13%, while Rhode Island covers 62%.(10) This variation is significantly different from other vaccines for adolescents (11–13). Efforts to increase HPV vaccination coverage could be undermined by potential backslides in coverage, for example, during the COVID-19 pandemic. (14) While restrictions during the pandemic have been lifted, HPV vaccination continues to be adversely impacted.(15,16) The longer-term impact of potential future disruptions on cancer incidence and death remains to be quantified. The aim of this analysis is to predict the impact of increased, static, or decreased vaccination coverage in three U.S. states (Texas, California, and New York) using a mathematical model of HPV and cervical cancer tailored to state-level cancer incidence and mortality. The three states together account for nearly 27% of the US population (17), and differ on HPV vaccination and screening coverage, cancer incidence, cancer mortality, and demographic characteristics (age, race/ethnicity) (17). We compare these results to those obtained for the U.S. as a whole to assess how geographically focused analyses could inform priorities.

## Methods

### Overview

We developed the CISNET CERVIX Collaborative (C3) model, a compartmental model of transmission of type-specific high-risk HPV in males and females, and cervical cancer carcinogenesis in females. The model incorporates state-specific demographic dynamics (i.e., background mortality), hysterectomy rates, and sexual behavior (i.e., age- and sex-specific sexual debut and mixing), accounting for historical population-level age-specific cervical cancer screening and HPV vaccination up until 2018 (Supplementary Material). We parameterized the model based on clinical and epidemiological data from published and publicly available pre-published studies. We calibrated the model to age-, sex-, and type-specific HPV prevalence prior to HPV vaccination and the proportion of cervical cancers caused by HPV vaccination types in the US. The calibrated model projected disease outcomes and uncertainty in these outcomes under various intervention scenarios from 2019 to 2100. Scenarios comprised varying vaccination coverage starting in 2019. We adapted C3 to the cervical cancer incidence and mortality in three states in the US, accounting for state-specific vaccination and screening coverage. We assumed that the natural history of cervical cancer was identical across states, and thus used parameters obtained from calibration to the national data.

### Model structure and assumptions

The C3 model is an age- and sex-structured system of ordinary differential equations (ODEs) model of HPV dynamic transmission and cervical cancer carcinogenesis. The model accounts for HPV acquisition, clearance, and progression through a susceptible-infected-recovered-vaccinated (SIRV) structure (see Supplementary Material) with demography and realistic sexual mixing patterns (18), enabling age- and sex-specific vaccination interventions to be considered. The HPV infection states and transitions between health states are stratified by HPV type into high-risk vaccine-prevented types and non-vaccine-prevented types, assuming the administration of the nonavalent (9v) HPV vaccine. Women are at risk of becoming infected with carcinogenic HPV types (16, 18, 31, 33, 45, 52, and 58) currently targeted by the 9v HPV vaccine or other high-risk HPV (non-vaccine type) types (see Supplementary Material). The model uses a histologic classification to represent the pre-cancer and cancer health states. That is, women can clear their infections, stay infected, or progress to cervical intraepithelial neoplasia grade 2 (CIN-2), a pre-cancerous lesion, which can regress, persist, or progress to CIN-3 or cancer. Women with cancer can be detected by stage-specific symptoms and are at risk of cancer death (see Supplementary Material). Women are also at risk of age-specific mortality from other non-cervical cancer causes and of undergoing hysterectomy for benign conditions, after which women are no longer at risk for cervical cancer. The model also allows for screening to detect and treat CIN or detect preclinical cancer. Forward projections with the C3 model can compare future scenarios and consider various outcomes (e.g., HPV infections, cancer cases, and deaths). The model was implemented in the R programming language.(19) A detailed description of the C3 model is included in the Supplementary Material.

### Data and Model Inputs

To inform the U.S. and state-specific models, we used aggregated national and state-specific demographic data, such as life tables and population size, (20) sexual behavior data, (21) epidemiological data on HPV prevalence and cervical cancer incidence without vaccination or screening, (22–24) and HPV vaccination (25) and screening coverage. (26)

### Scenarios

We used C3 to evaluate a range of vaccination policies under a realistic screening scenario, where we increased or decreased the proportion of the HPV-vaccinated population relative to coverage levels in 2019. Screening was assumed to be cytology-based, starting at age 21 and ending at age 65, with different women attending at different frequencies from every year to every 5 years or never being screened. Based on reported trends, we modeled historical vaccination coverage from 2006 and 2009 for females and males, respectively, until 2019.(25) Our base-case scenario assumes that the 2019 screening and vaccination levels will continue over time. The main vaccination coverage scenarios assumed reaching 80% coverage in adolescents and varied the time this proportion was reached linearly at either 5 or 10 years starting in 2019. We assumed these vaccination scenarios would be implemented in 2019 and carried out until 2100 (i.e., the implementation period). Given the age of vaccination and the decades-long natural history process from HPV infection to potential cancer onset, a long period is required to observe any differences in outcomes between vaccination scenarios. We also accounted for a potential reduction in HPV vaccination coverage (i.e., a backslide scenario) for each state. Specifically, we decreased the proportion of vaccinated adolescents by 25% relative to the levels observed in 2019 and assumed this backslide lasts 5 years.

We used C3 to model the impact of disparities in HPV vaccination and screening coverage among three states - California, New York, and Texas, on cervical cancer incidence and mortality and compared them to the outcomes in the US as a whole (**Table 1**). We conducted sensitivity analyses on potential future screening coverage trends, modeling three different scenarios starting in 2019: 1) an increase in cytology-based screening, with 50% of never screeners starting screening every 5 years, 2) an increase in HPV-based screening, with 50% of never screeners starting screening with HPV testing every 5 years, and 3) a decrease in cytology-based screening, with 50% of women who screen every 5 years becoming never screeners.

**Table 1.** Country and state-specific vaccination and screening coverage. Vaccination coverage is defined as the proportion of boys or girls aged 13-17 who have received at least two doses of the HPV vaccine. Screening coverage is defined as the proportion of women aged 21-65 who have had a Pap test in the past three years. The denominator has been adjusted for women who had hysterectomy.

| Country<br>or State | Vaccination coverage in 2019 (25) |  | Screening coverage in 2020 (26) |
| --- | --- | --- | --- |
|  | ≥ Two dose of<br>HPV vaccine, boys<br>13-17 | ≥ Two dose of HPV<br>vaccine, girls 13-17 | Women aged 21-65 who have had a Pap<br>test in the past three years |
| U.S. | 54.8% | 60.7% | 77.7% |
| California | 54.5% | 70.9% | 79.3% |
| New York | 57.1% | 64.0% | 79.8% |
| Texas | 49.6% | 54.20% | 75.0% |

### Outcomes

We assessed the impact of the different vaccination scenarios on cervical cancer incidence, mortality and vaccinations per 100,000 eligible population compared to the base-case scenario, assuming 2019 coverage levels remained unchanged over the analysis period. We generated overall and age-specific outcome measures over time and percentage differences in these outcomes between the scenarios and policies relative to 2019 coverage. Additionally, we used hexamaps to show the age-period-cohort effects over time. Hexamaps visualize longitudinal population survey data by distinguishing age, calendar year (period), and birth cohort along three axes. (27)

### Model Calibration, Uncertainty Analysis, and Validation

We used the incremental mixture importance sampling (IMIS) algorithm (4), a Bayesian method, to calibrate 35 model parameters that could not be directly estimated from data. The IMIS algorithm has previously been used to calibrate cancer health policy models.(24,28) The parameters concerned HPV transmission and cervical cancer natural history, such as progression and regression rates (Supplementary Material). To generate projections for all primary outcomes for all scenarios, we sampled 1,000 parameter sets from the posterior distribution of the calibrated parameters and screening-related parameters, such as frequency of screening and sensitivity and specificity of screening. We did not account for uncertainty in sexual behavior and mixing. For each outcome, we estimated the posterior model-predicted mean and the 95% credible intervals (CrI) from the 2.5th and 97.5th percentiles of the projected values.

We internally validated the calibrated model to calibration targets.(29) We externally validated the vaccination component of the model by simulating various vaccination strategies and predicting the population-level impact, following Brisson et al. (30). We also externally validated each state-specific model by comparing model-predicted outcomes to age-specific incidence and mortality from the Surveillance, Epidemiology, and End Results (SEER) data from 2003-2013 (31), with the screening coverage observed during the same period (see the Supplementary Material for more details).

## Results

If HPV vaccination coverage remains at the same level as in 2019, cervical cancer incidence and mortality are expected to continue declining, reaching elimination with only four cases per 100,000 people by 2051 (Crl: 2034-2064) in the U.S. Individual states are predicted to reach elimination as well, with California reaching the elimination threshold by 2049 (Crl: 2033-2061), Texas by 2052 (Crl: 2043-2061), and New York by 2057 (Crl: 2038-2071) (**Figure 1**). Cancer mortality also continues to decrease. By 2100, cancer mortality rates are predicted to be 0.44 (Crl: 0.26-0.65) per 100,000 in the U.S., 0.36 (Crl: 0.23-0.52) per 100,000 in CA, 0.49 (Crl: 0.35-0.68) per 100,000 in TX and 0.52 (Crl: 0.33-0.77) per 100,000 in NY.

**Figure 1.**
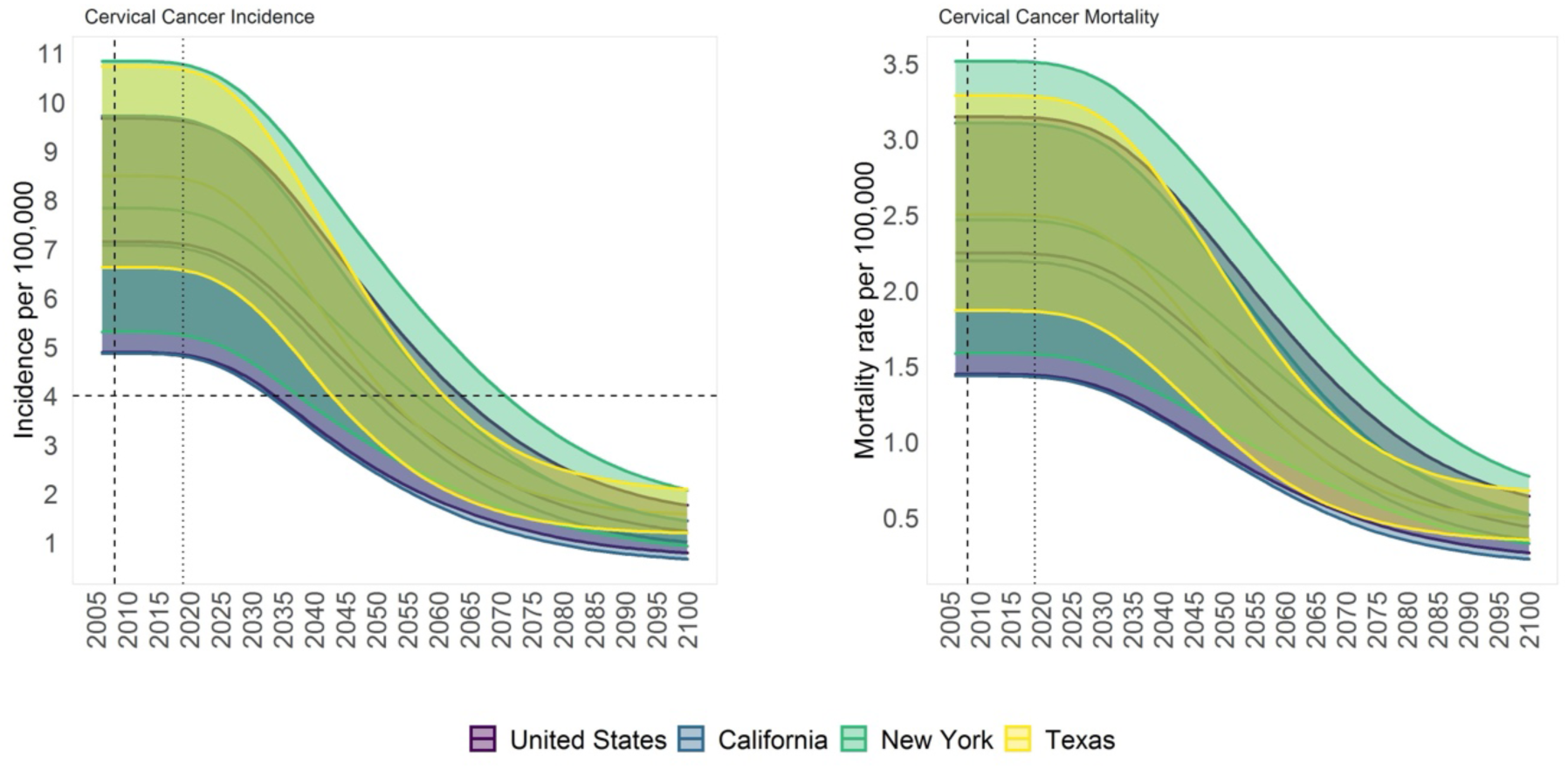
Cervical cancer incidence and mortality over time for the U.S., California, New York, and Texas, assuming HPV vaccination coverage in 2019. The vertical dashed line denotes the FDA approval of the HPV vaccine for women. The vertical dotted line denotes the beginning of the predicted outcomes. The horizontal dashed line in cervical cancer incidence at 4 per 100,00 represents the cervical cancer elimination goal. The shaded area shows the 95% posterior model-predictive interval (CrI) of the outcomes. The solid lines show the posterior model-predicted mean based on 1,000 simulations using samples from the posterior distribution.

A hypothetical backslide in vaccination coverage lasting 5 years compared to what it was in 2019 would increase cervical cancer incidence and mortality over the next 80 years. The impact will vary for each state and can be seen in **Figure 2** and **Figure 3**. Compared to if the 2019 coverage is continued at the national level, in the backslide scenario, cancer incidence in the US is likely to increase by 3.1% (CrI: 2.9-3.2%) in 2074 from 2.05 per 100,000 women (CrI: 1.26-2.97) to 2.11 (CrI: 1.30-3.06), and mortality is likely to increase by 3.0% (CrI: 2.9-3.2%) in 2082 from 0.64 per 100,000 women (CrI: 0.38-0.94) to 0.66 (CrI: 0.39-0.97). The increase is expected to remain at 1.8% (CrI: 1.6-2.1%) in incidence and 2.3% (CrI: 2.0-2.5%) in mortality by 2100 from 1.23 per 100,000 women (CrI: 0.79-1.76) to 1.26 (CrI: 0.80-1.80), and from 0.44 per 100,000 women (CrI: 0.27-0.64) to 0.45 (CrI: 0.27-0.66), respectively. In California, the impact would increase cervical cancer incidence by 3.4% (CrI: 3.2-3.6%) in 2076 from 1.66 per 100,000 women (CrI: 1.06-2.41) to 1.72 (CrI: 1.10-2.49) and mortality by 3.4% (CrI: 3.2-3.6%) in 2083 from 0.51 per 100,000 women (CrI: X0.32-0.76) to 0.53 (CrI: 0.33-0.78) relative to continuing with the 2019 coverage. The impact is expected to increase by 2.1% (CrI: 1.8-2.4%) in incidence from 1.01 per 100,000 women (CrI: 0.66-1.45) to 1.03 (CrI: 0.68-1.48) and 2.5% (CrI: 2.2-2.8%) in mortality from 0.35 per 100,000 women (CrI: 0.22-0.52) to 0.36 (CrI: 0.23-0.53) by 2100. Texas is likely to face the highest increase of 4.1% (CrI: 3.7-4.4%) in incidence in 2063 from 2.75 per 100,000 women (CrI: 1.97-3.74) to 2.86 (CrI: 2.05-3.88) and 3.9% (CrI: 3.7-4.1%) in mortality in 2070 from 0.79 per 100,000 women (CrI: 0.54-1.10) to 0.82 (CrI: 0.56-1.45) relative to continuing with the 2019 coverage. New York is likely to face the longest-lasting effect. It is expected to reach an increase of 2.8% (CrI: 2.6-2.9%) in incidence in 2080 from 2.14 per 100,000 women (CrI: 1.34-3.12) to 2.20 (CrI: 1.38-3.21) and 2.8% (CrI: 2.6-2.9%) in mortality in 2088 from 0.68 per 100,000 women (CrI: 0.41-1.00) to 0.70 (CrI: 0.43-1.03) relative to continuing with the 2019 coverage. The expected impact will likely remain at 2.5% (CrI: 2.3-2.7%) in both incidence and mortality by 2100 from 1.43 per 100,000 women (CrI: 0.93-2.07) to 1.47 (CrI: 0.95-2.12), and from 0.52 per 100,000 women (CrI: 0.33-0.77) to 0.54 (CrI: 0.34-0.79), respectively.

**Figure 2.**
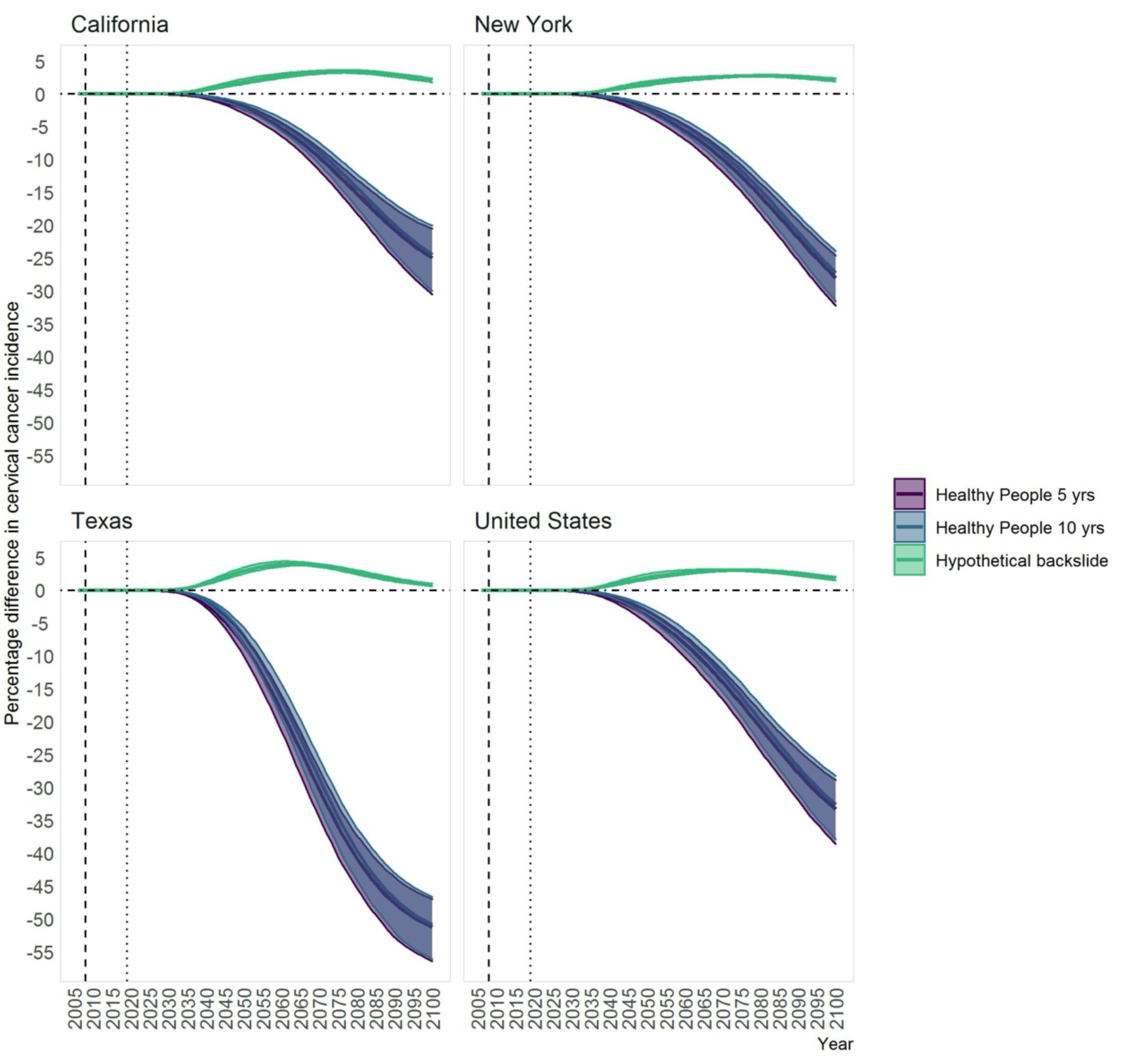
Percentage difference in cervical cancer incidence over time for different HPV vaccination coverage scenarios (i.e. 25% reduction in vaccination coverage for 5 years (’backslide’) or reaching 80% vaccination coverage (’healthy people’) in 5 or 10 years) relative to continuing with coverage in 2019 for the U.S. and three states. The shaded area shows the 95% posterior model-predictive credible interval (CrI) of the outcomes. The solid lines show the posterior model-predicted mean based on 1,000 simulations using samples from the posterior distribution.

**Figure 3.**
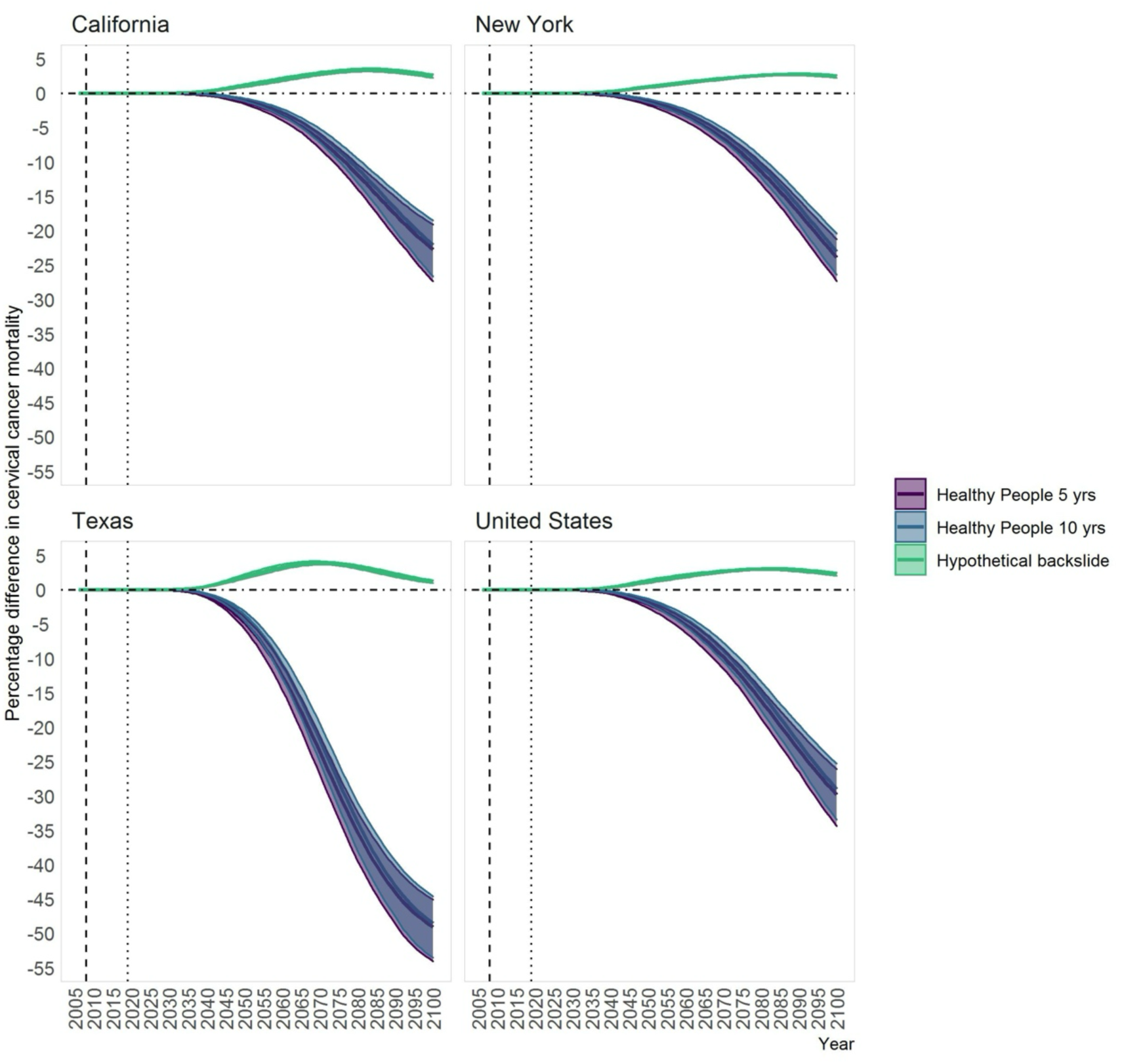
Percentage difference in cervical cancer mortality over time for different HPV vaccination coverage scenarios (i.e. 25% reduction in vaccination coverage for 5 years (’backslide’) or reaching 80% vaccination coverage (’healthy people’) in 5 or 10 years) relative to continuing with coverage in 2019 for the U.S. and three states. The shaded area shows the 95% posterior model-predictive credible interval (CrI) of the outcomes. The solid lines show the posterior model-predicted mean based on 1,000 simulations using samples from the posterior distribution.

Increasing HPV vaccination coverage to 80% following the Healthy People 2030 goals in five and 10 years would decrease the incidence and mortality of cervical cancer. The reduction will depend on the baseline vaccination coverage before increasing coverage. For instance, Texas, which had the lowest vaccination coverage among the three states in 2019, would see the largest decrease in cervical cancer incidence and mortality upon increasing the coverage. If 80% vaccination coverage is achieved within five years, the incidence of cervical cancer by 2100 could decrease by 51.2% (CrI: 47.0-56.5%) compared to 2019 levels from 1.58 (CrI: 1.19-2.09) to 0.77 (CrI: 0.57-1.01) per 100,000 women, requiring 9,844 additional vaccinations per 100,000 eligible individuals. If the same coverage is achieved within ten years, the decrease would be 50.9% (CrI: 46.6-56.1%) to 0.78 (CrI: 0.57-1.02) per 100,000 women (**Figure 2**), requiring 9,582 additional vaccinations per 100,000 eligible individuals. New York would reach a decrease of 28.0% (CrI: 24.6-32.3%) in cervical cancer incidence by 2100 from 1.43 (CrI: 0.93-2.07) to 1.03 (CrI: 0.65-1.51) per 100,000 women, with 80% coverage achieved in 5 years, and 27.3% (CrI: 23.9-31.5%) to 1.04 (Crl: 0.66-1.53) per 100,000 women with the same coverage achieved in 10 years, requiring 6,542 and 6,360 additional vaccinations per 100,000 eligible individuals, respectively. The smallest predicted effect would be in California, with decreases of 25.0% (CrI: 20.5-30.5%) and 24.4% (CrI: 20.0-30.0%) by 2100, from 1.01 (Crl: 0.66-1.44) to 0.76 (Crl: 0.49-1.09) and 0.76 (Crl: 0.50-1.09) per 100,000 women if 80% coverage is achieved in five and ten years, respectively. These predictions would require 5,792 and 5,630 additional vaccinations per 100,000 eligible individuals, respectively.

The hexamaps in **Figure 4** show the percentage differences in age-specific cervical cancer incidence by birth cohort and calendar year for each state, comparing different scenarios and policies to the coverage in 2019. This helps illustrate the potential impact of a hypothetical backslide and achieving an 80% vaccination coverage compared to 2019 levels. For instance, 80% vaccination coverage attained in 5 years will result in the lowest cancer incidence among cohorts born after 1994. Texas would benefit the most if 80% vaccination coverage is achieved within the next five to 10 years.

**Figure 4.**
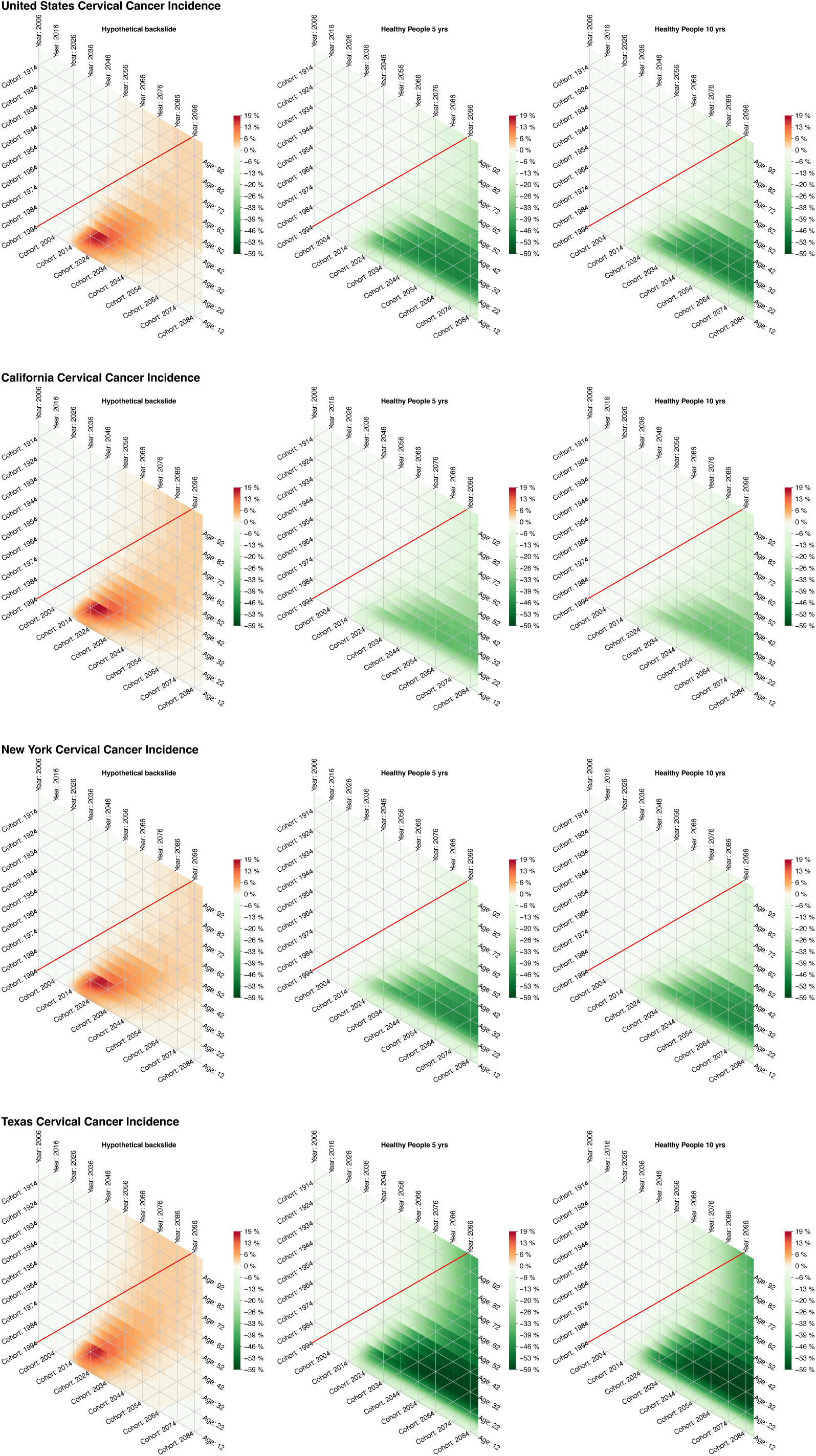
Hexamaps visualizing percentage differences in cervical cancer incidence for each scenario compared to continuing with 2019 coverage as a function of age, calendar year, and birth year. The vertical lines represent calendar years, the horizontal lines represent ages, and the diagonal lines represent the aging of each birth cohort with calendar years from the bottom left to the top right. In each scenario, the red diagonal line represents the birth cohort that became eligible to be vaccinated from the start of the simulation. Cohorts born after 1994 are represented by less heated colors at the bottom right of the red line.

The base-case results are influenced by potential changes in future screening trends. If screening rates increase in the future, with 50% of individuals who have never been screened starting to get screened every 5 years using either cytology-based or HPV testing, the time it takes to eliminate cervical cancer would decrease by 11 years for the entire US, 10 years for California, 14 years for New York, and 6 years for Texas. Furthermore, there would be a greater reduction in cervical cancer incidence and mortality if HPV vaccination coverage reaches 80% in five and 10 years, as per the Healthy People 2030 objectives, compared to the base-case screening coverage. Conversely, if screening rates decrease in the future, with 50% of women who currently screen every 5 years using cytology becoming never-screeners, the time to eliminate cervical cancer would increase by 6 years for the entire US, 4 years for California, 5 years for New York, and 7 years for Texas. However, a similar reduction in cervical cancer incidence and mortality would occur if HPV vaccination coverage reaches 80% in five and 10 years, as per the Healthy People 2030 objectives, compared to the base-case screening coverage.

## Discussion

We developed a new dynamic model of HPV transmission and cervical cancer carcinogenesis to assess different vaccination scenarios for the U.S. as a whole and compare them with three states that have variable vaccination and screening coverage levels. We also modeled a hypothetical backslide scenario of decreased vaccination coverage. If 2019 vaccination coverage continues, cervical cancer elimination would be reached in the US by 2051 (Crl: 2034-2064). However, the timeline by which individual states reach elimination could vary by decades. California, New York, and Texas could reach elimination by 2049 (Crl: 2033-2061), 2057 (Crl: 2038-2071), and 2052 (Crl: 2043-2061), respectively. We also found that the benefits of increased vaccination coverage to the Healthy People 2030 goals are heterogeneous across the different states. Our model predicts that even a slight decline in vaccination coverage over a short period could increase both cervical cancer incidence and mortality for the next 100 years. This will have a greater impact on states with currently lower vaccination coverage levels.

Our results align with a previous modeling study analyzing potential decreases in vaccination coverage due to the COVID-19 pandemic. For example, Daniels et al., (32) found that a decrease of 21-71% in the coverage of HPV vaccination in the U.S. in 2020 and 2021 could increase cervical cancer incidence over the next 100 years. The increase could be as high as 5% by 2060. Although our results align for the overall U.S., the relative impact would vary across states, from 2.8% in New York to 4.1% in Texas.

According to Saxena et al. (33), a 24% decline in HPV vaccination coverage in 2021 would take up to a decade to clear the accumulated deficit in HPV vaccination doses. Even though the evidence suggests that vaccination coverage for teenagers in the U.S. is returning to 2019 levels, it is uncertain how it varies by state and affects younger age groups eligible for the vaccine but not captured by NIS-TEEN.(11) Not only COVID-19 but also vaccine safety concerns have affected coverage.(34) It is possible that catch-up HPV vaccination may not reduce the impact of a one-time decrease in coverage if women in the catch-up program are older and have already been infected with HPV.

Our findings are consistent with previous research from a comparative model analysis showing that HPV vaccination can expedite cervical cancer elimination in the U.S. (4) However, NIS-Teen data show large differences in state-level HPV vaccination coverage, varying from 61% in Mississippi to 95% in Rhode Island. The difference becomes even greater in up-to-date regimens, from 38.5% in Mississippi to 85.2% in Rhode Island in 2022.(11) Moreover, there is significant variation in HPV vaccination coverage levels across states, and even within states, which is strongly associated with racial/ethnic minorities and poverty. Therefore, increasing vaccination rates, particularly in states with higher levels of poverty and/or larger historically disadvantaged racial/ethnic minority populations, has the potential to reduce health disparities and promote greater health equity.(9) Identifying which state populations are less likely to be vaccinated will help focus efforts on improving vaccine uptake, especially among marginalized groups.

Our study has several limitations. First, the screening strategy we modeled was based on a simplified cytology-based algorithm and did not include HPV testing. As a result, our model might not have fully considered the potential benefits of either HPV and cytology co-testing or primary HPV test-based screening, along with increased HPV vaccination in terms of reducing time to elimination. Second, we modeled the nonavalent HPV vaccine that protects against the seven most oncogenic HPV types, but not all vaccinations in the U.S. are with the nonavalent vaccine. Thus, the predicted outcomes of increasing HPV vaccination should be considered the maximum potential benefit of the scenarios, assuming the nonavalent vaccine is deployed. More detailed individual-level models, such as the CISNET CERVIX models, should be used to account for more detailed vaccination coverage and screening algorithms.(35,36) Finally, we did not consider international or between-state migration. Previous state-level modeling analysis by Durham et al. (21) showed that migration, which differs by state, could potentially weaken the effectiveness of vaccination programs at the state level. Therefore, it is essential to coordinate efforts across all states to achieve U.S.-level elimination goals. The implications for these varying migrations between states and into the US could be quantified by accounting for the age and gender of the migrant population, the prevalence of HPV (specifically types targeted by the nonavalent vaccine) based on country or region of origin, and assumptions regarding mixing by sex/gender and age with in-state residents. It may also be the case that HPV vaccination rates are higher for a particular migrant population than the rates in the state they migrate to and may differ based on whether migrants are documented or not (4). However, given the complexity of this topic, this is the subject of future research.

Our study has several strengths. First, we developed the C3 model, a dynamic model of HPV transmission and cervical cancer carcinogenesis that accounts for realistic age-specific sexual mixing patterns and the herd immunity effect of vaccination. Using a system of ODEs, C3 requires fewer computational resources than other more complex microsimulation models to simulate vaccination coverage scenarios and their effects on outcomes. C3 enables quantification and propagation of uncertainty on the natural history and external parameters to generate probabilistic projections— not only producing estimates of expected outcomes—to assess a range of possible outcomes, such as the time to cervical cancer elimination under different scenarios, in a parsimonious manner. We used comprehensive data on HPV prevalence, cervical cancer incidence and mortality, and screening and vaccination coverage to estimate the model’s parameters representing the transmission dynamics in the U.S. and each state. We also validated the C3 model at both the national and state-specific levels using a combination of natural history targets, vaccine efficacy, and screening and vaccine outcomes. This approach could be adapted to other states in the US and potentially other countries.

In summary, we used a new mathematical model to assess the impact of increasing HPV vaccination coverage to reach the Healthy People goal by either 2025 or 2030 on cervical cancer outcomes for the U.S. and three states with varying vaccination coverage levels. Our projections indicate that the findings for the U.S. do not apply to all states. Therefore, we recommend developing tailored models to identify and address local barriers to coverage. States with low current vaccination coverage face disproportionally worse outcomes if vaccination coverage declines, but would relatively benefit the most from increasing vaccination coverage as soon as possible. Increasing vaccination levels in states with low current coverage might reach similar benefits to uniformly increasing vaccination coverage across the entire country. State officials should prioritize efforts to promote HPV vaccination uptake to reduce the time needed to compensate for deficiencies in coverage due to historical and potential future disruptions in vaccination coverage.

## Data Availability Statement

Model outputs underpinning the analysis are available from the authors upon reasonable request.

## Acknowledgements

Previous versions of this work have been presented at prior annual and mid-year academic meetings of the Cancer Intervention and Surveillance Modeling Network (CISNET).

