## Supplementary Material for "State-level disparities in cervical cancer prevention and outcomes in the U.S.: A modeling study"

October 17, 2024

**6 Natural History Model Equations** **42**

### 1 Model Structure

The CISNET CERVIX Collaborative (C3) model is a compartmental dynamic transmission model of a heterosexual population stratified into 24 age groups and three sexual activity groups, implemented as a system of ordinary differential equations (ODEs) of type-specific HPV in males and females, and cervical cancer carcinogenesis in females, that incorporates U.S. and state-specific demographic dynamics and sexual behavior, accounting for historical population-level screening and vaccination and potential future interventions.

#### 1.1 Epidemiological structure

The structure of the model can be thought of as the combination of three simple models into one more complex model. The first is a **S**usceptible-**I**nfected-**R**ecovered (SIR) model [16], which represents the types of HPV for which vaccines are not available. The second is a **S**usceptible-**I**nfected-**R**ecovered-**V**accinated (SIRV) model accounting for breakthrough infections (**W**) [13], which represents the types of HPV for which a vaccine is available. The third is a model of cervical cancer carcinogenesis, describing the development of pre-cancerous cervical lesions from HPV infection and leading to cervical cancer.

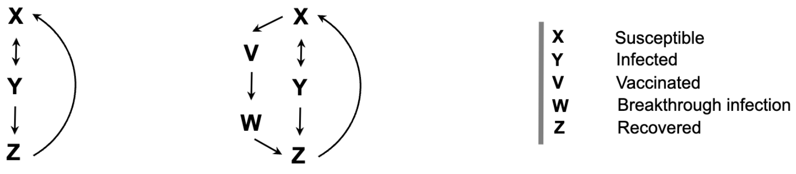

Supplementary Figure 1: Structure of SIR and SIRV models.

The combination of the first two models shown in Figure 1 results in a 15-epidemiological-state transmission model where individuals can simultaneously become infected and move through the epidemiological states for HPV caused by types for which a vaccine exists and for types for which a vaccine does not exist (Figure 2). In addition to being able to transition through any of the states, individuals who are infected with either type of HPV can develop cervical lesions and eventually cancer if the infection persists (Figure 3).

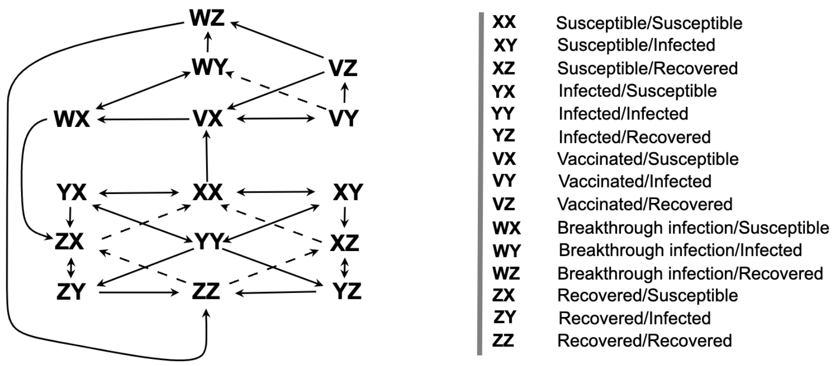

Supplementary Figure 2: Structure of the transmission model. The first letter in each state indicates the vaccine type of HPV infection status; the second letter indicates the non-vaccine type of HPV infection status, e.g., **YZ** is a state for individuals who are infected with a vaccine-type HPV and recovered from a non-vaccine-type HPV.

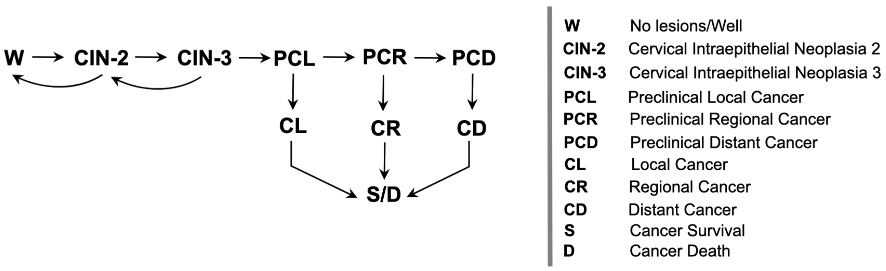

Supplementary Figure 3: Women infected with either type of HPV can develop cervical lesions, moving from the **No lesions/Well** state into **CIN-2** or **CIN-3** states, and eventually the cancer states while maintaining their infection status. Additionally, women over the age of 18 in the **No lesions/Well**, **CIN-2**, and **CIN-3** states will be at risk of getting a hysterectomy at a rate based on demographic data, after which women are no longer at risk for cervical cancer. All women are at risk of dying from all causes as a function of age.

The resulting structure from the combination of the above model subcomponents has 108 health states (see Table 1), which are also stratified into three sexual activity levels, twenty-four age groups, and some of them expanded to account for time-dependence in their transition rates [2]. All the model equations are shown in section 6.

Figure 4 shows the naming convention for most of the health states (i.e., model's variables).

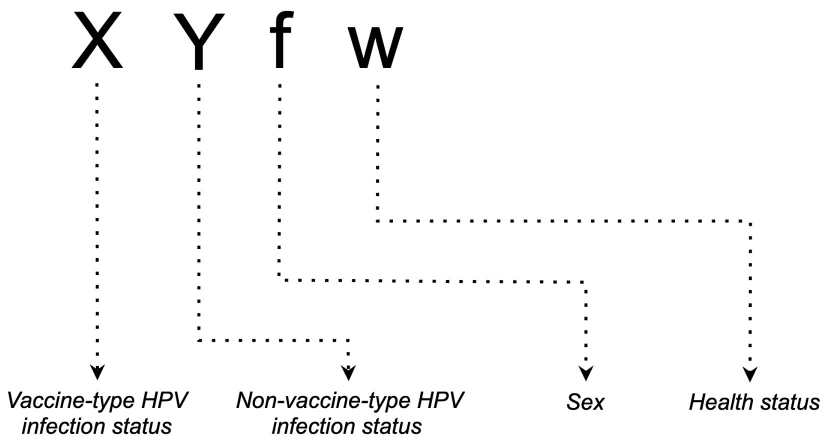

Supplementary Figure 4: State definitions showing how to interpret the letter abbreviations for health states of the model. The definitions are also listed in Table 1. Possible infection status are **X**-Susceptible, **Y**-Infected, **Z**-Recovered, **V**-Vaccinated, **W**-Breakthrough infection. Sex can either be **m**-Male or **f**-Female. The health status can either be **w**-No lesions/Well, **c2**-CIN-2, **c3**-CIN-3, **pcl**-Preclinical local cancer, **cl**-Local cancer, **pcr**-Preclinical regional cancer, **cr**-Regional cancer, **pcd**-Preclinical distant cancer, or **cd**-Distant cancer. Males can only have the health status **w**. There are exceptions to this scheme, which are noted in Table 1.

Supplementary Table 1: Model states

| State | Sex | Vaccine type HPV infection status | Non-vaccine type HPV infection status | Health Status |
|---|---|---|---|---|
| XXmw | Male | Susceptible | Susceptible | No lesions/Well |
| XXfw | Female | Susceptible | Susceptible | No lesions/Well |
| XYmw | Male | Susceptible | Infected | No lesions/Well |
| XYfw | Female | Susceptible | Infected | No lesions/Well |
| XYfc2 | Female | Susceptible | Infected | CIN-2 |
| XYfc3 | Female | Susceptible | Infected | CIN-3 |
| XYfpcl | Female | Susceptible | Infected | Preclinical local cancer |
| XYfcl | Female | Susceptible | Infected | Local cancer |
| XYfpcr | Female | Susceptible | Infected | Preclinical regional cancer |
| XYfcr | Female | Susceptible | Infected | Regional cancer |
| XYfpcd | Female | Susceptible | Infected | Preclinical distant cancer |

Model states (continued)

| State | Sex | Vaccine type HPV infection status | Non-vaccine type HPV infection status | Health Status |
|---|---|---|---|---|
| XYfcd | Female | Susceptible | Infected | Distant cancer |
| XZmw | Male | Susceptible | Recovered | No lesions/Well |
| XZfw | Female | Susceptible | Recovered | No lesions/Well |
| YXmw | Male | Infected | Susceptible | No lesions/Well |
| YXfw | Female | Infected | Susceptible | No lesions/Well |
| YXfc2 | Female | Infected | Susceptible | CIN-2 |
| YXfc3 | Female | Infected | Susceptible | CIN-3 |
| YXfpcl | Female | Infected | Susceptible | Preclinical local cancer |
| YXfcl | Female | Infected | Susceptible | Local cancer |
| YXfpcr | Female | Infected | Susceptible | Preclinical regional cancer |
| YXfcr | Female | Infected | Susceptible | Regional cancer |
| YXfpcd | Female | Infected | Susceptible | Preclinical distant cancer |
| YXfcd | Female | Infected | Susceptible | Distant cancer |
| YYmw | Male | Infected | Infected | No lesions/Well |
| YYfw | Female | Infected | Infected | No lesions/Well |
| YYfc2 | Female | Infected | Infected | CIN-2 |
| YYfc3 | Female | Infected | Infected | CIN-3 |
| YYfpcl | Female | Infected | Infected | Preclinical local cancer |
| YYfcl | Female | Infected | Infected | Local cancer |
| YYfpcr | Female | Infected | Infected | Preclinical regional cancer |
| YYfcr | Female | Infected | Infected | Regional cancer |
| YYfpcd | Female | Infected | Infected | Preclinical distant cancer |
| YYfcd | Female | Infected | Infected | Distant cancer |
| YZmw | Male | Infected | Recovered | No lesions/Well |
| YZfw | Female | Infected | Recovered | No lesions/Well |
| YZfc2 | Female | Infected | Recovered | CIN-2 |
| YZfc3 | Female | Infected | Recovered | CIN-3 |
| YZfpcl | Female | Infected | Recovered | Preclinical local cancer |
| YZfcl | Female | Infected | Recovered | Local cancer |
| YZfpcr | Female | Infected | Recovered | Preclinical regional cancer |
| YZfcr | Female | Infected | Recovered | Regional cancer |
| YZfpcd | Female | Infected | Recovered | Preclinical distant cancer |
| YZfcd | Female | Infected | Recovered | Distant cancer |
| VXmw | Male | Vaccinated | Susceptible | No lesions/Well |
| VXfw | Female | Vaccinated | Susceptible | No lesions/Well |
| VYmw | Male | Vaccinated | Infected | No lesions/Well |
| VYfw | Female | Vaccinated | Infected | No lesions/Well |

Model states (continued)

| State | Sex | Vaccine type HPV infection status | Non-vaccine type HPV infection status | Health Status |
|---|---|---|---|---|
| VYfc2 | Female | Vaccinated | Infected | CIN-2 |
| VYfc3 | Female | Vaccinated | Infected | CIN-3 |
| VYfpcl | Female | Vaccinated | Infected | Preclinical local cancer |
| VYfcl | Female | Vaccinated | Infected | Local cancer |
| VYfpcr | Female | Vaccinated | Infected | Preclinical regional cancer |
| VYfcr | Female | Vaccinated | Infected | Regional cancer |
| VYfpcd | Female | Vaccinated | Infected | Preclinical distant cancer |
| VYfcd | Female | Vaccinated | Infected | Distant cancer |
| VZmw | Male | Vaccinated | Recovered | No lesions/Well |
| VZfw | Female | Vaccinated | Recovered | No lesions/Well |
| WXmw | Male | Breakthrough infection | Susceptible | No lesions/Well |
| WXfw | Female | Breakthrough infection | Susceptible | No lesions/Well |
| WXfc2 | Female | Breakthrough infection | Susceptible | CIN-2 |
| WXfc3 | Female | Breakthrough infection | Susceptible | CIN-3 |
| WXfpcl | Female | Breakthrough infection | Susceptible | Preclinical local cancer |
| WXfcl | Female | Breakthrough infection | Susceptible | Local cancer |
| WXfpcr | Female | Breakthrough infection | Susceptible | Preclinical regional cancer |
| WXfcr | Female | Breakthrough infection | Susceptible | Regional cancer |
| WXfpcd | Female | Breakthrough infection | Susceptible | Preclinical distant cancer |
| WXfcd | Female | Breakthrough infection | Susceptible | Distant cancer |
| WYmw | Male | Breakthrough infection | Infected | No lesions/Well |
| WYfw | Female | Breakthrough infection | Infected | No lesions/Well |
| WYfc2 | Female | Breakthrough infection | Infected | CIN-2 |
| WYfc3 | Female | Breakthrough infection | Infected | CIN-3 |
| WYfpcl | Female | Breakthrough infection | Infected | Preclinical local cancer |
| WYfcl | Female | Breakthrough infection | Infected | Local cancer |
| WYfpcr | Female | Breakthrough infection | Infected | Preclinical regional cancer |
| WYfcr | Female | Breakthrough infection | Infected | Regional cancer |
| WYfpcd | Female | Breakthrough infection | Infected | Preclinical distant cancer |
| WYfcd | Female | Breakthrough infection | Infected | Distant cancer |
| WZmw | Male | Breakthrough infection | Recovered | No lesions/Well |
| WZfw | Female | Breakthrough infection | Recovered | No lesions/Well |
| WZfc2 | Female | Breakthrough infection | Recovered | CIN-2 |
| WZfc3 | Female | Breakthrough infection | Recovered | CIN-3 |
| WZfpcl | Female | Breakthrough infection | Recovered | Preclinical local cancer |
| WZfcl | Female | Breakthrough infection | Recovered | Local cancer |
| WZfpcr | Female | breakthrough infection | Recovered | Preclinical regional cancer |

Model states (continued)

| State | Sex | Vaccine type HPV infection status | Non-vaccine type HPV infection status | Health Status |
|---|---|---|---|---|
| WZfcr | Female | breakthrough infection | Recovered | Regional cancer |
| WZfpcd | Female | breakthrough infection | Recovered | Preclinical distant cancer |
| WZfcd | Female | breakthrough infection | Recovered | Distant cancer |
| ZXmw | Male | Recovered | Susceptible | No lesions/Well |
| ZXfw | Female | Recovered | Susceptible | No lesions/Well |
| ZYmw | Male | Recovered | Infected | No lesions/Well |
| ZYfw | Female | Recovered | Infected | No lesions/Well |
| ZYfc2 | Female | Recovered | Infected | CIN-2 |
| ZYfc3 | Female | Recovered | Infected | CIN-3 |
| ZYfpcl | Female | Recovered | Infected | Preclinical local cancer |
| ZYfcl | Female | Recovered | Infected | Local cancer |
| ZYfpcr | Female | Recovered | Infected | Preclinical regional cancer |
| ZYfcr | Female | Recovered | Infected | Regional cancer |
| ZYfpcd | Female | Recovered | Infected | Preclinical distant cancer |
| ZYfcd | Female | Recovered | Infected | Distant cancer |
| ZZmw | Male | Recovered | Recovered | No lesions/Well |
| ZZfw | Female | Recovered | Recovered | No lesions/Well |
| H* | Female | - | - | Hysterectomy |
| CDL* | Female | - | - | Clinically detected local cancer |
| CDR* | Female | - | - | Clinically detected regional cancer |
| CDD* | Female | - | - | Clinically detected distant cancer |
| CSurv* | Female | - | - | Cancer survivorship no longer at risk of cancer death |

* These states do not follow the naming rules laid out in Figure 4.

#### 1.2 Parameter and variable definitions

The subscripts for the different subgropups of the model are described in Table 2.

Supplementary Table 2: Description of variables and subscripts

| Symbol | Description |
|---|---|
| Subscripts | |
| $k$ | Sex ($f$: female, $m$: male) |
| $l$, $s$ | Sexual activity group (*low*, *medium*, and *high*) |
| $i$, $j$ | Age group |
| $v$ | HPV type, either vaccine type ($v$) strains 6, 11, 16, 18, 31, 33, 45, 52, and 58 or non-vaccine type ($n$) all other high-risk HPV types |
| Variables | |
| $\lambda_{kliv}$ | Force of infection for sex $k$, in sexual activity group $l$, in age group $i$, for HPV type $v$ |

#### 1.3 Demographic model structure

The demographic structure is included in the system of ODEs. We divided the population into 24 age groups that would allow us to generate age-group-specific outcomes for policy evaluations and model calibration. The size of these age groups is based on life tables for the state being modeled. People then move across age groups at a rate $d_i$, which is calculated to keep the age groups a constant size over time, and face a mortality rate of $\mu_i$ based on state-specific life tables [29]. A more detailed explanation of the calculation of $d_i$ and the age groups can be found in [10].

Briefly, we assume that the population distribution has reached a steady state with exponential growth or decay of the form $e^{qt}A(a)$, where $A(a)$ is the age-distribution function, which provides a system of $n$ ordinary differential equations for the sizes of the $n$ age groups [12]. In this formulation, the maximum age is not explicitly defined because we assume that the last interval includes all the individuals over age $a_{n-1}$ [5, p. 267], [12, p. 623]. We assume that age in the model is piecewise constant for each year of age and $q = 0$. Let $P_i$ be the proportion of the population with ages in $[a_{i-1}, a_i]$. Then $P_i$ is given by [12, p. 623]:

$$P_i = \int_{a_{i-1}}^{a_i} A(a)\ da \tag{1}$$

#### 1.4 Sexual mixing

Age-specific sexual mixing preferences for those under 18 years of age was obtained from Kaestle, Morisky, and Wiley [15]; and for those over 18 years of age was obtained from Martinez et al. [19] and Chandra, Goodwin, and Mosher [9].

Sexual mixing in the model is done according to the proposed method in Easterly et al. [10]. The sexual activity weight matrix is defined as follows:

$$S_{kl} = \begin{cases} \epsilon_k & \text{if } l = s \\ \frac{1-\epsilon_k}{n_S - 1} & \text{if } l \neq s \end{cases} \tag{2}$$

where $0 \leq \epsilon_k \leq 1$ is the sexual activity assortativeness (mean preference of sexual activity groups for each other) for sex $k$, $n_S$ is the total number of sexual activity groups in the model, and $l$ and $s$ is the sexual activity group either high, medium, or low.

Next, $S_{kl}$ is modified by the availability of sexual partners to give the sexual activity mixing matrix $\Lambda_{kls}$:

$$\Lambda_{kls} = \frac{S_{k'ls} \sum_{a=1}^{n_A} c_{k'sa} N_{k'sa}}{\sum_{a=1}^{n_A} \sum_{z=1}^{n_S} S_{k'za} c_{k'za} N_{k'za}}, \tag{3}$$

where $k'$ indicates that people in sex $k$ is mixing with the opposite sex, $c_{ksj}$ is the sexual partner acquisition rate, $N_{ksj}$ is the total number of available partners, and $c_{ksj}N_{sj}$ is the "pool" of available partners for sex $k$, sexual activity group $s$, and age $j$.

Then, let the age mixing matrix $\Omega$ contain the same information for age as $\Lambda$ does for sexual activity. Assuming that age mixing and sexual activity mixing are independent of each other, the global mixing matrix is:

$$\rho_{klisj} = \Omega_{kij} \Lambda_{kls}, \tag{4}$$

where $\rho_{klisj}$ is the proportion of the $kli$ partners in the $k'sj$ group.

#### 1.5 Sex Partner Acquisition Rate (SPAR)

Data from the National Survey of Family Growth [7] was used to calculate the sex partner acquisition rate for each age group and sexual activity group. The high, middle, and low sexual activity groups were defined to be the top 5th percentile, the next 10%, and the bottom 85th percentile respectively. The rates were then weighted by age- and state-specific probability of being sexually active for males and females [8], and multiplied by a US sexual contact matrix to get an age distribution of sex partners based on age, sex, and sexual activity, e.g. a female of age $i$ in sexual activity group $l$ would have a certain number of sex partners in each age group $j$.

To account for inaccuracies in the data which result in the female and male SPARs being imbalanced, the SPARs are balanced using a method proposed in Easterly et al. [10], which uses the calibrated parameter $\theta$ which represents the weight given to the male SPAR in balancing, to calculate a weighted average of the male and female SPAR designated by $c^*_{klisj}$:

$$c^*_{mlisj} = \frac{\theta[c_{mli} \rho_{mlisj} N_{li}] + (1-\theta)[c_{fsj} \rho_{fsjli} N_{sj}]}{\rho_{mlisj} N_{li}}, \tag{5}$$

$$c^*_{flisj} = \frac{(1-\theta)[c_{fli} \rho_{flisj} N_{li}] + \theta[c_{msj} \rho_{msjli} N_{sj}]}{\rho_{flisj} N_{li}}, \tag{6}$$

#### 1.6 Force of infection (FOI)

The force of infection for individuals of sex $k$, hpv type $v$, age $i$, and sexual activity $l$ ($\lambda_{kvil}$), is defined as the annual rate of infection per susceptible individual. There are four calculated forces of infection in the model: $\lambda_{fvil}$, $\lambda_{mvil}$, $\lambda_{fnil}$, and $\lambda_{mnil}$. All FOIs are calculated by multiplying the probability that a partnership results in an infection ($\beta_{kv}$) by the number of sex partners per year and the probability that a partner is infected, where $c^*_{klisj}$ is the sex partner acquisition rate (after balancing as described in section 1.5) and $\rho_{klisj}$ is the choice of partner both of which are dependent on sex, sexual activity group, and age of both the choosing partner and the selected partner. Details on sexual mixing can be found in Section 1.4.

The FOI for vaccine-type HPV is defined as:

$$\lambda_{fvil} = \sum_{a=1}^{n_A}\sum_{z=1}^{n_S} \beta_{fv} c^*_{fliza}\rho_{fliza}\frac{[YXmw_{il} + YYmw_{il} + YZmw_{il} + rk(WXmw_{il} + WYmw_{il} + WZmw_{il})]}{N_{il}} \tag{7}$$

$$\lambda_{mvil} = \sum_{a=1}^{n_A}\sum_{z=1}^{n_S} \beta_{mv} c^*_{mliza}\rho_{mliza}\frac{[Y_{fvil} + rkW_{fvil}]}{N_{il}} \tag{8}$$

where $n_A$ is the total number of age groups; $rk$ is the probability of transmission from a vaccinated individual, relative to an unvaccinated individual; $Y_{fvil}$ is the total number of females who have vaccine-type HPV in age group $i$ and sexual activity group $l$; and $W_{fil}$ is the total number of females who have a breakthrough infection of vaccine-type HPV in age group $i$ and sexual activity group $l$. Both $Y_{fvil}$ and $W_{fil}$ are defined as follows:

$$\begin{aligned} Y_{fvil} = {} & YXfw_{il} + YXfc2_{il} + YXfc3_{il} + YXfpcl_{il} + YXfpcr_{il} + YXfpcd_{il} \\ & + YYfw_{il} + YYfc2_{il} + YYfc3_{il} + YYfpcl_{il} + YYfpcr_{il} + YYfpcd_{il} \\ & + YZfw_{il} + YZfc2_{il} + YZfc3_{il} + YZfpcl_{il} + YZfpcr_{il} + YZfpcd_{il} \end{aligned} \tag{9}$$

$$\begin{aligned} W_{fvil} = {} & WXfw_{il} + WXfc2_{il} + WXfc3_{il} + WXfpcl_{il} + WXfpcr_{il} + WXfpcd_{il} \\ & + WYfw_{il} + WYfc2_{il} + WYfc3_{il} + WYfpcl_{il} + WYfpcr_{il} + WYfpcd_{il} \\ & + WZfw_{il} + WZfc2_{il} + WZfc3_{il} + WZfpcl_{il} + WZfpcr_{il} + WZfpcd_{il} \end{aligned} \tag{10}$$

Similarly, the FOI for non-vaccine type HPV is defined as:

$$\lambda_{fnil} = \sum_{a=1}^{n_A}\sum_{z=1}^{n_S} \beta_{fn} c_{fliza}\rho_{fliza}\frac{[XYmw_{il} + YYmw_{il} + VYmw_{il} + WYmw_{il} + ZYmw_{il}]}{N_{mil}} \tag{11}$$

$$\lambda_{mnil} = \sum_{a=1}^{n_A}\sum_{z=1}^{n_S} \beta_{mn} c_{mliza}\rho_{mliza}\frac{Y_{fnil}}{N_{fil}} \tag{12}$$

where $Y_{fnil}$ is the total number of women who have non-vaccine type HPV in age group $i$ and sexual activity group $l$:

$$
\begin{aligned}
Y_{fnil} = {} & XYfw_{il} + XYfc2_{il} + XYfc3_{il} + XYfpcl_{il} + XYfpcr_{il} + XYfpcd_{il} \\
& + YYfw_{il} + YYfc2_{il} + YYfc3_{il} + YYfpcl_{il} + YYfpcr_{il} + YYfpcd_{il} \\
& + VYfw_{il} + VYfc2_{il} + VYfc3_{il} + VYfpcl_{il} + VYfpcr_{il} + VYfpcd_{il} \\
& + WYfw_{il} + WYfc2_{il} + WYfc3_{il} + WYfpcl_{il} + WYfpcr_{il} + WYfpcd_{il} \\
& + ZYfw_{il} + ZYfc2_{il} + ZYfc3_{il} + ZYfpcl_{il} + ZYfpcr_{il} + ZYfpcd_{il}
\end{aligned} \tag{13}
$$

#### 1.7 Progression rate

The rate of progression between states is represented by a Weibull hazard function. We calibrate both scale ($\zeta$) and shape ($k$) parameters specific for each transition, and use it to calculate age-specific progression rates ($tr$):

$$
tr_{iq}(\bar{a}_i; \zeta_q, k_q) = \zeta_q k_q \bar{a}_i^{k_q - 1}, \tag{14}
$$

where $i$ represents the age group; $q$ the transition between states: {1: well to CIN-2, 2: well to CIN-3, 3: CIN-2 to CIN-3, 4: CIN-3 to preclinical cancer}; and $\bar{a}_i$ the average age of the $i$-th age group.

#### 1.8 Screening

We implemented a cytology-based screening to the natural history model defined by the equations in Section 6, starting at age 21 and ending at age 65, with different women attending at different frequencies from every year to every 5 years or never being screened defined by the *p_screen* parameter. Women with CIN-2 or CIN-3 (i.e., CIN-$2^+$) will either be screened or not depending on their age. If they are not screened, the disease will progress following natural history rate. If they are screened and the disease is detected, they are treated by loop electrosurgical excision procedure (LEEP). If the LEEP treatment is effective, they will move to the **S**usceptible-**w**ell compartment. Alternatively, if the screening returns a false negative or treatment is not effective, the disease will progress following the natural history. (Figure 5).

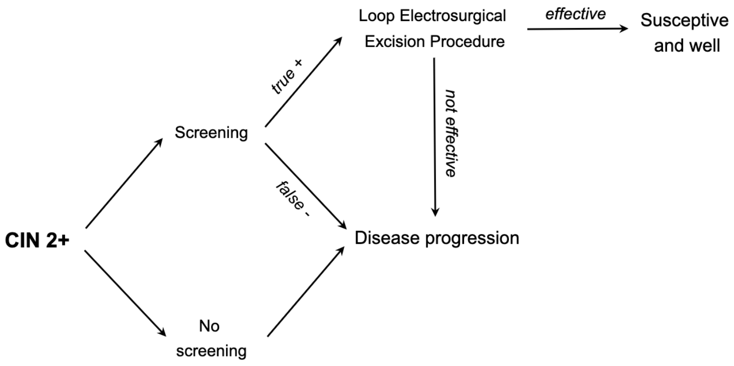

Supplementary Figure 5: Simplified screening algorithm for women with either CIN-2 or CIN-3 (i.e., CIN-$2^{+}$)

On the other hand, for women with preclinical cancer at any stage, screening can result in either detection and thus moving to a clinical cancer state or, if the screening results in a false negative, cancer will progress following the natural history (Figure 6).

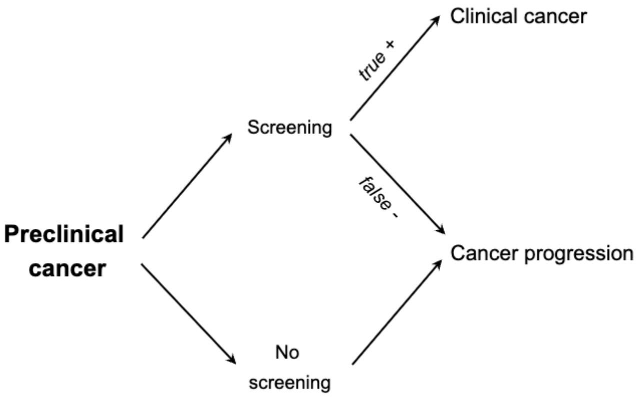

Supplementary Figure 6: Screening algorithm for women with preclinical cancer at any stage. Women will either be screened or not depending on their age. If they are not screened, the cancer will progress following the natural history. If they are screened and the cancer is detected they move to the cancer clinical compartment. Otherwise, if the screening returns a false negative the cancer will progress following the natural history.

### 2 Epidemiological outcomes

#### 2.1 Cervical cancer

##### 2.1.1 Cancer incidence

Cervical cancer incidence cases are calculated by summing the clinically detected cancers (i.e., when women transition from preclinical cancer to clinical cancer) over a given time interval. The incidence case equations (15) and (16) are the flow from the preclinical cancer compartments into the clinical cancer compartments, without any outflow. When they are integrated with respect to time ($t$), the equations give the total number of cases per person over a given time interval. The incidence is split into cancers caused by vaccine-type and non-vaccine-type HPV, $CIv_{il}$ and $CIn_{il}$, respectively. Cancer can develop when both HPV types are present, so we used proportional weight attribution to assign contributions from a single type [25]; $\xi$ represents the weight of vaccine-type HPV in multiple infections.

$$\begin{aligned}\frac{dCIv_{il}}{dt} &= detect_1(YXfpcl_{il} + \xi YYfpcl_{il} + YZfpcl_{il} + WXfpcl_{il} + \xi WYfpcl_{il} + WZfpcl_{il}) \\ &+ detect_2(YXfpcr_{il} + \xi YYfpcr_{il} + YZfpcr_{il} + WXfpcr_{il} + \xi WYfpcr_{il} + WZfpcr_{il}) \\ &+ detect_3(YXfpcd_{il} + \xi YYfpcd_{il} + YZfpcd_{il} + WXfpcd_{il} + \xi WYfpcd_{il} + WZfpcd_{il})\end{aligned} \tag{15}$$

$$\begin{aligned}\frac{dCIn_{il}}{dt} &= detect_1(XYfpcl_{il} + (1-\xi)YYfpcl_{il} + VYfpcl_{il} + (1-\xi)WYfpcl_{il} + ZYfpcl_{il}) \\ &+ detect_2(XYfpcr_{il} + (1-\xi)YYfpcr_{il} + VYfpcr_{il} + (1-\xi)WYfpcr_{il} + ZYfpcr_{il}) \\ &+ detect_3(XYfpcd_{il} + (1-\xi)YYfpcd_{il} + VYfpcd_{il} + (1-\xi)WYfpcd_{il} + ZYfpcd_{il})\end{aligned} \tag{16}$$

where $detect_q$ represents the detection rate in each cancer state $q$: {1: local, 2: regional, 3: distant}.

##### 2.1.2 Cancer Mortality

Cancer deaths caused by the vaccine- and non-vaccine-type HPV are counted into Equation (17), which represents the flow of deaths from every clinically detected cancer state. The number of deaths over a given time interval is calculated by integrating this equation with respect to time.

$$
\begin{aligned}
\frac{dCD_{il}}{dt} &= canc\_death_{1,1}(YXfcl_{il} + YYfcl_{il} + YZfcl_{il} + WXfcl_{il} \\
&+ WYfcl_{il} + WZfcl_{il} + XYfcl_{il} + VYfcl_{il} + ZYfcl_{il}) \\
&+ canc\_death_{2,1}(YXfcr_{il} + YYfcr_{il} + YZfcr_{il} + WXfcr_{il} \\
&+ WYfcr_{il} + WZfcr_{il} + XYfcr_{il} + VYfcr_{il} + ZYfcr_{il}) \\
&+ canc\_death_{3,1}(YXfcd_{il} + YYfcd_{il} + YZfcd_{il} + WXfcd_{il} \\
&+ WYfcd_{il} + WZfcd_{il} + XYfcd_{il} + VYfcd_{il} + ZYfcd_{il}) \\
&+ canc\_death_{1,2}CDL_{il,2} + canc\_death_{2,2}CDR_{il,2} + canc\_death_{3,2}CDD_{il,2} \\
&+ canc\_death_{1,3}CDL_{il,3} + canc\_death_{2,3}CDR_{il,3} + canc\_death_{3,3}CDD_{il,3} \\
&+ canc\_death_{1,4}CDL_{il,4} + canc\_death_{2,4}CDR_{il,4} + canc\_death_{3,4}CDD_{il,4} \\
&+ canc\_death_{1,5}CDL_{il,5} + canc\_death_{2,5}CDR_{il,5} + canc\_death_{3,5}CDD_{il,5} \\
&+ canc\_death_{1,6}CDL_{il,6} + canc\_death_{2,6}CDR_{il,6} + canc\_death_{3,6}CDD_{il,6} \\
&+ canc\_death_{1,7}CDL_{il,7} + canc\_death_{2,7}CDR_{il,7} + canc\_death_{3,7}CDD_{il,7} \\
&+ canc\_death_{1,8}CDL_{il,8} + canc\_death_{2,8}CDR_{il,8} + canc\_death_{3,8}CDD_{il,8} \\
&+ canc\_death_{1,9}CDL_{il,9} + canc\_death_{2,9}CDR_{il,9} + canc\_death_{3,9}CDD_{il,9} \\
&+ canc\_death_{1,10}CDL1_{il,10} + canc\_death_{2,10}CDR_{il,10} + canc\_death_{3,10}CDD_{il,10}
\end{aligned} \tag{17}
$$

where $canc\_death_{q,y}$ represents the age-specific cancer mortality rate at stage $q$: {1: for local, 2: for regional, and 3: for distant} depending on the survival year $y$: {1,2, . . . , 10}.

#### 2.2 Screened women

We computed the number of screened women by age and sexual activity group ($SCR_{il}$) in a period by integrating Equation (18) with respect to time, $t$.

$$
\begin{aligned}
\frac{dSCR_{il}}{dt} = p\_screen_i[&XXfw_{il} + XYfw_{il} + XZfw_{il} + YXfw_{il} + YYfw_{il} \\
&+ YZfw_{il} + VXfw_{il} + VYfw_{il} + VZfw_{il} + WXfw_{il} + WYfw_{il} \\
&+ WZfw_{il} + ZXfw_{il} + ZYfw_{il} + ZZfw_{il} + XYfc2_{il} + YXfc2_{il} \\
&+ YYfc2_{il} + YZfc2_{il} + VYfc2_{il} + WXfc2_{il} + WYfc2_{il} + WZfc2_{il} \\
&+ ZYfc2_{il} + XYfc3_{il} + YXfc3_{il} + YYfc3_{il} + YZfc3_{il} + VYfc3_{il} \\
&+ WXfc3_{il} + WYfc3_{il} + WZfc3_{il} + ZYfc3_{il} + XYfpcl_{il} + YXfpcl_{il} \\
&+ YYfpcl_{il} + YZfpcl_{il} + VYfpcl_{il} + WXfpcl_{il} + WYfpcl_{il} + WZfpcl_{il} \\
&+ ZYfpcl_{il} + XYfpcr_{il} + YXfpcr_{il} + YYfpcr_{il} + YZfpcr_{il} + VYfpcr_{il} \\
&+ WXfpcr_{il} + WYfpcr_{il} + WZfpcr_{il} + ZYfpcr_{il} + XYfpcd_{il} + YXfpcd_{il} \\
&+ YYfpcd_{il} + YZfpcd_{il} + VYfpcd_{il} + WXfpcd_{il} + WYfpcd_{il} + WZfpcd_{il} \\
&+ ZYfpcd_{il}]
\end{aligned} \tag{18}
$$

#### 2.3 False positives

For women who are well or have an HPV infection but no cervical disease, screening does not affect or change their health state in the model. However, false positives by age and sexual activity group ($FP_{il}$) are counted using the following equation:

$$
\begin{aligned}
\frac{dFP_{il}}{dt} = p\_screen_i(1 - spec\_screen)[&XXfw_{il} + XYfw_{il} + XZfw_{il} \\
&+ YXfw_{il} + YYfw_{il} + YZfw_{il} + VXfw_{il} + VYfw_{il} + VZfw_{il} \\
&+ WXfw_{il} + WYfw_{il} + WZfw_{il} + ZXfw_{il} + ZYfw_{il} + ZZfw_{il}]
\end{aligned} \tag{19}
$$

Equation (19) represents the total flow into the false positive compartment, and when integrated with respect to time, gives the number of false positives per person over a given time interval.

#### 2.4 Table of parameters

Table 3 describes the parameters of the different model subcomponents: demographic, sexual behavior, HPV dynamics, cervical disease, and screening.

Supplementary Table 3: Description of parameters. The *Value* column indicates either values (i) from the literature, (ii) assumed, or (iii) the mean (2.5% quantile, 97.5% quantile) of the posteriori estimates from the Bayesian calibration (see Section 3.1).

| Symbol | Description | Value | Source | Prior Dist. (Calibration) |
|---|---|---|---|---|
| **Demographic parameters** | | | | |
| $b$ | Birth rate; entry rate into youngest age group | 0.0129 | Section 1.3 | - |
| $\mu_i$ | Background mortality for age group $i$ | Varies with age | Section 1.3 | - |
| $d_i$ | Aging rate out of age group $i$ | Varies with age | Section 1.3 | - |
| $h_i$ | Hysterectomy rate | Varies with age | [20] | - |
| $n_A$ | Number of age groups | 24 | Assumed | - |
| Age groups | Model ages | 0-11,12,13,14, 15,16,17,18,19, 20,21,22,23,24, 25,26,27-31,32-36, 37-41,42-46,47-56, 57-66,67-76,77+ | Assumed | - |
| $n_S$ | Number of sexual activity groups: high, medium, low | 3 | Assumed | - |
| **Sexual behavior parameters** | | | | |
| $\epsilon_k$ | Sexual activity assortativeness for sex $k$ | 0.7 (F)<br>0.7 (M) | Assumed<br>Assumed | -<br>- |
| $c_{kli}$ | Sex partner | Varies with age | Section 1.5 | - |

Description of parameters (continued)

| Symbol | Description | Value | Source | Prior Dist. (Calibration) |
|---|---|---|---|---|
| | acquisition rate (SPAR) | and sexual activity group | | |
| $\rho_{klimj}$ | Probability of a person of sex $k$, sexual activity $l$, and age $i$ choosing someone of sex $k'$, sexual activity $m$, and age $j$ | Varies with age and sexual activity group | Section 1.4 | - |
| $\theta$ | Weight given to male SPAR in SPAR balancing | 0.792 (0.755, 0.822) | Calibrated | Unif(0.6, 1.0) |
| $\omega_l$ | Proportion in sexual activity group $l$: 1 is high, 2 is medium, 3 is low | 0.05 (1)<br>0.10 (2)<br>0.85 (3) | Assumed | - |
| Age of sexual viability | Individuals become sexually active after this age | 12 | Assumed | - |
| Average age of first sex | Average age of becoming sexually active | 17.2 | Assumed | - |
| **HPV dynamics parameters** | | | | |
| $\beta_{kv}$ | Transmission probability of HPV type $v$ from sex $k$ | 0.683 (0.650, 0.712) (vF)<br>0.812 (0.757, 0.876) (nF)<br>0.381 (0.358, 0.413) (vM)<br>0.847 (0.804, 0.891) (nM) | Calibrated | Unif(0.35, 0.95) |
| $\lambda_{kv}$ | Force of infection of HPV type $v$ and sex $k$ | Varies with age and sexual activity group | Section 1.6 | - |

Description of parameters (continued)

| Symbol | Description | Value | Source | Prior Dist. (Calibration) |
|---|---|---|---|---|
| $\tau_{kv}$ | Rate of waning of natural immunity for sex $k$ when recovered from HPV type $v$ | 0 | [6] | - |
| $1/\gamma_{kv}$ | Duration of infectiousness for individuals of sex $k$ when infected with HPV type $v$ (years):<br>1 is age group 11-20<br>2 is age group 21-26<br>3 is age group 27-46<br>4 is age group 47+ | 0.21 (0.17, 0.26) (vF)(1)<br>0.45 (0.41, 0.51) (vF)(2)<br>1.06 (0.98, 1.15) (vF)(3)<br>1.28 (1.17, 1.37) (vF)(4)<br>1.20 (1.12, 1.28) (nF)(1)<br>1.06 (1.00, 1.11) (nF)(2)<br>1.85 (1.75, 1.97) (nF)(3)<br>1.85 (1.75, 1.97) (nF)(4)<br>0.70 (0.53, 0.84) (vM)(1)<br>0.60 (0.47, 0.72) (vM)(2)<br>1.55 (1.44, 1.66) (vM)(3)<br>1.12 (0.94, 1.28) (vM)(4)<br>0.21 (0.14, 0.28) (nM)(1)<br>0.89 (0.82, 0.97) (nM)(2)<br>0.22 (0.19, 0.26) (nM)(3)<br>1.26 (1.16, 1.35) (nM)(4) | Calibrated<br>Calibrated<br>Calibrated<br>Calibrated<br>Calibrated<br>Calibrated<br>Calibrated<br>Calibrated<br>Calibrated<br>Calibrated<br>Calibrated<br>Calibrated<br>Calibrated<br>Calibrated<br>Calibrated<br>Calibrated | Unif(0.4, 10.0)<br>Unif(0.4, 10.0)<br>Unif(0.4, 10.0)<br>Unif(0.4, 10.0)<br>Unif(0.4, 10.0)<br>Unif(0.4, 10.0)<br>Unif(0.4, 10.0)<br>Unif(0.4, 10.0)<br>Unif(0.4, 10.0)<br>Unif(0.4, 10.0)<br>Unif(0.4, 10.0)<br>Unif(0.4, 10.0)<br>Unif(0.4, 10.0)<br>Unif(0.4, 10.0)<br>Unif(0.4, 10.0)<br>Unif(0.4, 10.0) |
| $\nu_k$ | Proportion of individuals of sex $k$ that develop immunity | 0.596 (0.582, 0.611) (F)<br>0 (M) | Calibrated<br>Assumed | Unif(0.4, 0.7) (F)<br>- |
| **Vaccine characteristics** | | | | |
| $\phi_{kil}$ | Vaccination rate | Varies with age sex and sexual activity | Assumed | - |
| $\delta_k$ | Vaccine efficacy: probability of | 0 | [14] | - |

Description of parameters (continued)

| Symbol | Description | Value | Source | Prior Dist. (Calibration) |
|---|---|---|---|---|
| | getting vaccine type HPV if vaccinated | | | |
| $\sigma$ | Rate of waning of vaccine protection | 0 | Assumed | - |
| $\alpha$ | Post-vaccination rate of infection, clearance relative to unvaccinated | 1 | Assumed | - |
| $\upsilon$ | Effectiveness of the vaccine if someone is already infected with vaccine-type HPV | 0 | Assumed | - |
| $rk$ | Probability of transmission of a vaccinated individual, relative to unvaccinated | 1 | Assumed | - |

Description of parameters (continued)

| Symbol | Description | Value | Source | Prior Dist. (Calibration) |
|---|---|---|---|---|
| **Cervical disease parameters** | | | | |
| $\zeta_q$ | Scale of the Weibull distribution* of the progression rate between states $q$: 1 is well to CIN-2, 2 is well to CIN-3, 3 is CIN-2 to CIN-3, and 4 is CIN-3 to preclinical cancer | 0.0063 (0.0058, 0.0067) (1)<br>0 (2)<br>0.0087 (0.0084, 0.0091) (3)<br>0.00093 (0.00085, 0.00100) (4) | Calibrated<br>Assumed<br>Calibrated<br>Calibrated | Unif(0.0002, 0.009) (<br>-<br>Unif(0.0040, 0.012) (<br>Unif(0.0005, 0.002) ( |
| $k_q$ | Shape of the Weibull distribution* of the progression rate between states $q$: 1 is well to CIN-2, 2 is well to CIN-3, 3 is CIN-2 to CIN-3, and 4 is CIN-3 to preclinical cancer | 1.22 (1.20, 1.23) (1)<br>0 (2)<br>1.38 (1.33, 1.42) (3)<br>1.37 (1.36, 1.39) (4) | Calibrated<br>Assumed<br>Calibrated<br>Calibrated | Unif(1.002, 1.65) (1<br>-<br>Unif(1.002, 1.75) (3<br>Unif(1.100, 1.50) (4 |
| $tr_{iq}$ | Progression rate between states $q$: 1 is well to CIN-2, 2 is well to CIN-3, 3 is CIN-2 to CIN-3, and 4 is CIN-3 to preclinical cancer | Varies by age and state | Section 1.7 | - |
| $reg\_n_q$ | Recovery rate of non-vaccine type HPV infection $q$: 1 is from CIN-2 to well, and 2 is from CIN-3 | 0.0127 (0.0117, 0.0139) (1)<br>0.0036 (0.0031, 0.0040) (2) | Calibrated<br>Calibrated | Unif(0.0010, 0.015) (<br>Unif(0.0005, 0.010) ( |

Description of parameters (continued)

| Symbol | Description | Value | Source | Prior Dist. (Calibration) |
| --- | --- | --- | --- | --- |
| | to CIN-2 | | | |

Description of parameters (continued)

| Symbol | Description | Value | Source | Prior Dist. (Calibration) |
|---|---|---|---|---|
| $hr\_reg\_v_q$ | Hazard ratio of recovery rate for vaccine type HPV infection $q$, relative to non-vaccine type: 1 is from CIN-2 to well, and 2 is from CIN-3 to CIN-2 | 0.38 (0.32, 0.46) (1)<br>0.65 (0.61, 0.69) (2) | Calibrated<br>Calibrated | Unif(0.1, 1.0) (1)<br>Unif(0.1, 1.0) (2) |
| $canc\_prog_q$ | Cancer progression rate $q$: 1 is from local to regional cancer, and 2 is from regional to distant cancer | 0.58 (1)<br>0.92 (2) | [26] | - |
| $detect_q$ | Cancer detection rate $q$: 1 is for local 2 is for regional, and 3 is for distant cancer | 0.15 (1)<br>0.41 (2)<br>0.90 (3) | [26] | - |
| $canc\_surv_q$ | Cancer survival rate $q$ (conditional in the model): 1 is for local, 2 is for regional, and 3 is for distant cancer | Varies with age and survival year | [22] | - |
| $canc\_death_q$ | Cancer death rate $q$ (conditional in the model): 1 is for local, 2 is for regional, and 3 is for distant cancer | Varies with age and survival year | [22] | - |

Description of parameters (continued)

| Symbol | Description | Value | Source | Prior Dist. (Calibration) |
|---|---|---|---|---|
| $HR_q$ | Hazard ratio of disease progression rate for vaccine type HPV relative to non-vaccine type HPV between states $q$: 1 is well and CIN-2, 2 is CIN-2 and CIN-3, and 3 is CIN-3 and cancer | 2.02 (1.96, 2.09) (1)<br>2.06 (1.95, 2.17) (2)<br>1.56 (1.47, 1.66) (3) | Calibrated<br>Calibrated<br>Calibrated | Unif(1.0, 2.5) (1)<br>Unif(1.2, 3.0) (2)<br>Unif(1.2, 3.0) (3) |
| **Screening parameters** | | | | |
| $p\_screen$ | Probability of a person being screened for cervical lesions | Varies by age | Section 1.8 | - |
| $p\_eff\_leep$ | Probability of Loop Electrosurgical Excision Procedure (LEEP) treatment being successful | 0.93 | [4] | - |
| $spec\_screen$ | Specificity of cytology in detecting disease (CIN-$2^+$) | 0.919 | [18] | - |
| $sens\_scn\_cin$ | Sensitivity of cytology in detecting disease (CIN-$2^+$) | 0.727 (0.707, 0.750) | [18] | - |
| $sens\_scn\_cc$ | Sensitivity of cytology for | 0.727 (0.707, 0.750) | [26] | - |

Description of parameters (continued)

| Symbol | Description | Value | Source | Prior Dist. (Calibration) |
|---|---|---|---|---|
| | cancer | | | |

[*] Weibull distribution

$$f(x;\zeta,k) = \begin{cases} \frac{k}{\zeta}\left(\frac{x}{\zeta}\right)^{k-1} e^{(-x/k)^k} & \text{if } x \geq 0 \\ 0 & \text{otherwise} \end{cases}$$

### 3 Parameter estimation

Parameters of mathematical models can be either unobserved or unobservable for various reasons (e.g., financial, practical, or ethical). Model calibration is the process of estimating values for unknown or uncertain parameters of a mathematical model by matching model outputs to observed clinical or epidemiological data (known as calibration targets). The goal is to identify parameter values that maximize the fit between model outputs and the calibration targets. Accordingly, we calibrated the 35 model parameters of the natural history part of the CISNET CERVIX Collaborative (C3) model to the observed target data using a Bayesian approach. The parameters concerned HPV transmission and cervical cancer natural history and are listed in Table 3. We assumed that the natural history of cervical cancer was identical across states after accounting for differences in vaccination and screening rates, so these parameters were obtained from calibration to the national data. Vaccination rates, screening rates, population age structure, were free to vary across states.

#### 3.1 Calibration targets

Calibration inferred values for model parameters by matching modeled outcomes to the calibration targets: sex-, age-, and HPV-type specific HPV prevalence, age-specific CIN-2 and CIN-3 prevalence, cervical incidence, and proportion of cancers that are vaccine type, described in Table 4.

Supplementary Table 4: Calibration targets

| Age group | Mean | 95% CI | Source |
|---|---|---|---|
| **Female vaccine-type HPV prevalence** | | | |
| <21 | 0.224 | (0.21, 0.23) | [30] |
| 21 - 24 | 0.213 | (0.20, 0.22) | |
| 25 - 29 | 0.142 | (0.14, 0.15) | |
| 30 - 49 | 0.069 | (0.07, 0.07) | |
| 50 - 70 | 0.041 | (0.04, 0.05) | |

Calibration targets (continued)

| Age group | Mean | 95% CI | Source |
|---|---|---|---|
| **Female non vaccine-type HPV prevalence** | | | |
| <21 | 0.131 | (0.12, 0.14) | [30] |
| 21 - 24 | 0.112 | (0.11, 0.12) | |
| 25 - 29 | 0.076 | (0.07, 0.08) | |
| 30 - 49 | 0.046 | (0.04, 0.05) | |
| 50 - 70 | 0.028 | (0.02, 0.03) | |
| **Male vaccine-type HPV prevalence** | | | |
| 18 - 19 | 0.065 | (0.03, 0.10) | [11] |
| 20 - 24 | 0.078 | (0.03, 0.12) | |
| 25 - 29 | 0.089 | (0.04, 0.14) | |
| 30 - 34 | 0.093 | (0.04, 0.15) | |
| 35 - 39 | 0.082 | (0.04, 0.13) | |
| 40 - 44 | 0.083 | (0.04, 0.13) | |
| 45 - 70 | 0.077 | (0.03, 0.12) | |
| **Male non vaccine-type HPV prevalence** | | | |
| 18 - 19 | 0.131 | (0.09, 0.17) | [11] |
| 20 - 24 | 0.155 | (0.11, 0.20) | |
| 25 - 29 | 0.177 | (0.13, 0.23) | |
| 30 - 34 | 0.185 | (0.13, 0.24) | |
| 35 - 39 | 0.164 | (0.12, 0.21) | |
| 40 - 44 | 0.165 | (0.12, 0.21) | |
| 45 - 70 | 0.153 | (0.11, 0.20) | |
| **Cervical cancer incidence (per 100,000)** | | | |
| 10 - 14 | 0.30 | (0, 0.60) | [26] |
| 15 - 19 | 2.65 | (1.37, 3.93) | |
| 20 - 24 | 3.96 | (1.32, 6.59) | |
| 25 - 29 | 16.82 | (13.45, 20.18) | |
| 30 - 34 | 32.12 | (28.16, 36.08) | |
| 35 - 39 | 43.52 | (38.77, 48.27) | |
| 40 - 44 | 49.88 | (44.82, 54.94) | |
| 45 - 49 | 52.06 | (46.37, 57.76) | |
| 50 - 54 | 52.02 | (45.68, 58.37) | |
| 55 - 59 | 50.25 | (43.53, 56.97) | |
| 60 - 64 | 46.65 | (39.63, 53.67) | |
| 65 - 69 | 43.95 | (35.72, 52.17) | |
| 70 - 74 | 42.66 | (32.99, 52.32) | |

Calibration targets (continued)

| Age group | Mean | 95% CI | Source |
|---|---|---|---|
| **Proportion of cancers that are vaccine type** | | | |
| <35 | 0.900 | (0.85, 0.95) | [25] |
| 35 - 44 | 0.894 | (0.85, 0.93) | |
| 35 - 54 | 0.784 | (0.72, 0.84) | |
| 55 - 64 | 0.759 | (0.68, 0.84) | |
| 65+ | 0.673 | (0.60, 0.75) | |
| **CIN 2 prevalence (per 100)** | | | |
| 21 - 24 | 4.95 | (3.45, 6.46) | [26] |
| 25 - 34 | 4.00 | (2.64, 5.37) | |
| 35 - 44 | 1.24 | (0.65, 1.82) | |
| 45+ | 0.81 | (0.38, 1.22) | |
| **CIN 3 prevalence (per 100)** | | | |
| 21 - 24 | 1.50 | (1.0, 2.0) | [26] |
| 25 - 34 | 3.00 | (2.0, 4.0) | |
| 35 - 44 | 3.50 | (2.5, 4.5) | |
| 45+ | 3.50 | (2.5, 4.5) | |

##### 3.2 Incremental Mixture Importance Sampling

We used the incremental mixture importance sampling (IMIS) algorithm [27, 23], a Bayesian method, to calibrate 35 model parameters that could not be directly estimated from data. IMIS has been previously used to calibrate health policy models [24, 26]. The parameters concerned HPV transmission and cervical cancer natural history, such as progression and regression rates. Briefly, we sampled 10,000 parameter sets from our priors in the first stage, followed by 1,000 samples for up to a maximum of 500 consecutive updating sampling stages. This procedure yielded a posterior distribution from which we obtained 1,000 samples, used for our projections and analyses. The marginal posterior distributions are shown in Supplementary Figure 7.

Calibration inferred values for model parameters by matching model-predicted outcomes to the observed calibration targets. We measured the goodness of fit of the model-predicted outcomes against empirical data using a likelihood function, which we constructed by assuming that targets follow normal distributions with means given by the model-predicted outputs and the standard errors from empirical data. We defined uniform prior distributions for all calibrated parameters with ranges based on existing evidence, epidemic theory, and plausibility. Calibration resulted in a joint posterior distribution with uncertainty for the calibrated model parameters, from which we obtained 1,000 samples for our projections and analyses. The marginal posterior distributions are shown in Figure 7.

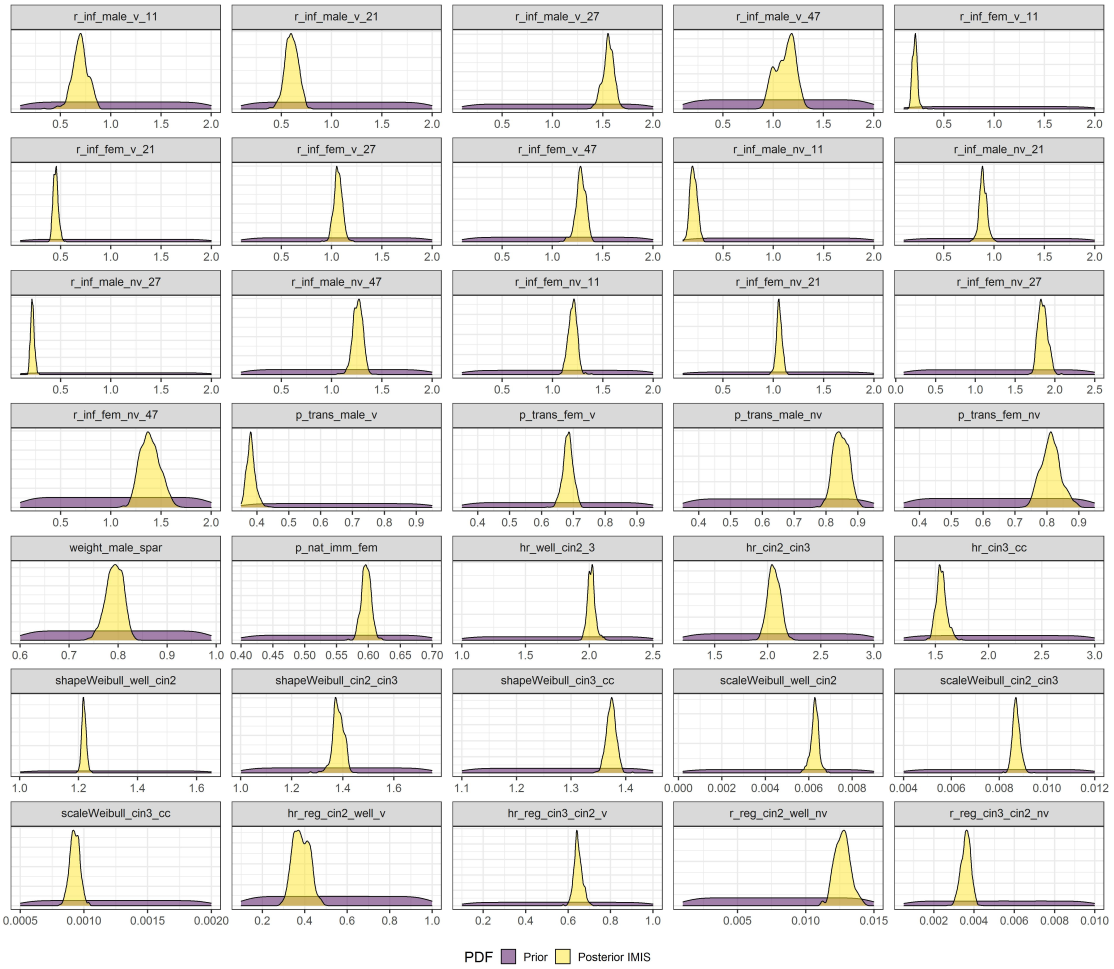

Supplementary Figure 7: Comparison of the prior vs posterior distributions of the 35 calibrated parameters

#### 3.3 Likelihood function

The goodness of fit of the model-predicted outcomes against empirical data was measured by a likelihood function, which we constructed by assuming that targets follow normal distributions with means given by the model-predicted outputs and the standard errors from empirical data.

#### 3.4 Priors

The prior distributions represent the information on the parameters before these are updated by the epidemiological calibration targets. We defined uniform prior distributions for all calibrated parameters with ranges based on existing evidence, epidemic theory, and plausibility (Table 3). Calibration resulted in an estimate of the joint posterior uncertainty distribution for the model parameters.

### 4 Model validation

To validate the C3 model, we conducted both an internal and external validation [1] propagating the uncertainty of the calibrated parameters by randomly drawing 1,000 parameter sets from their joint posterior distribution obtained from the IMIS algorithm.

#### 4.1 Internal validation

For internal validation, the calibrated parameter sets were used to run the model for 140 years to ensure it had reached a steady state, and then the output was compared to the calibration targets. Figures 8, 9 and 10 show the fit of the model-predicted outcomes and the calibration targets.

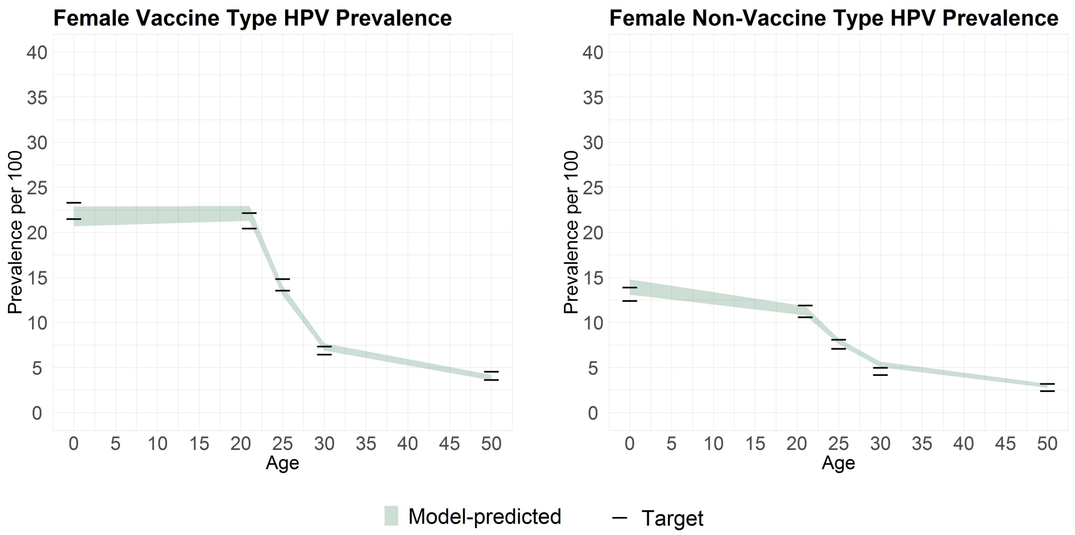

Supplementary Figure 8: Internal validation for female vaccine- and non-vaccine-type HPV prevalence. Shaded areas shows the 95% posterior model-predictive interval of the outcome.

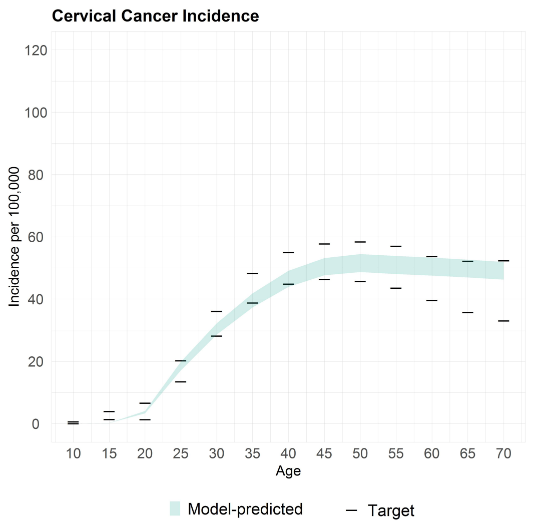

Supplementary Figure 9: Internal validation for cervical cancer incidence. Shaded areas shows the 95% posterior model-predictive interval of the outcome.

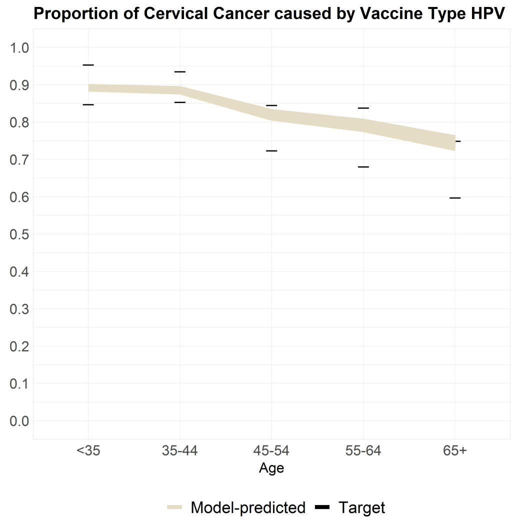

Supplementary Figure 10: Internal validation for the proportion of cervical cancer caused by the vaccine-type HPV. Shaded areas shows the 95% posterior model-predictive interval of the outcome.

#### 4.2 External validation

##### 4.2.1 Cervical cancer and mortality

To externally validate the natural history of the C3 model, we took a two-fold approach: First, we modeled a cytology-based screening algorithm under different screening frequencies in combination with the C3 natural history model and compared the model-predicted incidence and mortality (with all women alive included in the denominator) to the U.S. Surveillance, Epidemiology and End Results (SEER) data [28] and state-specific data [28] on cervical cancer incidence and mortality. We use data from Sawaya et al. [26] on screening frequencies – including non-adherence [17] to individuals who never screen – to run and compute a weighted average of the different screening scenarios, using as weights the proportions of the population by the frequency of screening every 1, 2, 3, 4 and 5 years or that have never been screened (Table 5).

To propagate the uncertainty on both the screening frequencies and test characteristics, we conducted an uncertainty analysis by randomly drawing the natural history parameters from their joint posterior distribution. For the sensitivity of the screening test, we assigned a beta distribution. For the frequency of screening, we assumed a Dirichlet distribution where we calculated the parameters using a method of moments [21] using the means and standard errors obtained from the lower and upper bounds of the confidence intervals previously reported.

We then compared the output of the model evaluated at 1,000 different combinations of parameter sets with the screening algorithm described above to the minimum and maximum values of cervical cancer incidence and mortality in SEER from 2003-2013.

Supplementary Table 5: US screening frequency and proportion

| Scenario | Frequency of screening | Proportion of population (95% CI) |
|---|---|---|
| 1 | Never screened | 14.4% (7.2, 21.5) |
| 2 | Screened in the past year | 9.20% (3.6, 14.9) |
| 3 | Screened in the past two years | 16.2% (11.3, 21.1) |
| 4 | Screened in the past three years | 10.6% (3.6, 17.6) |
| 5 | Screened in the past four years | 35.2% (26.4, 44.0) |
| 6 | Screened in the past five years | 14.4% (7.2, 21.5) |

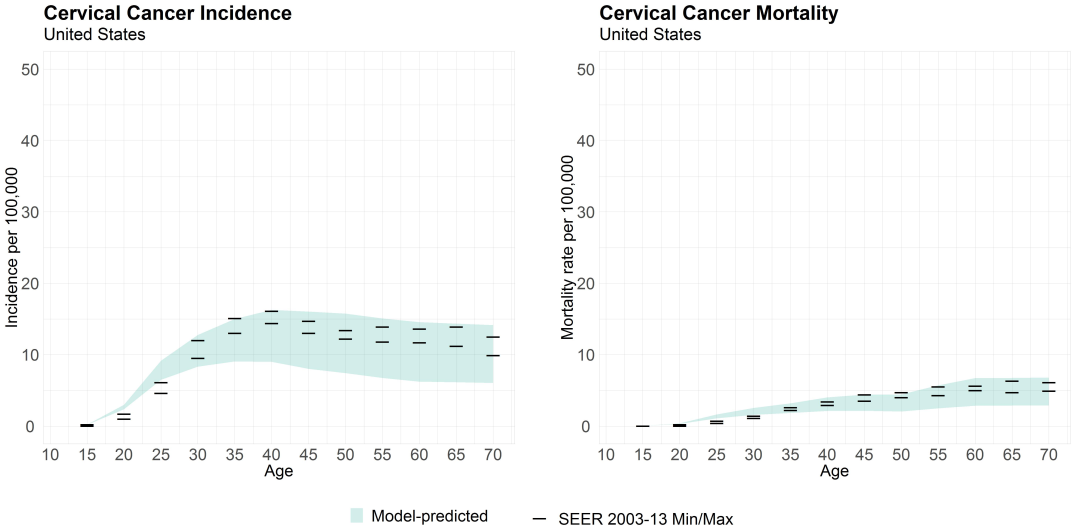

Supplementary Figure 11: External validation. Cervical cancer incidence and mortality in the US. Shaded areas shows the 95% posterior model-predictive interval of the outcome.

State-specific screening weights can be approximated to maintain the same value across the U.S. weights when a relative proportion of screened individuals by state is applied. We used the state-specific percentage of screened women within the last three years from America's Health Rankings [3] to compute these relative proportions and propose the following equation

$$\sum_{i=1}^{6} w_i^s p_{it} = \sum_{i=1}^{6} w_i^{US} RP^s p_{it}, \tag{20}$$

where the subscript $s$ refers to a U.S. state; the subscript $i$ denotes each one of the six scenarios presented in Table 5; $RP^s$ is a relative proportion of the percentage of screened women within the last three years between the state $s$ and the U.S.; $p_{it}$ is the proportion of screened female individuals at a rate $i$ at time $t$ (e.g., $p_{i3} = p_i(3) = 1 - exp(-3 \cdot \text{rate}_i)$)

Equation (20) has infinite solutions; however, it can be solved numerically by adding constraints (21)-(24) to narrow the parameter space. We defined the constraints (23) and (24) because we expect the proportion of screened individuals to be higher for the U.S. than for a particular state $s$.

$$0 \leq w_i^s \leq 1 \quad (21)$$

$$\sum_{i=1}^{6} w_i^s = 1 \quad (22)$$

$$w_i^s > w_i^{US} \quad \text{if } i = 1, 5, 6 \quad (23)$$

$$w_i^s < w_i^{US} \quad \text{if } i = 2, 3, 4 \quad (24)$$

We solved Equation (20) using the Nelder Mead algorithm by adding the constraints in (21) -(24). To solve the equation, we drew 1,000 random starting points from a uniform distribution. We selected the vector $w_i^s$ with the closest value to the solution. We present our results in Table 6.

Supplementary Table 6: State-specific screening proportions*

| Frequency of screening | US | California | New York | Texas |
|---|---|---|---|---|
| Never screened | 0.174 | 0.174 | 0.177 | 0.181 |
| Screened in the past year | 0.089 | 0.089 | 0.089 | 0.062 |
| Screened in the past two years | 0.156 | 0.067 | 0.060 | 0.127 |
| Screened in the past three years | 0.102 | 0.096 | 0.100 | 0.068 |
| Screened in the past four years | 0.340 | 0.396 | 0.430 | 0.422 |
| Screened in the past five years | 0.139 | 0.179 | 0.144 | 0.140 |

*Taking into account non-adherence to colposcopy.

Finally, we used each of the 1,000 calibrated parameter to run the six scenarios by state and compute a weighted average across them. Figures 12, 13, and 14 show the external validation of our model to state-specific cervical cancer incidence and mortality.

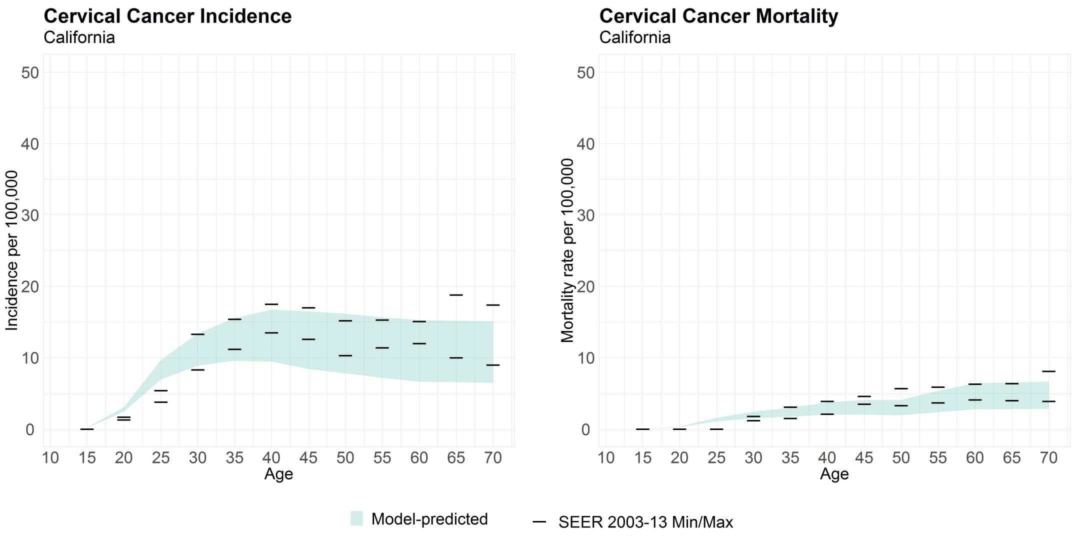

Supplementary Figure 12: External validation. Cervical cancer incidence and mortality in California. Shaded areas shows the 95% posterior model-predictive interval of the outcome.

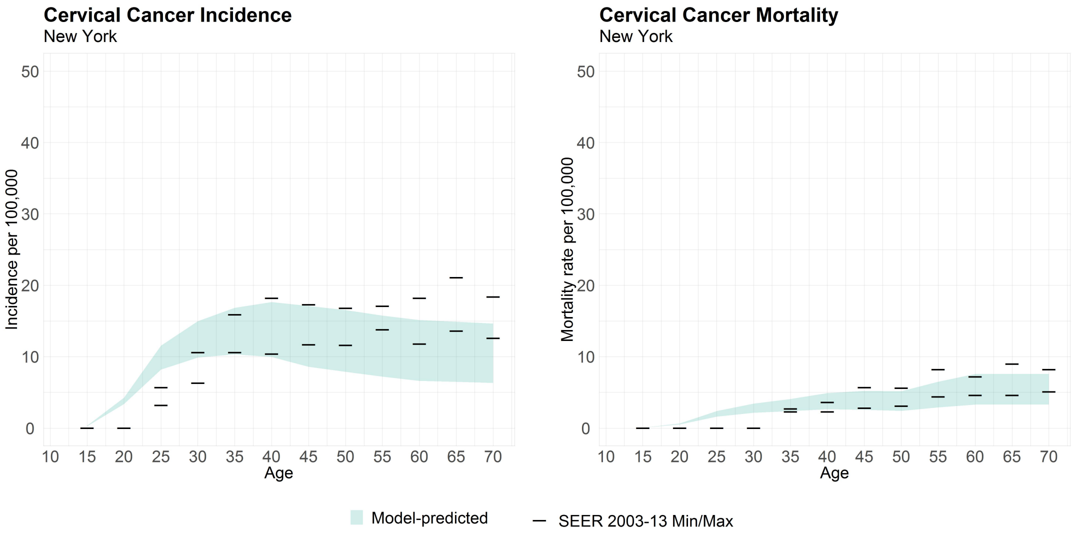

Supplementary Figure 13: External validation. Cervical cancer incidence and mortality in New York. Shaded areas shows the 95% posterior model-predictive interval of the outcome.

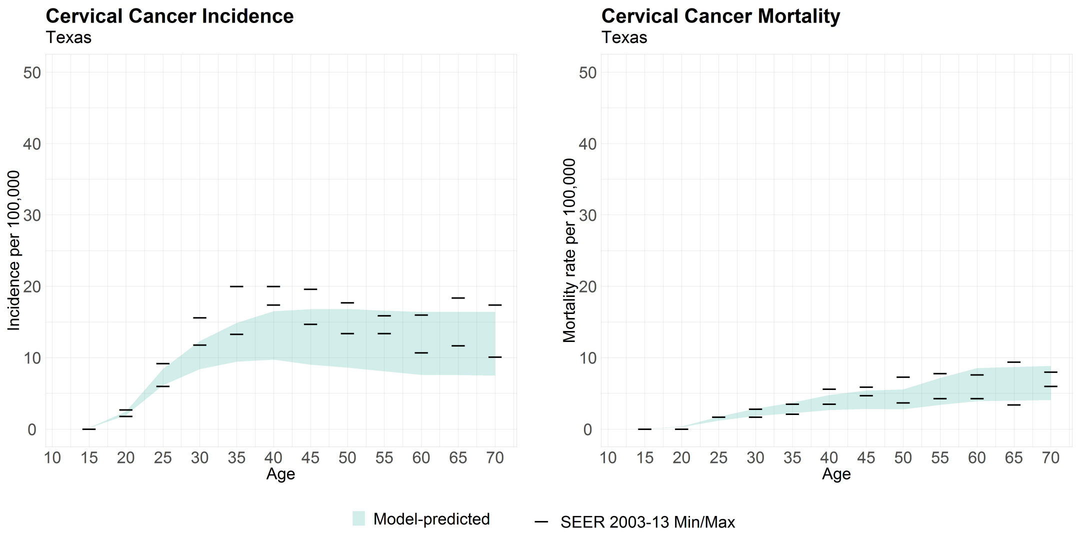

Supplementary Figure 14: External validation. Cervical cancer incidence and mortality in Texas. Shaded areas shows the 95% posterior model-predictive interval of the outcome.

###### 4.2.2 Vaccination

We externally validated the vaccination component of the model by simulating various vaccination strategies, including vaccinating girls only, and girls and boys at age 12 years, and predicting the population-level impact in terms of reduction in vaccine-type HPV prevalence, and compared them to those predicted by Brisson et al. [6] (Figure 15). We ran the model to a steady state using the calibrated parameters and recorded the female prevalence of the vaccine-type HPV for women between 15 and 57 years old over 70 years.

The vaccination scenarios were defined as in Brisson et al. [6]:

A 40% of girls age 12 vaccinated

B 80% of girls age 12 vaccinated

C 40% of girls and boys age 12 vaccinated

D 80% of girls and boys age 12 vaccinated

We computed the relative reduction ($rRed$) for scenarios A-B, and the incremental relative reduction for scenarios C and D in female vaccine-type HPV prevalence. We assume 100% vaccine efficacy.

$$rRed_i = \frac{X_{SQ} - X_{vax_i}}{X_{SQ}} \times 100\% \tag{25}$$

where the status quo scenario ($SQ$) is defined as no vaccination.

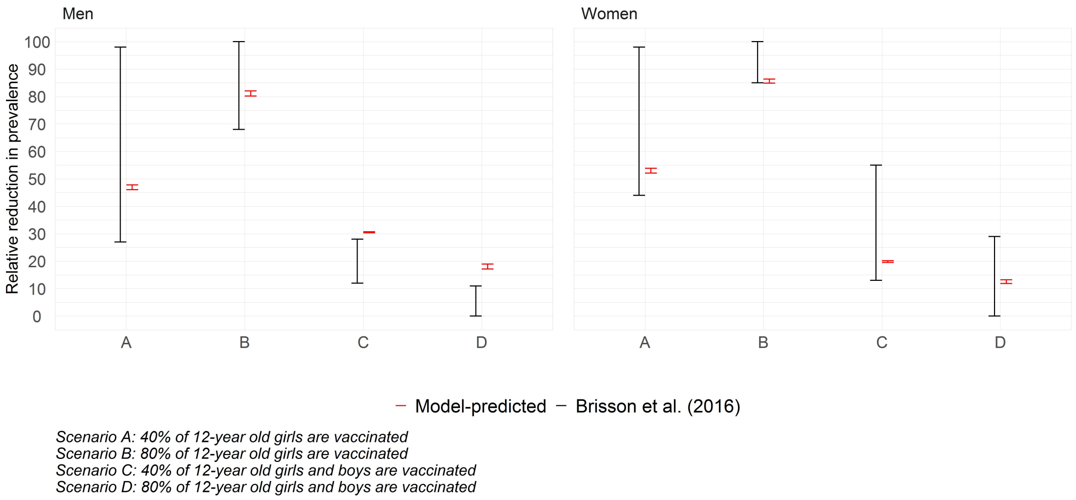

Supplementary Figure 15: External validation for vaccination.

### 5 Additional results

#### 5.1 Hexamaps visualizing cervical cancer incidence for each scenario as a function of age, calendar year, and birth year

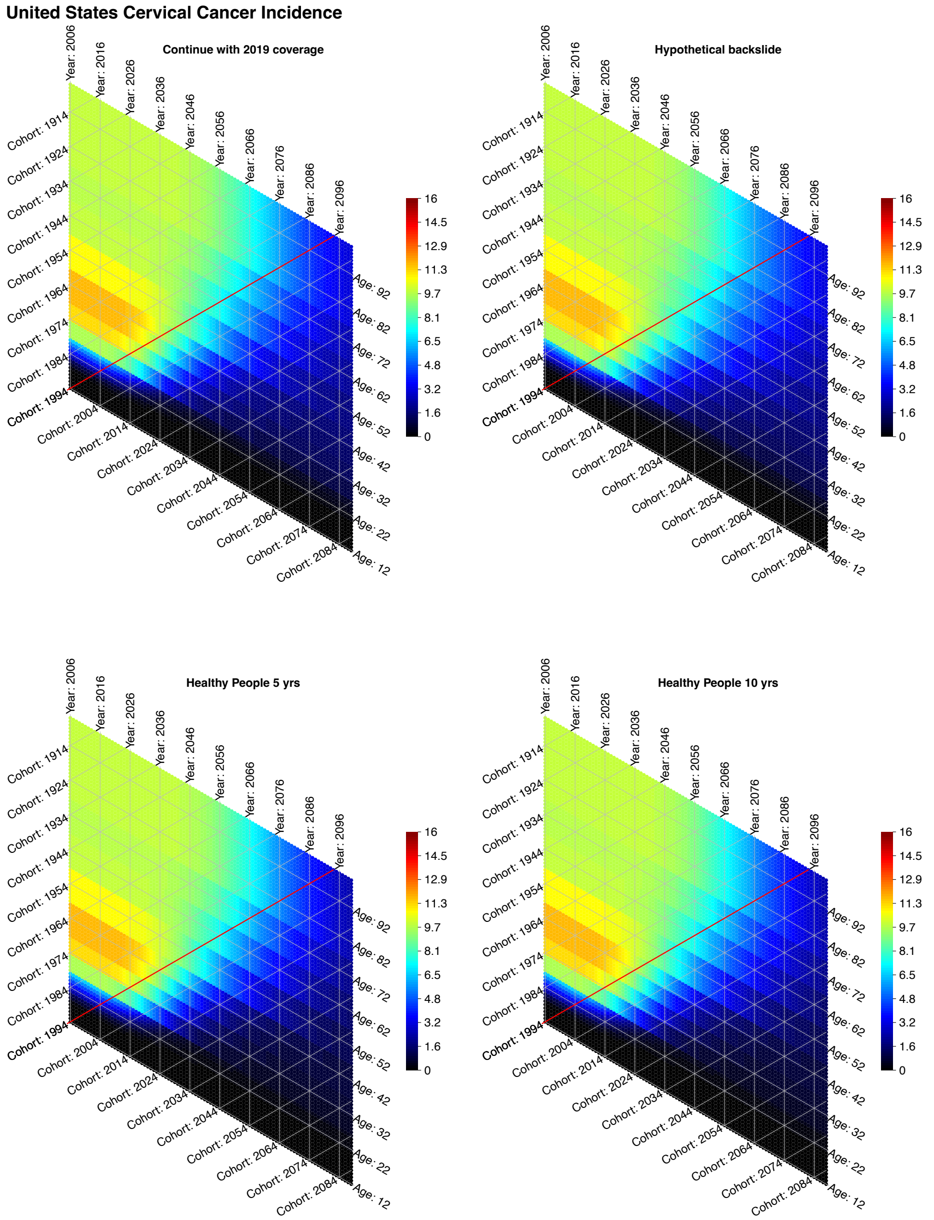

Supplementary Figure 16: Hexamaps visualizing cervical cancer incidence rate per 100,000 population in the US as a function of age, calendar year, and birth year in the data. The vertical lines represent calendar years, the horizontal lines represent ages, and the diagonal lines represent the aging of each birth cohort with calendar years from the bottom left to the top right. In each scenario, the red diagonal line represents the birth cohort that became eligible to be vaccinated from the start of the simulation. Cohorts born after 1994 are represented by less heated colors at the bottom right of the red line.

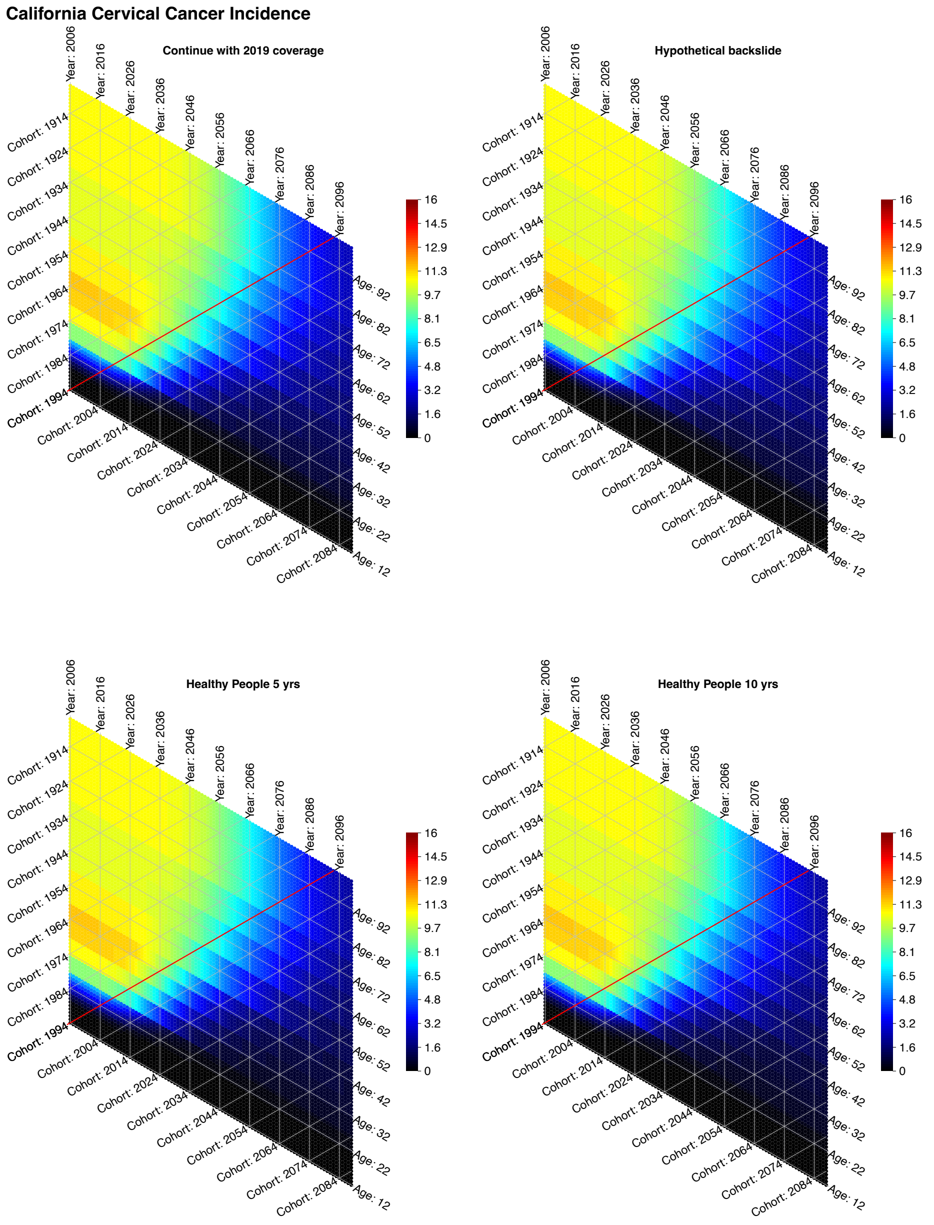

Supplementary Figure 17: Hexamaps visualizing cervical cancer incidence rate per 100,000 population in California as a function of age, calendar year, and birth year in the data. The vertical lines represent calendar years, the horizontal lines represent ages, and the diagonal lines represent the aging of each birth cohort with calendar years from the bottom left to the top right. In each scenario, the red diagonal line represents the birth cohort that became eligible to be vaccinated from the start of the simulation. Cohorts born after 1994 are represented by less heated colors at the bottom right of the red line.

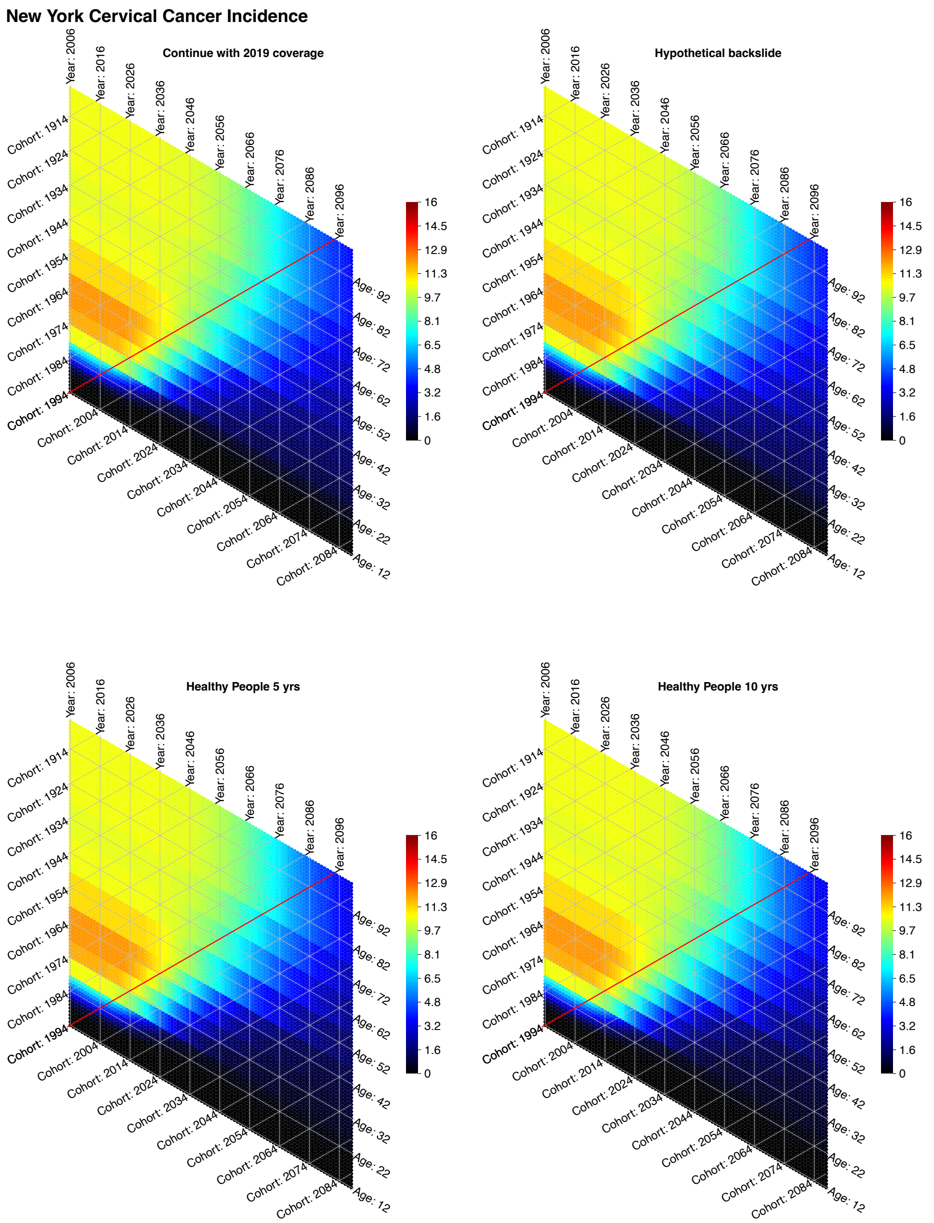

Supplementary Figure 18: Hexamaps visualizing cervical cancer incidence rate per 100,000 population in New York as a function of age, calendar year, and birth year in the data. The vertical lines represent calendar years, the horizontal lines represent ages, and the diagonal lines represent the aging of each birth cohort with calendar years from the bottom left to the top right. In each scenario, the red diagonal line represents the birth cohort that became eligible to be vaccinated from the start of the simulation. Cohorts born after 1994 are represented by less heated colors at the bottom right of the red line.

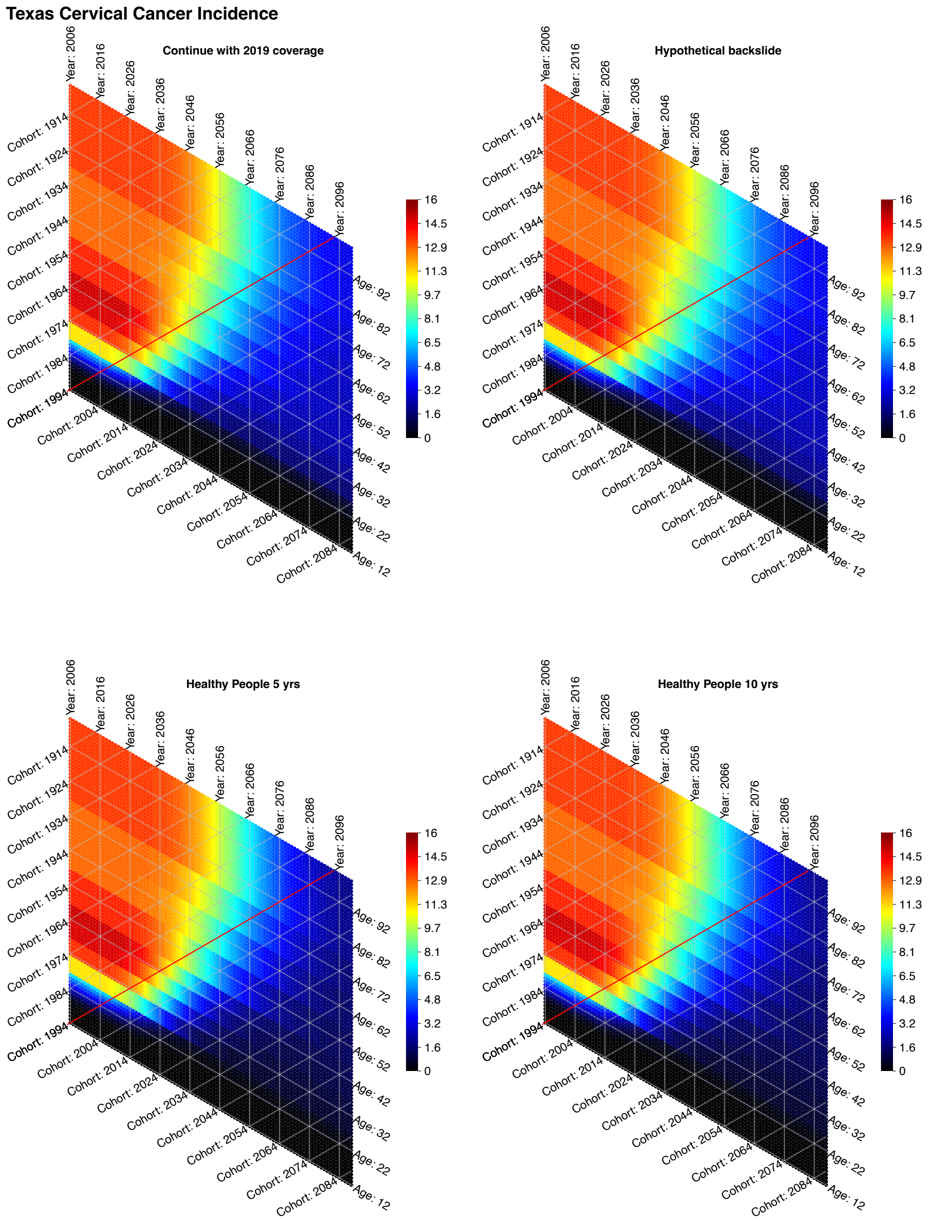

Supplementary Figure 19: Hexamaps visualizing cervical cancer incidence rate per 100,000 population in Texas as a function of age, calendar year, and birth year in the data. The vertical lines represent calendar years, the horizontal lines represent ages, and the diagonal lines represent the aging of each birth cohort with calendar years from the bottom left to the top right. In each scenario, the red diagonal line represents the birth cohort that became eligible to be vaccinated from the start of the simulation. Cohorts born after 1994 are represented by less heated colors at the bottom right of the red line.

#### 5.2 Sensitivity analysis on screening coverage

Supplementary Table 7: Sensitivity analysis on potential future screening coverage trends

| | | | | | |
|---|---|---|---|---|---|
| | | **Base case pap-based screening** | | | |
| | | US | CA | NY | TX |
| Elimination year | Status-quo | 2051 (2034-2064) | 2049 (2033-2061) | 2057 (2038-2071) | 2052 (2043-2061) |
| | backslide % change | year 2074 increase 3.06% (2.90-3.21) | year 2076 increase 3.42% (3.18-3.65) | year 2080 increase 2.76% (2.60-2.90) | year 2063 increase 4.06% (3.75-4.41) |
| Incidence | HI5yrs % change | year 2100 decrease 33.27% (28.88-38.57) | year 2100 decrease 24.98% (20.50-30.53) | year 2100 decrease 27.99% (24.61-32.26) | year 2100 decrease 51.21% (46.97-56.46) |
| | HI10yrs % change | year 2100 decrease 32.63% (28.26-37.92) | year 2100 decrease24.45% (20.04-29.96) | year 2100 decrease 27.28% (23.92-31.54) | year 2100 decrease 50.86% (46.63-56.07) |
| | backslide % change | year 2082 increase 3.02% (2.86-3.17) | year 2083 increase 3.41% (3.16-3.64) | year 2088 increase 2.75% (2.59-2.89) | year 2070 increase 3.90% (3.66-4.15) |
| Mortality | HI5yrs % change | year 2100 decrease29.78% (26.09-34.36) | year 2100 decrease 22.72% (19.08-27.35) | year 2100 decrease 23.88% (21.21-27.31) | year 2100 decrease 49.07% (45.08-54.07) |
| | HI10yrs % change | year 2100 decrease 28.98% (25.31-33.48) | year 2100 decrease 22.07% (18.49-26.65) | year 2100 decrease 23.04% (20.40-26.40) | year 2100 decrease 48.57% (44.60-53.55) |
| | | **Increase Pap: 50% never screeners start screening every 5 yrs** | | | |
| | | US | CA | NY | TX |
| Elimination year | Status-quo | 2040 (2029-2050) | 2039 (2028-2048) | 2043 (2031-2055) | 2046 (2039-2053) |
| | backslide % change | year 2068 increase 3.17% (2.96-3.45) | year 2072 increase 3.48% (3.23-3.72) | year 2075 increase 2.78% (2.61-2.94) | year 2061 increase 4.36% (3.99-4.78) |
| Incidence | HI5yrs % change | year 2100 decrease 34.31% (29.46-40.14) | year 2100 decrease 25.33% (20.31-31.53) | year 2100 decrease 29.33% (25.50-34.24) | year 2100 decrease 51.95% (47.53-57.35) |
| | HI10yrs % change | year 2100 decrease 33.76% (28.91-39.51) | year 2100 decrease24.86% (19.91-31.01) | year 2100 decrease 28.69% (24.86-33.59) | year 2100 decrease 51.67% (47.26-57.08) |
| | backslide % change | year 2079 increase 3.02% (2.86-3.17) | year 2080 increase 3.41% (3.16-3.63) | year 2086 increase 2.72% (2.56-2.86) | year 2068 increase 4.09% (3.81-4.39) |
| Mortality | HI5yrs % change | year 2100 decrease30.78% (26.69-35.69) | year 2100 decrease 23.15% (19.11-28.30) | year 2100 decrease 24.99% (21.93-28.94) | year 2100 decrease 49.92% (45.75-55.12) |
| | HI10yrs % change | year 2100 decrease 30.05% (25.97-34.88) | year 2100 decrease 22.55% (18.58-27.65) | year 2100 decrease 24.21% (21.16-28.13) | year 2100 decrease 49.50% (45.35-54.70) |
| | | **Decrease Pap: 50% women who screen every 5 yr become never screeners** | | | |
| | | US | CA | NY | TX |
| Elimination year | Status-quo | 2057 (2039-2068) | 2053 (2036-2064) | 2062 (2042-2076) | 2059 (2050-2068) |
| | backslide % change | year 2075 increase 3.29% (3.07-3.51) | year 2076 increase 3.64% (3.36-3.91) | year 2080 increase 2.97% (2.76-3.18) | year 2065 increase 4.10% (3.86-4.35) |
| Incidence | HI5yrs % change | year 2100 decrease 33.33% (29.20-38.33) | year 2100 decrease 25.15% (20.90-30.52) | year 2100 decrease 28.20% (24.93-32.28) | year 2100 decrease 50.69% (46.62-55.61) |
| | HI10yrs % change | year 2100 decrease 32.64% (28.52-37.58) | year 2100 decrease 24.59% (20.39-29.89) | year 2100 decrease 27.43% (24.19-31.48) | year 2100 decrease 50.27% (46.21-55.15) |
| | backslide % change | year 2082 increase 3.26% (3.05-3.47) | year 2082 decrease 3.63% (3.36-3.90) | year 2088 increase 2.96% (2.76-3.17) | year 2071 increase 4.02% (3.80-4.23) |
| Mortality | HI5yrs % change | year 2100 decrease 30.06% (26.57-34.41) | year 2100 decrease 22.96% (19.41-27.47) | year 2100 decrease 24.32% (21.64-27.63) | year 2100 decrease 48.59% (44.77-53.22) |
| | HI10yrs % change | year 2100 decrease 29.20% (25.73-33.48) | year 2100 decrease 22.28% (18.78-26.70) | year 2100 decrease 23.43% (20.80-26.69) | year 2100 decrease 48.01% (44.22-52.59) |
| | | **Iincrease HPV test: 50% never screeners start screening with HPV self-test every 5 yrs** | | | |
| | | US | CA | NY | TX |
| Elimination year | Status-quo | 2040 (2030-2050) | 2039 (2029-2048) | 2043 (2032-2054) | 2046 (2039-2052) |
| | backslide % change | year 2068 increase 3.14% (2.93-3.43) | year 2073 increase 3.45% (3.20-3.68) | year 2076 increase 2.76% (2.59-2.91) | year 2061 increase 4.34% (3.96-4.77) |
| Incidence | HI5yrs % change | year 2100 decrease 34.20% (29.37-39.97) | year 2100 decrease 25.25% (20.26-31.41) | year 2100 decrease 29.18% (25.37-34.07) | year 2100 decrease 51.91% (47.49-57.33) |
| | HI10yrs % change | year 2100 decrease 33.65% (28.82-39.34) | year 2100 decrease24.78% (19.86-30.90) | year 2100 decrease 28.53% (24.72-33.42) | year 2100 decrease 51.63% (47.22-57.06) |
| | backslide % change | year 2079 increase 3.01% (2.85-3.16) | year 2081 increase 3.39% (3.14-3.62) | year 2086 increase 2.71% (2.55-2.85) | year 2068 increase 4.07% (3.78-4.38) |
| Mortality | HI5yrs % change | year 2100 decrease30.66% (26.59-35.51) | year 2100 decrease 23.07% (19.07-28.17) | year 2100 decrease 24.84% (21.80-28.76) | year 2100 decrease 49.87% (45.70-55.07) |
| | HI10yrs % change | year 2100 decrease 29.93% (25.87-34.72) | year 2100 decrease 22.46% (18.53-27.52) | year 2100 decrease 24.05% (21.02-27.95) | year 2100 decrease 49.44% (45.30-54.66) |

### 6 Natural History Model Equations

$$\begin{aligned}\frac{dXXmw_{1l}}{dt} &= 0.5b\omega_l(1-\phi_{mil}) + \sigma VXmw_{1l} + \tau_{mv}ZXmw_{1l} + \tau_{mn}XZmw_{1l} \\ &+ (1-\nu_m)[\gamma_{mn}XYmw_{1l} + \gamma_{mv}(YXmw_{1l} + \alpha WXmw_{il})] \\ &- (\lambda_{mv} + \lambda_{mn} + d_1 + \mu_1)XXmw_{1l}\end{aligned} \tag{26}$$

$$\begin{aligned}\frac{dXXmw_{il}}{dt} &= \sigma VXmw_{il} + \tau_{mv}ZXmw_{il} + \tau_{mn}XZmw_{il} \\ &+ (1-\nu_m)[\gamma_{mn}XYmw_{il} + \gamma_{mv}(YXmw_{il} + \alpha WXmw_{il})] \\ &- (\lambda_{mv} + \lambda_{mn} + d_i + \mu_i + \phi_{mil})XXmw_{il} \\ &+ d_{i-1}XXmw_{(i-1)l}\end{aligned} \tag{27}$$

$$\begin{aligned}\frac{dXXfw_{1l}}{dt} &= 0.5b\omega_l(1-\phi_{fil}) + \sigma VXfw_{1l} + \tau_{fv}ZXfw_{1l} + \tau_{fn}XZfw_{1l} \\ &+ (1-\nu_f)[reg\_n_1XYfc2_{1l} \\ &+ \gamma_{fv}(YXfw_{1l} + \alpha WXfw_{1l}) + \gamma_{fn}XYfw_{1l} \\ &+ reg\_v_1(YXfc2_{1l} + WXfc2_{1l})] \\ &- (\lambda_{fv} + \lambda_{fn} + d_1 + \mu_1 + h_1)XXfw_{1l}\end{aligned} \tag{28}$$

$$\begin{aligned}\frac{dXXfw_{il}}{dt} &= \sigma VXfw_{il} + \tau_{fv}ZXfw_{il} + \tau_{fn}XZfw_{il} \\ &+ (1-\nu_f)[reg\_n_1XYfc2_{il} \\ &+ \gamma_{fv}(YXfw_{il} + \alpha WXfw_{il}) + \gamma_{fn}XYfw_{il} \\ &+ reg\_v_1(YXfc2_{il} + WXfc2_{il})] \\ &- (\lambda_{fv} + \lambda_{fn} + d_i + \mu_i + h_i + \phi_{fil})XXfw_{il} \\ &+ d_{i-1}XXfw_{(i-1)l}\end{aligned} \tag{29}$$

$$\begin{aligned}\frac{dXYmw_{1l}}{dt} &= \lambda_{mn}XXmw_{1l} + \tau_{mv}ZYmw_{1l} \\ &+ (1-\nu_m)\gamma_{mv}[YYmw_{1l} + \alpha WYmw_{1l}] \\ &- (\gamma_{mn} + d_1 + \mu_1 + \lambda_{mv})XYmw_{1l}\end{aligned} \tag{30}$$

$$\begin{aligned}\frac{dXYmw_{il}}{dt} &= \lambda_{mn}XXmw_{il} + \tau_{mv}ZYmw_{il} \\ &+ (1-\nu_m)\gamma_{mv}[YYmw_{il} + \alpha WYmw_{il}] \\ &- (\gamma_{mn} + d_i + \mu_i + \lambda_{mv} + \phi_{mil})XYmw_{il} \\ &+ d_{i-1}XYmw_{(i-1)l}\end{aligned} \tag{31}$$

$$\begin{aligned}\frac{dXYfw_{1l}}{dt} &= \lambda_{fn}XXfw_{1l} + +\tau_{fv}ZYfw_{1l} \\ &+ (1-\nu_f)[\gamma_{fv}(YYfw_{1l} + \alpha WYfw_{1l}) \\ &+ reg\_v_1(YYfc2_{1l} + WYfc2_{1l})] \\ &- (\gamma_{fn} + d_1 + \mu_1 + \lambda_{fv} + tr_{11} + tr_{12} + h_1)XYfw_{1l}\end{aligned} \tag{32}$$

$$\begin{aligned}\frac{dXYfw_{il}}{dt} &= \lambda_{fn}XXfw_{il} + \tau_{fv}ZYfw_{il} \\ &+ (1-\nu_f)[\gamma_{fv}(YYfw_{il} + \alpha WYfw_{il}) \\ &+ reg\_v_1(YYfc2_{il} + WYfc2_{il})] \\ &- (\gamma_{fn} + d_i + \mu_i + \lambda_{fv} + tr_{i1} + tr_{i2} + h_i + \phi_{fil})XYfw_{il} \\ &+ d_{i-1}XYfw_{(i-1)l}\end{aligned} \tag{33}$$

$$\begin{aligned}\frac{dXYfc2_{1l}}{dt} &= tr_{11}XYfw_{1l} + \tau_{fv}ZYfc2_{1l} + reg\_n_2XYfc3_{1l} \\ &- (d_1 + \mu_1 + tr_{13} + \lambda_{fv} + reg\_n_1 + h_1)XYfc2_{1l}\end{aligned} \tag{34}$$

$$\begin{aligned}\frac{dXYfc2_{il}}{dt} &= tr_{i1}XYfw_{il} + \tau_{fv}ZYfc2_{il} + reg\_n_2XYfc3_{il} \\ &- (d_i + \mu_i + tr_{i3} + \lambda_{fv} + reg\_n_1 + h_i + \phi_{fil})XYfc2_{il} \\ &+ d_{i-1}XYfc2_{(i-1)l}\end{aligned} \tag{35}$$

$$\begin{aligned}\frac{dXYfc3_{1l}}{dt} &= tr_{12}XYfw_{1l} + tr_{13}XYfc2_{1l} + \tau_{fv}ZYfc3_{1l} \\ &- (d_1 + \mu_1 + tr_{14} + \lambda_{fv} + reg\_n_2 + h_1)XYfc3_{1l}\end{aligned} \tag{36}$$

$$\begin{aligned}\frac{dXYfc3_{il}}{dt} &= tr_{i2}XYfw_{il} + tr_{i3}XYfc2_{il} + \tau_{fv}ZYfc3_{il} \\ &- (d_i + \mu_i + tr_{i4} + \lambda_{fv} + reg\_n_2 + h_i + \phi_{fil})XYfc3_{il} \\ &+ d_{i-1}XYfc3_{(i-1)l}\end{aligned} \tag{37}$$

$$\begin{aligned}\frac{dXYfpcl_{1l}}{dt} &= tr_{14}XYfc3_{1l} + \tau_{fv}ZYfpcl_{1l} \\ &- (d_1 + \mu_1 + canc\_prog_1 + detect_1)XYfpcl_{1l}\end{aligned} \tag{38}$$

$$\begin{aligned}\frac{dXYfpcl_{il}}{dt} &= tr_{i4}XYfc3_{il} + \tau_{fv}ZYfpcl_{il} \\ &- (d_i + \mu_i + canc\_prog_1 + detect_1)XYfpcl_{il} \\ &+ d_{i-1}XYfpcl_{(i-1)l}\end{aligned} \tag{39}$$

$$\begin{aligned}\frac{dXYfcl_{1l}}{dt} &= detect_1 XYpcl_{1l} \\ &- (d_1 + \mu_1 + canc\_surv_1 + canc\_death_1) XYfcl_{1l}\end{aligned} \tag{40}$$

$$\begin{aligned}\frac{dXYfcl_{il}}{dt} &= detect_1 XYpcl_{il} \\ &- (d_i + \mu_i + canc\_surv_1 + canc\_death_1) XYfcl_{il} \\ &+ d_{i-1} XYfcl_{(i-1)l}\end{aligned} \tag{41}$$

$$\begin{aligned}\frac{dXYfpcr_{1l}}{dt} &= canc\_prog_1 XYfpcl_{1l} + \tau_{fv} ZYfpcr_{1l} \\ &- (d_1 + \mu_1 + canc\_prog_2 + detect_2) XYfpcr_{1l}\end{aligned} \tag{42}$$

$$\begin{aligned}\frac{dXYfpcr_{il}}{dt} &= canc\_prog_1 XYfpcl_{il} + \tau_{fv} ZYfpcr_{il} \\ &- (d_i + \mu_i + canc\_prog_2 + detect_2) XYfpcr_{il} \\ &+ d_{i-1} XYfpcr_{(i-1)l}\end{aligned} \tag{43}$$

$$\begin{aligned}\frac{dXYfcr_{1l}}{dt} &= detect_2 XYpcr_{1l} \\ &- (d_1 + \mu_1 + canc\_surv_2 + canc\_death_2) XYfcr_{1l}\end{aligned} \tag{44}$$

$$\begin{aligned}\frac{dXYfcr_{il}}{dt} &= detect_2 XYpcr_{il} \\ &- (d_i + \mu_i + canc\_surv_2 + canc\_death_2) XYfcr_{il} \\ &+ d_{i-1} XYfcr_{(i-1)l}\end{aligned} \tag{45}$$

$$\begin{aligned}\frac{dXYfpcd_{1l}}{dt} &= canc\_prog_2 XYfpcr_{1l} + \tau_{fv} ZYfpcd_{1l} \\ &- (d_1 + \mu_1 + detect_3) XYfpcd_{1l}\end{aligned} \tag{46}$$

$$\begin{aligned}\frac{dXYfpcd_{il}}{dt} &= canc\_prog_2 XYfpcr_{il} + \tau_{fv} ZYfpcd_{il} \\ &- (d_i + \mu_i + detect_3) XYfpcd_{il} \\ &+ d_{i-1} XYfpcd_{(i-1)l}\end{aligned} \tag{47}$$

$$\begin{aligned}\frac{dXYfcd_{1l}}{dt} &= detect_3 XYpcd_{1l} \\ &- (d_1 + \mu_1 + canc\_surv_3 + canc\_death_3) XYfcd_{1l}\end{aligned} \tag{48}$$

$$
\begin{aligned}
\frac{dXYfcd_{il}}{dt} &= detect_3 XYpcd_{il} \\
&- (d_i + \mu_i + canc\_surv_3 + canc\_death_3) XYfcd_{il} \\
&+ d_{i-1} XYfcd_{(i-1)l}
\end{aligned} \tag{49}
$$

$$
\begin{aligned}
\frac{dXZmw_{1l}}{dt} &= \nu_m \gamma_{mn} XYmw_{1l} + \tau_{mv} ZZmw_{1l} \\
&+ (1 - \nu_m)[\gamma_{mv}(YZmw_{1l} + \alpha WZmw_{1l})] \\
&- (d_1 + \mu_1 + \lambda_{mv} + \tau_{mn}) XZmw_{1l}
\end{aligned} \tag{50}
$$

$$
\begin{aligned}
\frac{dXZmw_{il}}{dt} &= \nu_m \gamma_{mn} XYmw_{il} + \tau_{mv} ZZmw_{il} \\
&+ (1 - \nu_m)[\gamma_{mv}(YZmw_{il} + \alpha WZmw_{il})] \\
&- (d_i + \mu_i + \lambda_{mv} + \phi_{mil} + \tau_{mn}) XZmw_{il} \\
&+ d_{i-1} XZmw_{(i-1)l}
\end{aligned} \tag{51}
$$

$$
\begin{aligned}
\frac{dXZfw_{1l}}{dt} &= \nu_f (\gamma_{fn} XYfw_{1l} + reg\_n_1 XYfc2_{1l}) \\
&+ (1 - \nu_f)[\gamma_{fv}(YZfw_{1l} + \alpha WZfw_{1l}) \\
&+ reg\_v_1(YZfc2_{1l} + WZfc2_{1l})] \\
&+ \tau_{fv} ZZmw_{1l} \\
&- (d_1 + \mu_1 + \lambda_{fv} + \tau_{fn} + h_1) XZfw_{1l}
\end{aligned} \tag{52}
$$

$$
\begin{aligned}
\frac{dXZfw_{il}}{dt} &= \nu_f (\gamma_{fn} XYfw_{il} + reg\_n_1 XYfc2_{il}) \\
&+ (1 - \nu_f)[\gamma_{fv}(YZfw_{il} + \alpha WZfw_{il}) \\
&+ reg\_v_1(YZfc2_{il} + WZfc2_{il})] \\
&+ \tau_{fv} ZZmw_{il} \\
&- (d_i + \mu_i + \lambda_{fv} + \phi_{fil} + \tau_{fn} + h_i) XZfw_{il} \\
&+ d_{i-1} XZfw_{(i-1)l}
\end{aligned} \tag{53}
$$

$$
\begin{aligned}
\frac{dYXmw_{1l}}{dt} &= \lambda_{mv} XXmw_{1l} + (1 - \nu_m)\gamma_{mn} YYmw_{1l} + \tau_{mn} YZmw_{1l} \\
&- (d_1 + \mu_1 + \lambda_{mn} + \gamma_{mv}) YXmw_{1l}
\end{aligned} \tag{54}
$$

$$
\begin{aligned}
\frac{dYXmw_{il}}{dt} &= \lambda_{mv} XXmw_{il} + (1 - \nu_m)\gamma_{mn} YYmw_{il} + \tau_{mn} YZmw_{il} \\
&- (d_i + \mu_i + \lambda_{mn} + \gamma_{mv} + \upsilon \phi_{mil}) YXmw_{il} \\
&+ d_{i-1} YXmw_{(i-1)l}
\end{aligned} \tag{55}
$$

$$\begin{aligned}\frac{dYXfw_{1l}}{dt} &= \lambda_{fv}XXfw_{1l} + \tau_{fn}YZfw_{1l} \\ &+ (1-\nu_f)(\gamma_{fn}YYfw_{1l} + reg\_n_1YYfc2_{1l}) \\ &- [d_1 + \mu_1 + \lambda_{fn} + \gamma_{fv} + \upsilon\phi_{fil} + h_1 + HR_1(tr_{11} + tr_{12})]YXfw_{1l}\end{aligned} \tag{56}$$

$$\begin{aligned}\frac{dYXfw_{il}}{dt} &= \lambda_{fv}XXfw_{il} + \tau_{fn}YZfw_{il} \\ &+ (1-\nu_f)(\gamma_{fn}YYfw_{il} + reg\_n_1YYfc2_{il}) \\ &- [d_i + \mu_i + \lambda_{fn} + \gamma_{fv} + \upsilon\phi_{fil} + h_i + HR_1(tr_{i1} + tr_{i2})]YXfw_{il} \\ &+ d_{i-1}YXfw_{(i-1)l}\end{aligned} \tag{57}$$

$$\begin{aligned}\frac{dYXfc2_{1l}}{dt} &= HR_1tr_{i1}YXfw_{1l} + \tau_{fn}YZfc2_{1l} + reg\_v_2YXfc3_{1l} \\ &- (d_1 + \mu_1 + HR_2tr_{13} + \lambda_{fn} + reg\_v_1 + h_1)YXfc2_{il}\end{aligned} \tag{58}$$

$$\begin{aligned}\frac{dYXfc2_{il}}{dt} &= HR_1tr_{1i}YXfw_{il} + \tau_{fn}YZfc2_{il} + reg\_v_2YXfc3_{il} \\ &- (d_i + \mu_i + HR_2tr_{i3} + \lambda_{fn} + reg\_v_1 + h_i + \upsilon\phi_{fil})YXfc2_{il} \\ &+ d_{i-1}YXfc2_{(i-1)l}\end{aligned} \tag{59}$$

$$\begin{aligned}\frac{dYXfc3_{1l}}{dt} &= HR_1tr_{12}YXfw_{1l} + HR_2tr_{13}YXfc2_{1l} + \tau_{fn}YZfc3_{1l} \\ &- (d_1 + \mu_1 + HR_3tr_{14} + \lambda_{fn} + reg\_v_2 + h_1)YXfc3_{1l}\end{aligned} \tag{60}$$

$$\begin{aligned}\frac{dYXfc3_{il}}{dt} &= HR_1tr_{i2}YXfw_{il} + HR_2tr_{i3}YXfc2_{il} + \tau_{fn}YZfc3_{il} \\ &- (d_i + \mu_i + HR_3tr_{i4} + \lambda_{fn} + reg\_v_2 + h_i + \upsilon\phi_{fil})YXfc3_{il} \\ &+ d_{i-1}YXfc3_{(i-1)l}\end{aligned} \tag{61}$$

$$\begin{aligned}\frac{dYXfpcl_{1l}}{dt} &= HR_3tr_{14}YXfc3_{1l} + \tau_{fn}YZfpcl_{1l} \\ &- (d_1 + \mu_1 + canc\_prog_1 + detect_1)YXfpcl_{1l}\end{aligned} \tag{62}$$

$$\begin{aligned}\frac{dYXfpcl_{il}}{dt} &= HR_3tr_{i4}YXfc3_{il} + \tau_{fn}YZfpcl_{il} \\ &- (d_i + \mu_i + canc\_prog_1 + detect_1)YXfpcl_{il} \\ &+ d_{i-1}YXfpcl_{(i-1)l}\end{aligned} \tag{63}$$

$$\begin{aligned}\frac{dYXfcl_{1l}}{dt} &= detect_1 YXfpcl_{1l} \\ &- (d_i + \mu_i + canc\_surv_1 + canc\_death_1) YXfcl_{1l}\end{aligned} \tag{64}$$

$$\begin{aligned}\frac{dYXfcl_{il}}{dt} &= detect_1 YXfpcl_{il} \\ &- (d_i + \mu_i + canc\_surv_1 + canc\_death_1) YXfcl_{il} \\ &+ d_{i-1} YXfcl_{(i-1)l}\end{aligned} \tag{65}$$

$$\begin{aligned}\frac{dYXfpcr_{il}}{dt} &= canc\_prog_1 YXfpcl_{1l} + \tau_{fn} YZfpcr_{1l} \\ &- (d_1 + \mu_1 + canc\_prog_2 + detect_2) YXfpcr_{1l}\end{aligned} \tag{66}$$

$$\begin{aligned}\frac{dYXfpcr_{il}}{dt} &= canc\_prog_1 YXfpcl_{il} + \tau_{fn} YZfpcr_{il} \\ &- (d_i + \mu_i + canc\_prog_2 + detect_2) YXfpcr_{il} \\ &+ d_{i-1} YXfpcr_{(i-1)l}\end{aligned} \tag{67}$$

$$\begin{aligned}\frac{dYXfcr_{1l}}{dt} &= detect_2 YXpcr_{1l} \\ &- (d_1 + \mu_1 + canc\_surv_2 + canc\_death_2) YXfcr_{1l}\end{aligned} \tag{68}$$

$$\begin{aligned}\frac{dYXfcr_{il}}{dt} &= detect_2 YXpcr_{il} \\ &- (d_i + \mu_i + canc\_surv_2 + canc\_death_2) YXfcr_{il} \\ &+ d_{i-1} YXfcr_{(i-1)l}\end{aligned} \tag{69}$$

$$\begin{aligned}\frac{dYXfpcd_{1l}}{dt} &= canc\_prog_2 YXfpcr_{1l} + \tau_{fn} YZfpcd_{1l} \\ &- (d_1 + \mu_1 + detect_3) YXfpcd_{1l}\end{aligned} \tag{70}$$

$$\begin{aligned}\frac{dYXfpcd_{il}}{dt} &= canc\_prog_2 YXfpcr_{il} + \tau_{fn} YZfpcd_{il} \\ &- (d_i + \mu_i + detect_3) YXfpcd_{il} \\ &+ d_{i-1} YXfpcd_{(i-1)l}\end{aligned} \tag{71}$$

$$\begin{aligned}\frac{dYXfcd_{1l}}{dt} &= detect_3 YXpcd_{1l} \\ &- (d_1 + \mu_1 + canc\_surv_3 + canc\_death_3) YXfcd_{1l}\end{aligned} \tag{72}$$

$$\frac{dYXfcd_{il}}{dt} = detect_3 YXpcd_{il} - (d_i + \mu_i + canc\_surv_3 + canc\_death_3) YXfcd_{il} + d_{i-1} YXfcd_{(i-1)l} \tag{73}$$

$$\frac{dYYmw_{1l}}{dt} = \lambda_{mv} XYmw_{1l} + \lambda_{mn} YXmw_{1l} - (d_1 + \mu_1 + \gamma_{mv} + \gamma_{mn}) YYmw_{1l} \tag{74}$$

$$\frac{dYYmw_{il}}{dt} = \lambda_{mv} XYmw_{il} + \lambda_{mn} YXmw_{il} - (d_i + \mu_i + \gamma_{mv} + \gamma_{mn} + \upsilon\phi_{mil}) YYmw_{il} + d_{i-1} YYmw_{(i-1)l} \tag{75}$$

$$\frac{dYYfw_{1l}}{dt} = \lambda_{fv} XYfw_{1l} + \lambda_{fn} YXfw_{1l} - [d_1 + \mu_1 + \gamma_{fv} + \gamma_{fn} + h_1 + (1 + HR_1)tr_{11} + (1 + HR_1)tr_{12}] YYfw_{1l} \tag{76}$$

$$\frac{dYYfw_{il}}{dt} = \lambda_{fv} XYfw_{il} + \lambda_{fn} YXfw_{il} - [d_i + \mu_i + \gamma_{fv} + \gamma_{fn} + \upsilon\phi_{fil} + h_i + (1 + HR_1)tr_{i1} + (1 + HR_1)tr_{i2}] YYfw_{il} + d_{i-1} YYfw_{(i-1)l} \tag{77}$$

$$\frac{dYYfc2_{1l}}{dt} = (1 + HR_1)tr_{11} YYfw_{1l} + (reg\_n_2 + reg\_v_2) YYfc3_{1l} + \lambda_{fv} XYfc2_{1l} + \lambda_{fn} YXfc2_{1l} - [d_1 + \mu_1 + (1 + HR_2)tr_{13} + reg\_v_1 + reg\_n_1 + h_1] YYfc2_{1l} \tag{78}$$

$$\frac{dYYfc2_{il}}{dt} = (1 + HR_1)tr_{i1} YYfw_{il} + (reg\_n_2 + reg\_v_2) YYfc3_{il} + \lambda_{fv} XYfc2_{il} + \lambda_{fn} YXfc2_{il} - [d_i + \mu_i + (1 + HR_2)tr_{i3} + reg\_v_1 + reg\_n_1 + h_i + \upsilon\phi_{fil}] YYfc2_{il} + d_{i-1} YYfc2_{(i-1)l} \tag{79}$$

$$\frac{dYYfc3_{1l}}{dt} = (1+HR_1)tr_{12}YYfw_{1l} + (1+HR_2)tr_{13}YYfc2_{1l} + \lambda_{fv}XYfc3_{1l} + \lambda_{fn}YXfc3_{1l} - [d_1 + \mu_1 + (1+HR_3)tr_{14} + reg\_v_2 + reg\_n_2 + h_1]YYfc3_{1l} \tag{80}$$

$$\frac{dYYfc3_{il}}{dt} = (1+HR_1)tr_{i2}YYfw_{il} + (1+HR_2)tr_{i3}YYfc2_{il} + \lambda_{fv}XYfc3_{il} + \lambda_{fn}YXfc3_{il} - [d_i + \mu_i + (1+HR_3)tr_{i4} + reg\_v_2 + reg\_n_2 + h_i + v\phi_{fil}]YYfc3_{il} + d_{i-1}YYfc3_{(i-1)l} \tag{81}$$

$$\frac{dYYfpcl_{1l}}{dt} = (1+HR_3)tr_{14}YYfc3_{1l} - (d_1 + \mu_1 + canc\_prog_1 + detect_1)YYfpcl_{1l} \tag{82}$$

$$\frac{dYYfpcl_{il}}{dt} = (1+HR_3)tr_{i4}YYfc3_{il} - (d_i + \mu_i + canc\_prog_1 + detect_1)YYfpcl_{il} + d_{i-1}YYfpcl_{(i-1)l} \tag{83}$$

$$\frac{dYYfcl_{1l}}{dt} = detect_1 YYpcl_{1l} - (d_1 + \mu_1 + canc\_surv_1 + canc\_death_1)YYfcl_{1l} \tag{84}$$

$$\frac{dYYfcl_{il}}{dt} = detect_1 YYpcl_{il} - (d_i + \mu_i + canc\_surv_1 + canc\_death_1)YYfcl_{il} + d_{i-1}YYfcl_{(i-1)l} \tag{85}$$

$$\frac{dYYfpcr_{1l}}{dt} = canc\_prog_1 YYfpcl_{1l} - (d_1 + \mu_1 + canc\_prog_2 + detect_2)YYfpcr_{1l} \tag{86}$$

$$\frac{dYYfpcr_{il}}{dt} = canc\_prog_1 YYfpcl_{il} - (d_i + \mu_i + canc\_prog_2 + detect_2)YYfpcr_{il} + d_{i-1}YYfpcr_{(i-1)l} \tag{87}$$

$$\frac{dYYfcr_{1l}}{dt} = detect_2 YYpcr_{il} - (d_1 + \mu_1 + canc\_surv_2 + canc\_death_2) YYfcr_{1l} \tag{88}$$

$$\frac{dYYfcr_{il}}{dt} = detect_2 YYpcr_{il} - (d_i + \mu_i + canc\_surv_2 + canc\_death_2) YYfcr_{il} + d_{i-1} YYfcr_{(i-1)l} \tag{89}$$

$$\frac{dYYfpcd_{1l}}{dt} = canc\_prog_2 YYfpcr_{1l} - (d_1 + \mu_1 + detect_3) YYfpcd_{1l} \tag{90}$$

$$\frac{dYYfpcd_{il}}{dt} = canc\_prog_2 YYfpcr_{il} - (d_i + \mu_i + detect_3) YYfpcd_{il} + d_{i-1} YYfpcd_{(i-1)l} \tag{91}$$

$$\frac{dYYfcd_{1l}}{dt} = detect_3 YYpcd_{1l} - (d_1 + \mu_1 + canc\_surv_3 + canc\_death_3) YYfcd_{1l} \tag{92}$$

$$\frac{dYYfcd_{il}}{dt} = detect_3 YYpcd_{il} - (d_i + \mu_i + canc\_surv_3 + canc\_death_3) YYfcd_{il} + d_{i-1} YYfcd_{(i-1)l} \tag{93}$$

$$\frac{dYZmw_{1l}}{dt} = \lambda_{mv} XZmw_{1l} + \nu_m \gamma_{mn} YYmw_{1l} - (d_1 + \mu_1 + \gamma_{mv} + \tau_{mn}) YZmw_{1l} \tag{94}$$

$$\frac{dYZmw_{il}}{dt} = \lambda_{mv} XZmw_{il} + \nu_m \gamma_{mn} YYmw_{il} - (d_i + \mu_i + \gamma_{mv} + \tau_{mn} + \upsilon \phi_{mil}) YZmw_{il} + d_{i-1} YZmw_{(i-1)l} \tag{95}$$

$$\frac{dYZfw_{1l}}{dt} = \lambda_{fv} XYfw_{1l} + \nu_f (\gamma_{fn} YYfw_{1l} + reg\_n_1 YYfc2_{1l}) - (d_1 + \mu_1 + h_1 + HR_1 tr_{11} + HR_1 tr_{12} + \gamma_{fv} + \tau_{fn}) YZfw_{1l} \tag{96}$$

$$\frac{dYZfw_{il}}{dt} = \lambda_{fv}XYfw_{il} + \nu_f(\gamma_{fn}YYfw_{il} + reg\_n_1YYfc2_{il}) \\ - (d_i + \mu_i + h_i + HR_1tr_{i1} + HR_1tr_{i2} + \gamma_{fv} + \tau_{fn} + \upsilon\phi_{fil})YZfw_{il} \\ + d_{i-1}YZfw_{(i-1)l} \tag{97}$$

$$\frac{dYZfc2_{1l}}{dt} = HR_1tr_{11}YZfw_{1l} + reg\_v_2YZfc3_{1l} \\ - (d_1 + \mu_1 + HR_2tr_{13} + reg\_v + h_1 + \tau_{fn})YZfc2_{1l} \tag{98}$$

$$\frac{dYZfc2_{il}}{dt} = HR_1tr_{i1}YZfw_{il} + reg\_v_2YZfc3_{il} \\ - (d_i + \mu_i + HR_2tr_{i3} + reg\_v + h_i + \tau_{fn} + \upsilon\phi_{fil})YZfc2_{il} \\ + d_{i-1}YZfc2_{(i-1)l} \tag{99}$$

$$\frac{dYZfc3_{1l}}{dt} = HR_1tr_{12}YZfw_{1l} + HR_2tr_{13}YZfc2_{1l} \\ - (d_1 + \mu_1 + HR_3tr_{14} + reg\_v_2 + h_1 + \tau_{fn})YZfc3_{1l} \tag{100}$$

$$\frac{dYZfc3_{il}}{dt} = HR_1tr_{i2}YZfw_{il} + HR_2tr_{i3}YZfc2_{il} \\ - (d_i + \mu_i + HR_3tr_{i4} + reg\_v_2 + h_i + \tau_{fn} + \upsilon\phi_{fil})YZfc3_{il} \\ + d_{i-1}YZfc3_{(i-1)l} \tag{101}$$

$$\frac{dYZfpcl_{1l}}{dt} = HR_3tr_{14}YZfc3_{1l} \\ - (d_1 + \mu_1 + canc\_prog_1 + detect_1 + \tau_{fn})YZfpcl_{1l} \tag{102}$$

$$\frac{dYZfpcl_{il}}{dt} = HR_3tr_{i4}YZfc3_{il} \\ - (d_i + \mu_i + canc\_prog_1 + detect_1 + \tau_{fn})YZfpcl_{il} \\ + d_{i-1}YZfpcl_{(i-1)l} \tag{103}$$

$$\frac{dYZfcl_{1l}}{dt} = detect_1YZpcl_l \\ - (d_1 + \mu_1 + canc\_surv_1 + canc\_death_1)YZfcl_l \tag{104}$$

$$\frac{dYZfcl_{il}}{dt} = detect_1 YZpcl_{il} - (d_i + \mu_i + canc\_surv_1 + canc\_death_1) YZfcl_{il} + d_{i-1} YZfcl_{(i-1)l} \tag{105}$$

$$\frac{dYZfpcr_{1l}}{dt} = canc\_prog_1 YZfpcl_{1l} - (d_1 + \mu_1 + canc\_prog_2 + detect_2 + \tau_{fn}) YZfpcr_{1l} \tag{106}$$

$$\frac{dYZfpcr_{il}}{dt} = canc\_prog_1 YZfpcl_{il} - (d_i + \mu_i + canc\_prog_2 + detect_2 + \tau_{fn}) YZfpcr_{il} + d_{i-1} YZfpcr_{(i-1)l} \tag{107}$$

$$\frac{dYZfcr_{1l}}{dt} = detect_2 YZpcr_{1l} - (d_1 + \mu_1 + canc\_surv_2 + canc\_death_2) YZfcr_{1l} \tag{108}$$

$$\frac{dYZfcr_{il}}{dt} = detect_2 YZpcr_{il} - (d_i + \mu_i + canc\_surv_2 + canc\_death_2) YZfcr_{il} + d_{i-1} YZfcr_{(i-1)l} \tag{109}$$

$$\frac{dYZfpcd_{1l}}{dt} = canc\_prog_2 YZfpcr_{1l} - (d_1 + \mu_1 + detect_3 + \tau_{fn}) YZfpcd_{1l} \tag{110}$$

$$\frac{dYZfpcd_{il}}{dt} = canc\_prog_2 YZfpcr_{il} - (d_i + \mu_i + detect_3 + \tau_{fn}) YZfpcd_{il} + d_{i-1} YZfpcd_{(i-1)l} \tag{111}$$

$$\frac{dYZfcd_{1l}}{dt} = detect_3 YZpcd_{1l} - (d_1 + \mu_1 + canc\_surv_3 + canc\_death_3) YZfcd_{1l} \tag{112}$$

$$\begin{aligned}\frac{dYZfcd_{il}}{dt} &= detect_3 YZpcd_{il} \\ &- (d_i + \mu_i + canc\_surv_3 + canc\_death_3)YZfcd_{il} \\ &+ d_{i-1}YZfcd_{(i-1)l}\end{aligned} \tag{113}$$

$$\begin{aligned}\frac{dVXmw_{1l}}{dt} &= 0.5b\omega_l\phi_{m1l} + (1-\nu_m)\gamma_{mn}VYmw_{1l} + \tau_{mn}VZmw_{1l} \\ &- (d_1 + \mu_1 + \delta_m\lambda_{mv} + \lambda_{mn} + \sigma)VXmw_{1l}\end{aligned} \tag{114}$$

$$\begin{aligned}\frac{dVXmw_{il}}{dt} &= \phi_{mil}XXmw_{il} + (1-\nu_m)\gamma_{mn}VYmw_{il} + \tau_{mn}VZmw_{il} \\ &- (d_i + \mu_i + \delta_m\lambda_{mv} + \lambda_{mn} + \sigma)VXmw_{il} \\ &+ d_{i-1}VXmw_{(i-1)l}\end{aligned} \tag{115}$$

$$\begin{aligned}\frac{dVXfw_{1l}}{dt} &= 0.5b\omega_l\phi_{f1l} + \tau_{fn}VZfw_{1l} \\ &+ (1-\nu_f)(\gamma_{fn}VYfw_{1l} + reg\_n_1VYfc2_{1l}) \\ &- (\delta_f\lambda_{fv} + \lambda_{fn} + d_1 + \mu_1 + h_1 + \sigma)VXfw_{1l}\end{aligned} \tag{116}$$

$$\begin{aligned}\frac{dVXfw_{il}}{dt} &= \phi_{fil}XXfw_{il} + \tau_{fn}VZfw_{il} \\ &+ (1-\nu_f)(\gamma_{fn}VYfw_{il} + reg\_n_1VYfc2_{il}) \\ &- (\delta_f\lambda_{fv} + \lambda_{fn} + d_i + \mu_i + h_i + \sigma)VXfw_{il} \\ &+ d_{i-1}VXfw_{(i-1)l}\end{aligned} \tag{117}$$

$$\begin{aligned}\frac{dVYmw_{1l}}{dt} &= \lambda_{mn}VXmw_{1l} \\ &- (\gamma_{mn} + d_1 + \mu_1 + \delta_m\lambda_{mv})VYmw_{1l}\end{aligned} \tag{118}$$

$$\begin{aligned}\frac{dVYmw_{il}}{dt} &= \lambda_{mn}VXmw_{il} + \phi_{mil}XYmw_{il} \\ &- (\gamma_{mn} + d_i + \mu_i + \delta_m\lambda_{mv})VYmw_{il} \\ &+ d_{i-1}VYmw_{(i-1)l}\end{aligned} \tag{119}$$

$$\begin{aligned}\frac{dVYfw_{il}}{dt} &= \lambda_{fn}VXfw_{1l} \\ &- (\gamma_{fn} + d_1 + \mu_1 + h_1 + \delta_f\lambda_{fv} + tr_{11} + tr_{12})VYfw_{1l}\end{aligned} \tag{120}$$

$$\frac{dVYfw_{il}}{dt} = \lambda_{fn}VXfw_{il} + \phi_{fil}XYfw_{il} - (\gamma_{fn} + d_i + \mu_i + h_i + \delta_f\lambda_{fv} + tr_{i1} + tr_{i2})VYfw_{il} + d_{i-1}VYfw_{(i-1)l} \tag{121}$$

$$\frac{dVYfc2_{1l}}{dt} = tr_{11}VYfw_{1l} + reg\_n_2VYfc3_{1l} - (d_1 + \mu_1 + tr_{13} + reg\_n_1 + h_1 + \delta_f\lambda_{fv})VYfc2_{1l} \tag{122}$$

$$\frac{dVYfc2_{il}}{dt} = tr_{i1}VYfw_{il} + \phi_{fil}XYfc2_{il} + reg\_n_2VYfc3_{il} - (d_i + \mu_i + tr_{i3} + reg\_n_1 + h_i + \delta_f\lambda_{fv})VYfc2_{il} + d_{i-1}VYfc2_{(i-1)l} \tag{123}$$

$$\frac{dVYfc3_{1l}}{dt} = tr_{12}VYfw_{1l} + tr_{13}VYfc2_{1l} - (d_1 + \mu_1 + tr_{14} + \delta_f\lambda_{fv} + reg\_n_2 + h_1)VYfc3_{1l} \tag{124}$$

$$\frac{dVYfc3_{il}}{dt} = tr_{i2}VYfw_{il} + tr_{i3}VYfc2_{il} + \phi_{fil}XYfc3_{il} - (d_i + \mu_i + tr_{i4} + \delta_f\lambda_{fv} + reg\_n_2 + h_i)VYfc3_{il} + d_{i-1}VYfc3_{(i-1)l} \tag{125}$$

$$\frac{dVYfpcl_{1l}}{dt} = tr_{14}VYfc3_{1l} - (d_1 + \mu_1 + canc\_prog_1 + detect_1)VYfpcl_{1l} \tag{126}$$

$$\frac{dVYfpcl_{il}}{dt} = tr_{i4}VYfc3_{il} - (d_i + \mu_i + canc\_prog_1 + detect_1)VYfpcl_{il} + d_{i-1}VYfpcl_{(i-1)l} \tag{127}$$

$$\frac{dVYfcl_{1l}}{dt} = detect_1VYpcl_{1l} - (d_1 + \mu_1 + canc\_surv_1 + canc\_death_1)VYfcl_{1l} \tag{128}$$

$$\frac{dVYfcl_{il}}{dt} = detect_1 VYpcl_{il} - (d_i + \mu_i + canc\_surv_1 + canc\_death_1) VYfcl_{il} + d_{i-1} VYfcl_{(i-1)l} \tag{129}$$

$$\frac{dVYfpcr_{1l}}{dt} = canc\_prog_1 VYfpcl_{1l} - (d_1 + \mu_1 + canc\_prog_2 + detect_2) VYfpcr_{1l} \tag{130}$$

$$\frac{dVYfpcr_{il}}{dt} = canc\_prog_1 VYfpcl_{il} - (d_i + \mu_i + canc\_prog_2 + detect_2) VYfpcr_{il} + d_{i-1} VYfpcr_{(i-1)l} \tag{131}$$

$$\frac{dVYfcr_{1l}}{dt} = detect_2 VYpcr_{1l} - (d_1 + \mu_1 + canc\_surv_2 + canc\_death_2) VYfcr_{1l} \tag{132}$$

$$\frac{dVYfcr_{il}}{dt} = detect_2 VYpcr_{il} - (d_i + \mu_i + canc\_surv_2 + canc\_death_2) VYfcr_{il} + d_{i-1} VYfcr_{(i-1)l} \tag{133}$$

$$\frac{dVYfpcd_{il}}{dt} = canc\_prog_2 VYfpcr_{1l} - (d_1 + \mu_1 + detect_3) VYfpcd_{1l} \tag{134}$$

$$\frac{dVYfpcd_{il}}{dt} = canc\_prog_2 VYfpcr_{il} - (d_i + \mu_i + detect_3) VYfpcd_{il} + d_{i-1} VYfpcd_{(i-1)l} \tag{135}$$

$$\frac{dVYfcd_{1l}}{dt} = detect_3 VYfpcd_{1l} - (d_1 + \mu_1 + canc\_surv_3 + canc\_death_3) VYfcd_{1l} \tag{136}$$

$$\frac{dVYfcd_{il}}{dt} = detect_3 VYfpcd_{il} - (d_i + \mu_i + canc\_surv_3 + canc\_death_3)VYfcd_{il} + d_{i-1}VYfcd_{(i-1)l} \tag{137}$$

$$\frac{dVZmw_{il}}{dt} = \nu_m\gamma_{mn}VYmw_{1l} - (d_1 + \mu_1 + \delta_m\lambda_{mv} + \tau_{mn})VZmw_{1l} \tag{138}$$

$$\frac{dVZmw_{il}}{dt} = \nu_m\gamma_{mn}VYmw_{il} + \phi_{mil}XZmw_{il} - (d_i + \mu_i + \delta_m\lambda_{mv} + \tau_{mn})VZmw_{il} + d_{i-1}VZmw_{(i-1)l} \tag{139}$$

$$\frac{dVZfw_{1l}}{dt} = \nu_f(\gamma_{fn}VYfw_{1l} + reg\_n_1VYfc2_{1l}) - (d_1 + \mu_1 + \delta_f\lambda_{fv} + \tau_{fn} + h_1)VZfw_{il} \tag{140}$$

$$\frac{dVZfw_{il}}{dt} = \nu_f(\gamma_{fn}VYfw_{il} + reg\_n_1VYfc2_{il}) + \phi_{fil}XZfw_{il} - (d_i + \mu_i + \delta_f\lambda_{fv} + \tau_{fn} + h_i)VZfw_{il} + d_{i-1}VZfw_{(i-1)l} \tag{141}$$

$$\frac{dWXmw_{il}}{dt} = \delta_m\lambda_{mv}VXmw_{1l} + (1-\nu_m)\gamma_{mn}WYmw_{1l} + \tau_{mn}WZmw_{1l} - (d_1 + \mu_1 + \lambda_{mn} + \alpha\gamma_{mv})WXmw_{1l} \tag{142}$$

$$\frac{dWXmw_{il}}{dt} = \upsilon\phi_{mil}YXmw_{il} + \delta_m\lambda_{mv}VXmw_{il} + (1-\nu_m)\gamma_{mn}WYmw_{il} + \tau_{mn}WZmw_{il} - (d_i + \mu_i + \lambda_{mn} + \alpha\gamma_{mv})WXmw_{il} + d_{i-1}WXmw_{(i-1)l} \tag{143}$$

$$\frac{dWXfw_{1l}}{dt} = \delta_f\lambda_{fv}VXfw_{1l} + \tau_{fn}WZfw_{1l} + (1-\nu_f)[\gamma_{fn}WYfw_{1l} + reg\_n_1WYfc2_{1l}] - [d_1 + \mu_1 + h_1 + \lambda_{fn} + \alpha\gamma_{fv} + HR_1(tr_{11} + tr_{12})]WXfw_{1l} \tag{144}$$

$$\begin{aligned}\frac{dWXfw_{il}}{dt} &= \upsilon\phi_{fil}YXfw_{il} + \delta_f\lambda_{fv}VXfw_{il} + \tau_{fn}WZfw_{il} \\ &+ (1-\nu_f)[\gamma_{fn}WYfw_{il} + reg\_n_1WYfc2_{il}] \\ &- [d_i + \mu_i + h_i + \lambda_{fn} + \alpha\gamma_{fv} + HR_1(tr_{i1} + tr_{i2})]WXfw_{il} \\ &+ d_{i-1}WXfw_{(i-1)l}\end{aligned} \tag{145}$$

$$\begin{aligned}\frac{dWXfc2_{1l}}{dt} &= HR_1tr_{11}WXfw_{1l} + \tau_{fn}WZfc2_{1l} + reg\_v_2WXfc3_{1l} \\ &- (d_1 + \mu_1 + HR_2tr_{13} + \lambda_{fn} + reg\_v_1 + h_1)WXfc2_{1l}\end{aligned} \tag{146}$$

$$\begin{aligned}\frac{dWXfc2_{il}}{dt} &= \upsilon\phi_{fil}YXfc2_{il} + HR_1tr_{i1}WXfw_{il} \\ &+ \tau_{fn}WZfc2_{il} + reg\_v_2WXfc3_{il} \\ &- (d_i + \mu_i + HR_2tr_{i3} + \lambda_{fn} + reg\_v_1 + h_i)WXfc2_{il} \\ &+ d_{i-1}WXfc2_{(i-1)l}\end{aligned} \tag{147}$$

$$\begin{aligned}\frac{dWXfc3_{1l}}{dt} &= HR_1tr_{12}WXfw_{1l} + HR_2tr_{13}WXfc2_{1l} + \tau_{fn}WZfc3_{1l} \\ &- (d_1 + \mu_1 + HR_3tr_{14} + \lambda_{fn} + reg\_v_2 + h_1)WXfc3_{1l}\end{aligned} \tag{148}$$

$$\begin{aligned}\frac{dWXfc3_{il}}{dt} &= \upsilon\phi_{fil}YXfc3_{il} + HR_1tr_{i2}WXfw_{il} + HR_2tr_{i3}WXfc2_{il} + \tau_{fn}WZfc3_{il} \\ &- (d_i + \mu_i + HR_3tr_{i4} + \lambda_{fn} + reg\_v_2 + h_i)WXfc3_{il} \\ &+ d_{i-1}WXfc3_{(i-1)l}\end{aligned} \tag{149}$$

$$\begin{aligned}\frac{dWXfpcl_{1l}}{dt} &= HR_3tr_{14}WXfc3_{1l} + \tau_{fn}WZfpcl_{1l} \\ &- (d_1 + \mu_1 + canc\_prog_1 + detect_1)WXfpcl_{1l}\end{aligned} \tag{150}$$

$$\begin{aligned}\frac{dWXfpcl_{il}}{dt} &= HR_3tr_{i4}WXfc3_{il} + \tau_{fn}WZfpcl_{il} \\ &- (d_i + \mu_i + canc\_prog_1 + detect_1)WXfpcl_{il} \\ &+ d_{i-1}WXfpcl_{(i-1)l}\end{aligned} \tag{151}$$

$$\begin{aligned}\frac{dWXfcl_{1l}}{dt} &= detect_1WXpcl_{1l} \\ &- (d_1 + \mu_1 + canc\_surv_1 + canc\_death_1)WXfcl_{1l}\end{aligned} \tag{152}$$

$$\frac{dWXfcl_{il}}{dt} = detect_1 WXpcl_{il} - (d_i + \mu_i + canc\_surv_1 + canc\_death_1)WXfcl_{il} + d_{i-1}WXfcl_{(i-1)l} \tag{153}$$

$$\frac{dWXfpcr_{1l}}{dt} = canc\_prog_1 WXfpcl_{1l} + \tau_{fn}WZfpcr_{1l} - (d_1 + \mu_1 + canc\_prog_2 + detect_2)WXfpcr_{1l} \tag{154}$$

$$\frac{dWXfpcr_{il}}{dt} = canc\_prog_1 WXfpcl_{il} + \tau_{fn}WZfpcr_{il} - (d_i + \mu_i + canc\_prog_2 + detect_2)WXfpcr_{il} + d_{i-1}WXfpcr_{(i-1)l} \tag{155}$$

$$\frac{dWXfcr_{1l}}{dt} = detect_2 WXpcr_{1l} - (d_1 + \mu_1 + canc\_surv_2 + canc\_death_2)WXfcr_{1l} \tag{156}$$

$$\frac{dWXfcr_{il}}{dt} = detect_2 WXpcr_{il} - (d_i + \mu_i + canc\_surv_2 + canc\_death_2)WXfcr_{il} + d_{i-1}WXfcr_{(i-1)l} \tag{157}$$

$$\frac{dWXfpcd_{1l}}{dt} = canc\_prog_2 WXfpcr_{1l} + \tau_{fn}WZfpcd_{1l} - (d_1 + \mu_1 + detect_3)WXfpcd_{1l} \tag{158}$$

$$\frac{dWXfpcd_{il}}{dt} = canc\_prog_2 WXfpcr_{il} + \tau_{fn}WZfpcd_{il} - (d_i + \mu_i + detect_3)WXfpcd_{il} + d_{i-1}WXfpcd_{(i-1)l} \tag{159}$$

$$\frac{dWXfcd_{1l}}{dt} = detect_3 WXpcd_{1l} - (d_1 + \mu_1 + canc\_surv_3 + canc\_death_3)WXfcd_{1l} \tag{160}$$

$$\begin{aligned}\frac{dWXfcd_{il}}{dt} &= detect_3 WXpcd_{il} \\ &- (d_i + \mu_i + canc\_surv_3 + canc\_death_3)WXfcd_{il} \\ &+ d_{i-1}WXfcd_{(i-1)l}\end{aligned} \tag{161}$$

$$\begin{aligned}\frac{dWYmw_{1l}}{dt} &= \delta_m \lambda_{mv} VYmw_{1l} + \lambda_{mn} WXmw_{il} \\ &- (d_1 + \mu_1 + \alpha\gamma_{mv} + \gamma_{mn})WYmw_{1l}\end{aligned} \tag{162}$$

$$\begin{aligned}\frac{dWYmw_{il}}{dt} &= \upsilon\phi_{mil} YYmw_{il} \\ &+ \delta_m \lambda_{mv} VYmw_{il} + \lambda_{mn} WXmw_{il} \\ &- (d_i + \mu_i + \alpha\gamma_{mv} + \gamma_{mn})WYmw_{il} \\ &+ d_{i-1}WYmw_{(i-1)l}\end{aligned} \tag{163}$$

$$\begin{aligned}\frac{dWYfw_{1l}}{dt} &= \delta_f \lambda_{fv} VYfw_{1l} + \lambda_{fn} WXfw_{1l} \\ &- [d_1 + \mu_1 + h_1 + \alpha\gamma_{fv} + \gamma_{fn} + (1 + HR_1)(tr_{11} + tr_{12})]WYfw_{1l}\end{aligned} \tag{164}$$

$$\begin{aligned}\frac{dWYfw_{il}}{dt} &= \upsilon\phi_{fil} YYfw_{il} \\ &+ \delta_f \lambda_{fv} VYfw_{il} + \lambda_{fn} WXfw_{il} \\ &- [d_i + \mu_i + h_i + \alpha\gamma_{fv} + \gamma_{fn} + (1 + HR_1)(tr_{i1} + tr_{i2})]WYfw_{il} \\ &+ d_{i-1}WYfw_{(i-1)l}\end{aligned} \tag{165}$$

$$\begin{aligned}\frac{dWYfc2_{1l}}{dt} &= (HR_1 + 1)tr_{11} WYfw_{1l} + reg\_n_2 WYfc3_{1l} \\ &+ \delta_f \lambda_{fv} VYfc2_{1l} + \lambda_{fn} WXfc2_{1l} \\ &- [d_1 + \mu_1 + (1 + HR_2)tr_{13} + reg\_v_1 + reg\_n_1]WYfc2_{1l}\end{aligned} \tag{166}$$

$$\begin{aligned}\frac{dWYfc2_{il}}{dt} &= \upsilon\phi_{fil} YYfc2_{il} \\ &+ (HR_1 + 1)tr_{i1} WYfw_{il} + reg\_n_2 WYfc3_{il} \\ &+ \delta_f \lambda_{fv} VYfc2_{il} + \lambda_{fn} WXfc2_{il} \\ &- [d_i + \mu_i + (1 + HR_2)tr_{i3} + reg\_v_1 + reg\_n_1]WYfc2_{il} \\ &+ d_{i-1}WYfc2_{(i-1)l}\end{aligned} \tag{167}$$

$$
\begin{aligned}
\frac{dWYfc3_{1l}}{dt} &= (1+HR_1)tr_{12}WYfw_{1l} + (1+HR_2)tr_{13}WYfc2_{1l} \\
&+ \delta_f \lambda_{fv} VYfc3_{1l} + \lambda_{fn} WXfc3_{1l} \\
&- [d_1 + \mu_1 + (1+HR_3)tr_{14} + reg\_v_2 + reg\_n_2 + h_1]WYfc3_{1l}
\end{aligned} \tag{168}
$$

$$
\begin{aligned}
\frac{dWYfc3_{il}}{dt} &= \upsilon \phi_{fil} YYfc3_{il} \\
&+ (1+HR_1)tr_{i2}WYfw_{il} + (1+HR_2)tr_{i3}WYfc2_{il} \\
&+ \delta_f \lambda_{fv} VYfc3_{il} + \lambda_{fn} WXfc3_{il} \\
&- [d_i + \mu_i + (1+HR_3)tr_{i4} + reg\_v_2 + reg\_n_2 + h_i]WYfc3_{il} \\
&+ d_{i-1}WYfc3_{(i-1)l}
\end{aligned} \tag{169}
$$

$$
\begin{aligned}
\frac{dWYfpcl_{1l}}{dt} &= (1+HR_3)tr_{14}WYfc3_{1l} \\
&- (d_1 + \mu_1 + canc\_prog_1 + detect_1)WYfpcl_{1l}
\end{aligned} \tag{170}
$$

$$
\begin{aligned}
\frac{dWYfpcl_{il}}{dt} &= (1+HR_3)tr_{i4}WYfc3_{il} \\
&- (d_i + \mu_i + canc\_prog_1 + detect_1)WYfpcl_{il} \\
&+ d_{i-1}WYfpcl_{(i-1)l}
\end{aligned} \tag{171}
$$

$$
\begin{aligned}
\frac{dWYfcl_{1l}}{dt} &= detect_1 WYpcl_{1l} \\
&- (d_1 + \mu_1 + canc\_surv_1 + canc\_death_1)WYfcl_{1l}
\end{aligned} \tag{172}
$$

$$
\begin{aligned}
\frac{dWYfcl_{il}}{dt} &= detect_1 WYpcl_{il} \\
&- (d_i + \mu_i + canc\_surv_1 + canc\_death_1)WYfcl_{il} \\
&+ d_{i-1}WYfcl_{(i-1)l}
\end{aligned} \tag{173}
$$

$$
\begin{aligned}
\frac{dWYfpcr_{1l}}{dt} &= canc\_prog_1 WYfpcl_{1l} \\
&- (d_1 + \mu_1 + canc\_prog_2 + detect_2)WYfpcr_{1l}
\end{aligned} \tag{174}
$$

$$
\begin{aligned}
\frac{dWYfpcr_{il}}{dt} &= canc\_prog_1 WYfpcl_{il} \\
&- (d_i + \mu_i + canc\_prog_2 + detect_2)WYfpcr_{il} \\
&+ d_{i-1}WYfpcr_{(i-1)l}
\end{aligned} \tag{175}
$$

$$\frac{dWYfcr_{1l}}{dt} = detect_2 WYpcr_{1l} - (d_1 + \mu_1 + canc\_surv_2 + canc\_death_2) WYfcr_{1l} \tag{176}$$

$$\frac{dWYfcr_{il}}{dt} = detect_2 WYpcr_{il} - (d_i + \mu_i + canc\_surv_2 + canc\_death_2) WYfcr_{il} + d_{i-1} WYfcr_{(i-1)l} \tag{177}$$

$$\frac{dWYfpcd_{1l}}{dt} = canc\_prog_2 WYfpcr_{1l} - (d_1 + \mu_1 + detect_3) WYfpcd_{1l} \tag{178}$$

$$\frac{dWYfpcd_{il}}{dt} = canc\_prog_2 WYfpcr_{il} - (d_i + \mu_i + detect_3) WYfpcd_{il} + d_{i-1} WYfpcd_{(i-1)l} \tag{179}$$

$$\frac{dWYfcd_{1l}}{dt} = detect_3 WYpcd_{1l} - (d_1 + \mu_1 + canc\_surv_3 + canc\_death_3) WYfcd_{1l} \tag{180}$$

$$\frac{dWYfcd_{il}}{dt} = detect_3 WYpcd_{il} - (d_i + \mu_i + canc\_surv_3 + canc\_death_3) WYfcd_{il} + d_{i-1} WYfcd_{(i-1)l} \tag{181}$$

$$\frac{dWZmw_{1l}}{dt} = \delta_m \lambda_{mv} VZmw_{1l} + \nu_m \gamma_{mn} WYmw_{1l} - (d_1 + \mu_1 + \alpha\gamma_{mv} + \tau_{mn}) WZmw_{1l} \tag{182}$$

$$\frac{dWZmw_{il}}{dt} = \delta_m \lambda_{mv} VZmw_{il} + \upsilon\phi_{mil} YZmw_{il} + \nu_m \gamma_{mn} WYmw_{il} - (d_i + \mu_i + \alpha\gamma_{mv} + \tau_{mn}) WZmw_{il} + d_{i-1} WZmw_{(i-1)l} \tag{183}$$

$$\begin{aligned}\frac{dWZfw_{1l}}{dt} &= \delta_f\lambda_{fv}VZfw_{1l}\\ &+ \nu_f(\gamma_{fn}WYfw_{1l} + reg\_n_1WYfc2_{1l})\\ &- [d_1 + \mu_1 + h_1 + \alpha\gamma_{fv} + HR_1(tr_{11} + tr_{12}) + \tau_{fn}]WZfw_{1l}\end{aligned} \tag{184}$$

$$\begin{aligned}\frac{dWZfw_{il}}{dt} &= \delta_f\lambda_{fv}VZfw_{il} + \upsilon\phi_{fil}YZfw_{il}\\ &+ \nu_f(\gamma_{fn}WYfw_{il} + reg\_n_1WYfc2_{il})\\ &- [d_i + \mu_i + h_i + \alpha\gamma_{fv} + HR_1(tr_{i1} + tr_{i2}) + \tau_{fn}]WZfw_{il}\\ &+ d_{i-1}WZfw_{(i-1)l}\end{aligned} \tag{185}$$

$$\begin{aligned}\frac{dWZfc2_{1l}}{dt} &= HR_1tr_{11}WZfw_{1l} + reg\_v_2WZfc3_{1l}\\ &- (d_1 + \mu_1 + HR_2tr_{13} + reg\_v_1 + h_1 + \tau_{fn})WZfc2_{1l}\end{aligned} \tag{186}$$

$$\begin{aligned}\frac{dWZfc2_{il}}{dt} &= HR_1tr_{i1}WZfw_{il} + \upsilon\phi_{fil}YZfc2_{il} + reg\_v_2WZfc3_{il}\\ &- (d_i + \mu_i + HR_2tr_{i3} + reg\_v_1 + h_i + \tau_{fn})WZfc2_{il}\\ &+ d_{i-1}WZfc2_{(i-1)l}\end{aligned} \tag{187}$$

$$\begin{aligned}\frac{dWZfc3_{1l}}{dt} &= HR_1tr_{12}WZfw_{1l} + HR_2tr_{13}WZfc2_{1l}\\ &- (d_1 + \mu_1 + h_1 + HR_3tr_{14} + reg\_v_2 + \tau_{fn})WZfc3_{1l}\end{aligned} \tag{188}$$

$$\begin{aligned}\frac{dWZfc3_{il}}{dt} &= HR_1tr_{i2}WZfw_{il} + HR_2tr_{i3}WZfc2_{il} + \upsilon\phi_{fil}YZfc3_{il}\\ &- (d_i + \mu_i + h_i + HR_3tr_{i4} + reg\_v_2 + \tau_{fn})WZfc3_{il}\\ &+ d_{i-1}WZfc3_{(i-1)l}\end{aligned} \tag{189}$$

$$\begin{aligned}\frac{dWZfpcl_{1l}}{dt} &= HR_3tr_{14}WZfc3_{1l}\\ &- (d_1 + \mu_1 + canc\_prog_1 + detect_1 + \tau_{fn})WZfpcl_{1l}\end{aligned} \tag{190}$$

$$\begin{aligned}\frac{dWZfpcl_{il}}{dt} &= HR_3tr_{i4}WZfc3_{il}\\ &- (d_i + \mu_i + canc\_prog_1 + detect_1 + \tau_{fn})WZfpcl_{il}\\ &+ d_{i-1}WZfpcl_{(i-1)l}\end{aligned} \tag{191}$$

$$\begin{aligned}\frac{dWZfcl_{1l}}{dt} &= detect_1 WZpcl_{1l} \\ &- (d_1 + \mu_1 + canc\_surv_1 + canc\_death_1)WZfcl_{1l}\end{aligned} \tag{192}$$

$$\begin{aligned}\frac{dWZfcl_{il}}{dt} &= detect_1 WZpcl_{il} \\ &- (d_i + \mu_i + canc\_surv_1 + canc\_death_1)WZfcl_{il} \\ &+ d_{i-1} WZfcl_{(i-1)l}\end{aligned} \tag{193}$$

$$\begin{aligned}\frac{dWZfpcr_{1l}}{dt} &= canc\_prog_1 WZfpcl_{1l} \\ &- (d_1 + \mu_1 + canc\_prog_2 + detect_2 + \tau_{fn})WZfpcr_{1l}\end{aligned} \tag{194}$$

$$\begin{aligned}\frac{dWZfpcr_{il}}{dt} &= canc\_prog_1 WZfpcl_{il} \\ &- (d_i + \mu_i + canc\_prog_2 + detect_2 + \tau_{fn})WZfpcr_{il} \\ &+ d_{i-1} WZfpcr_{(i-1)l}\end{aligned} \tag{195}$$

$$\begin{aligned}\frac{dWZfcr_{1l}}{dt} &= detect_2 WZpcr_{1l} \\ &- (d_1 + \mu_1 + canc\_surv_2 + canc\_death_2)WZfcr_{1l}\end{aligned} \tag{196}$$

$$\begin{aligned}\frac{dWZfcr_{il}}{dt} &= detect_2 WZpcr_{il} \\ &- (d_i + \mu_i + canc\_surv_2 + canc\_death_2)WZfcr_{il} \\ &+ d_{i-1} WZfcr_{(i-1)l}\end{aligned} \tag{197}$$

$$\begin{aligned}\frac{dWZfpcd_{1l}}{dt} &= canc\_prog_2 WZfpcr_{1l} \\ &- (d_1 + \mu_1 + detect_3 + \tau_{fn})WZfpcd_{1l}\end{aligned} \tag{198}$$

$$\begin{aligned}\frac{dWZfpcd_{il}}{dt} &= canc\_prog_2 WZfpcr_{il} \\ &- (d_i + \mu_i + detect_3 + \tau_{fn})WZfpcd_{il} \\ &+ d_{i-1} WZfpcd_{(i-1)l}\end{aligned} \tag{199}$$

$$\begin{aligned}\frac{dWZfcd_{1l}}{dt} &= detect_3 WZpcd_{1l} \\ &- (d_1 + \mu_1 + canc\_surv_3 + canc\_death_3)WZfcd_{1l}\end{aligned} \tag{200}$$

$$
\begin{aligned}
\frac{dWZfcd_{il}}{dt} &= detect_3 WZpcd_{il} \\
&- (d_i + \mu_i + canc\_surv_3 + canc\_death_3) WZfcd_{il} \\
&+ d_{i-1} WZfcd_{(i-1)l}
\end{aligned}
\tag{201}
$$

$$
\begin{aligned}
\frac{dZXmw_{1l}}{dt} &= \nu_m \gamma_{mv} (YXmw_{1l} + \alpha WXmw_{1l}) \\
&+ (1 - \nu_m) \gamma_{mn} ZYmw_{1l} \\
&+ \tau_{mn} ZZmw_{1l} \\
&- (d_1 + \mu_1 + \lambda_{mn} + \tau_{mv}) ZXmw_{1l}
\end{aligned}
\tag{202}
$$

$$
\begin{aligned}
\frac{dZXmw_{il}}{dt} &= \nu_m \gamma_{mv} (YXmw_{il} + \alpha WXmw_{il}) \\
&+ (1 - \nu_m) \gamma_{mn} ZYmw_{il} \\
&+ \tau_{mn} ZZmw_{il} \\
&- (d_i + \mu_i + \lambda_{mn} + \tau_{mv}) ZXmw_{il} \\
&+ d_{i-1} ZXmw_{(i-1)l}
\end{aligned}
\tag{203}
$$

$$
\begin{aligned}
\frac{dZXfw_{1l}}{dt} &= \nu_f [\gamma_{fv} (YXfw_{1l} + \alpha WXfw_{1l}) \\
&+ reg\_v_1 (YXfc2_{1l} + WXfc2_{1l}) \\
&+ (1 - \nu_f)(\gamma_{fn} ZYfw_{1l} + reg\_n_1 ZYfc2_{1l}) \\
&+ \tau_{fn} ZZfw_{1l} \\
&- (d_1 + \mu_1 + h_1 + \lambda_{fn} + \tau_{fv}) ZXfw_{1l}
\end{aligned}
\tag{204}
$$

$$
\begin{aligned}
\frac{dZXfw_{il}}{dt} &= \nu_f [\gamma_{fv} (YXfw_{il} + \alpha WXfw_{il}) \\
&+ reg\_v_1 (YXfc2_{il} + WXfc2_{il}) \\
&+ (1 - \nu_f)(\gamma_{fn} ZYfw_{il} + reg\_n_1 ZYfc2_{il}) \\
&+ \tau_{fn} ZZfw_{il} \\
&- (d_i + \mu_i + h_i + \lambda_{fn} + \tau_{fv}) ZXfw_{il} \\
&+ d_{i-1} ZXfw_{(i-1)l}
\end{aligned}
\tag{205}
$$

$$
\begin{aligned}
\frac{dZYmw_{1l}}{dt} &= \nu_m \gamma_{mv} (YYmw_{1l} + \alpha WYmw_{1l}) \\
&+ \lambda_{mn} ZXmw_{1l} \\
&- (d_1 + \mu_1 + \gamma_{mn} + \tau_{mv}) ZYmw_{1l}
\end{aligned}
\tag{206}
$$

$$\begin{aligned}\frac{dZYmw_{il}}{dt} &= \nu_m\gamma_{mv}(YYmw_{il} + \alpha WYmw_{il}) \\ &+ \lambda_{mn}ZXmw_{il} \\ &- (d_i + \mu_i + \gamma_{mn} + \tau_{mv})ZYmw_{il} \\ &+ d_{i-1}ZYmw_{(i-1)l}\end{aligned} \tag{207}$$

$$\begin{aligned}\frac{dZYfw_{1l}}{dt} &= \nu_f[\gamma_{fv}(YYfw_{1l} + \alpha WYfw_{1l}) \\ &+ reg\_v_1(YYfc2_{1l} + WYfc2_{1l})] \\ &+ \lambda_{fn}ZXfw_{1l} \\ &- (d_1 + \mu_1 + h_1 + \gamma_{fn} + \tau_{fv} + tr_{11} + tr_{12})ZYfw_{1l}\end{aligned} \tag{208}$$

$$\begin{aligned}\frac{dZYfw_{il}}{dt} &= \nu_f[\gamma_{fv}(YYfw_{il} + \alpha WYfw_{il}) \\ &+ reg\_v_1(YYfc2_{il} + WYfc2_{il})] \\ &+ \lambda_{fn}ZXfw_{il} \\ &- (d_i + \mu_i + h_i + \gamma_{fn} + \tau_{fv} + tr_{i1} + tr_{i2})ZYfw_{il} \\ &+ d_{i-1}ZYfw_{(i-1)l}\end{aligned} \tag{209}$$

$$\begin{aligned}\frac{dZYfc2_{1l}}{dt} &= tr_{11}ZYfw_{1l} + reg\_n_2ZYfc3_{1l} \\ &- (d_1 + \mu_1 + tr_{13} + reg\_n_1 + \tau_{fv} + h_1)ZYfc2_{1l}\end{aligned} \tag{210}$$

$$\begin{aligned}\frac{dZYfc2_{il}}{dt} &= tr_{i1}ZYfw_{il} + reg\_n_2ZYfc3_{il} \\ &- (d_i + \mu_i + tr_{i3} + reg\_n_1 + \tau_{fv} + h_i)ZYfc2_{il} \\ &+ d_{i-1}ZYfc2_{(i-1)l}\end{aligned} \tag{211}$$

$$\begin{aligned}\frac{dZYfc3_{1l}}{dt} &= tr_{12}ZYfw_{1l} + tr_{13}ZYfc2_{1l} \\ &- (d_1 + \mu_i + tr_{14} + reg\_n_2 + \tau_{fv} + h_1)ZYfc3_{1l}\end{aligned} \tag{212}$$

$$\begin{aligned}\frac{dZYfc3_{il}}{dt} &= tr_{i2}ZYfw_{il} + tr_{i3}ZYfc2_{il} \\ &- (d_i + \mu_i + tr_{i4} + reg\_n_2 + \tau_{fv} + h_i)ZYfc3_{il} \\ &+ d_{i-1}ZYfc3_{(i-1)l}\end{aligned} \tag{213}$$

$$\begin{aligned}\frac{dZYfpcl_{1l}}{dt} &= tr_{14}ZYfc3_{1l} \\ &- (d_1 + \mu_1 + canc\_prog_1 + detect_1 + \tau_{fv})ZYfpcl_{1l}\end{aligned} \tag{214}$$

$$\begin{aligned}\frac{dZYfpcl_{il}}{dt} &= tr_{i4}ZYfc3_{il} \\ &- (d_i + \mu_i + canc\_prog_1 + detect_1 + \tau_{fv})ZYfpcl_{il} \\ &+ d_{i-1}ZYfpcl_{(i-1)l}\end{aligned} \tag{215}$$

$$\begin{aligned}\frac{dZYfcl_{1l}}{dt} &= detect_1 ZYpcl_{1l} \\ &- (d_1 + \mu_1 + canc\_surv_1 + canc\_death_1)ZYfcl_{1l}\end{aligned} \tag{216}$$

$$\begin{aligned}\frac{dZYfcl_{il}}{dt} &= detect_1 ZYpcl_{il} \\ &- (d_i + \mu_i + canc\_surv_1 + canc\_death_1)ZYfcl_{il} \\ &+ d_{i-1}ZYfcl_{(i-1)l}\end{aligned} \tag{217}$$

$$\begin{aligned}\frac{dZYfpcr_{1l}}{dt} &= canc\_prog_1 ZYfpcl_{1l} \\ &- (d_1 + \mu_1 + canc\_prog_2 + detect_2 + \tau_{fv})ZYfpcr_{1l}\end{aligned} \tag{218}$$

$$\begin{aligned}\frac{dZYfpcr_{il}}{dt} &= canc\_prog_1 ZYfpcl_{il} \\ &- (d_i + \mu_i + canc\_prog_2 + detect_2 + \tau_{fv})ZYfpcr_{il} \\ &+ d_{i-1}ZYfpcr_{(i-1)l}\end{aligned} \tag{219}$$

$$\begin{aligned}\frac{dZYfcr_{1l}}{dt} &= detect_2 ZYpcr_{1l} \\ &- (d_1 + \mu_1 + canc\_surv_2 + canc\_death_2)ZYfcr_{1l}\end{aligned} \tag{220}$$

$$\begin{aligned}\frac{dZYfcr_{il}}{dt} &= detect_2 ZYpcr_{il} \\ &- (d_i + \mu_i + canc\_surv_2 + canc\_death_2)ZYfcr_{il} \\ &+ d_{i-1}ZYfcr_{(i-1)l}\end{aligned} \tag{221}$$

$$\begin{aligned}\frac{dZYfpcd_{1l}}{dt} &= canc\_prog_2 ZYfpcr_{1l} \\ &- (d_1 + \mu_1 + detect_3 + \tau_{fv})ZYfpcd_{1l}\end{aligned} \tag{222}$$

$$\begin{aligned}\frac{dZYfpcd_{il}}{dt} &= canc\_prog_2 ZYfpcr_{il} \\ &- (d_i + \mu_i + detect_3 + \tau_{fv})ZYfpcd_{il} \\ &+ d_{i-1}ZYfpcd_{(i-1)l}\end{aligned} \tag{223}$$

$$\begin{aligned}\frac{dZYfcd_{1l}}{dt} &= detect_3 ZYpcd_{1l} \\ &- (d_1 + \mu_1 + canc\_surv_3 + canc\_death_3)ZYfcd_{1l}\end{aligned} \tag{224}$$

$$\begin{aligned}\frac{dZYfcd_{il}}{dt} &= detect_3 ZYpcd_{il} \\ &- (d_i + \mu_i + canc\_surv_3 + canc\_death_3)ZYfcd_{il} \\ &+ d_{i-1}ZYfcd_{(i-1)l}\end{aligned} \tag{225}$$

$$\begin{aligned}\frac{dZZmw_{1l}}{dt} &= \nu_m[\gamma_{mv}(YZmw_{1l} + \alpha WZmw_{1l}) + \gamma_{mn}ZYmw_{1l}] \\ &- (d_1 + \mu_1 + \tau_{mv} + \tau_{mn})ZZmw_{1l}\end{aligned} \tag{226}$$

$$\begin{aligned}\frac{dZZmw_{il}}{dt} &= \nu_m[\gamma_{mv}(YZmw_{il} + \alpha WZmw_{il}) + \gamma_{mn}ZYmw_{il}] \\ &- (d_i + \mu_i + \tau_{mv} + \tau_{mn})ZZmw_{il} \\ &+ d_{i-1}ZZmw_{(i-1)l}\end{aligned} \tag{227}$$

$$\begin{aligned}\frac{dZZfw_{1l}}{dt} &= \nu_f[\gamma_{fv}(YZfw_{1l} + \alpha WZfw_{1l}) + \gamma_{fn}ZYfw_{1l} \\ &+ reg\_v_1(YZfc2_{1l} + WZfc2_{1l}) \\ &+ reg\_n_1 ZYfc2_{1l}] \\ &- (d_1 + \mu_1 + \tau_{fv} + \tau_{fn} + h_1)ZZfw_{1l}\end{aligned} \tag{228}$$

$$\begin{aligned}\frac{dZZfw_{il}}{dt} &= \nu_f[\gamma_{fv}(YZfw_{il} + \alpha WZfw_{il}) + \gamma_{fn}ZYfw_{il} \\ &+ reg\_v_1(YZfc2_{il} + WZfc2_{il}) \\ &+ reg\_n_1 ZYfc2_{il}] \\ &- (d_i + \mu_i + \tau_{fv} + \tau_{fn} + h_i)ZZfw_{il} \\ &+ d_{i-1}ZZfw_{(i-1)l}\end{aligned} \tag{229}$$

$$
\begin{aligned}
\frac{dH_{1l}}{dt} &= h_1(XXfw_{1l} + XYfw_{1l} + XYfc2_{1l} + XYfc3_{1l} + XZfw_{1l} \\
&+ YXfw_{1l} + YXfc2_{1l} + XYfc3_{1l} + YYfw_{1l} + YYfc2_{1l} \\
&+ YYfc3_{1l} + YZfw_{1l} + YZfc2_{1l} + YZfc3_{1l} + VXfw_{1l} \\
&+ VYfw_{1l} + VYfc2_{1l} + VYfc3_{1l} + VZfw_{1l} + WXfw_{1l} \\
&+ WXfc2_{1l} + WXfc3_{1l} + WYfw_{1l} + WYfc2_{1l} + WYfc3_{1l} \\
&+ WZfw_{1l} + WZfc2_{1l} + WZfc3_{1l} + ZXfw_{1l} + ZYfw_{1l} \\
&+ ZYfc2_{1l} + ZYfc3_{1l} + ZZfw_{1l}) \\
&- (d_1 + \mu_1)H_{1l}
\end{aligned} \tag{230}
$$

$$
\begin{aligned}
\frac{dH_{il}}{dt} &= h_i(XXfw_{il} + XYfw_{il} + XYfc2_{il} + XYfc3_{il} + XZfw_{il} \\
&+ YXfw_{il} + YXfc2_{il} + XYfc3_{il} + YYfw_{il} + YYfc2_{il} \\
&+ YYfc3_{il} + YZfw_{il} + YZfc2_{il} + YZfc3_{il} + VXfw_{il} \\
&+ VYfw_{il} + VYfc2_{il} + VYfc3_{il} + VZfw_{il} + WXfw_{il} \\
&+ WXfc2_{il} + WXfc3_{il} + WYfw_{il} + WYfc2_{il} + WYfc3_{il} \\
&+ WZfw_{il} + WZfc2_{il} + WZfc3_{il} + ZXfw_{il} + ZYfw_{il} \\
&+ ZYfc2_{il} + ZYfc3_{il} + ZZfw_{il}) \\
&- (d_i + \mu_i)H_{il} \\
&+ d_{i-1}H_{(i-1)l}
\end{aligned} \tag{231}
$$

$$
\begin{aligned}
\frac{dCDL2yr_{1l}}{dt} &= canc\_surv_1(XYfcl_{1l} + YXfcl_{1l} + YYfcl_{1l} \\
&+ YZfcl_{1l} + ZYfcl_{1l} + VYfcl_{1l} \\
&+ WXfcl_{1l} + WYfcl_{1l} + WZfcl_{1l}) \\
&- (d_1 + \mu_1 + canc\_surv_4 + canc\_death_4)CDL2yr_{1l}
\end{aligned} \tag{232}
$$

$$
\begin{aligned}
\frac{dCDL2yr_{il}}{dt} &= canc\_surv_1(XYfcl_{il} + YXfcl_{il} + YYfcl_{il} \\
&+ YZfcl_{il} + ZYfcl_{il} + VYfcl_{il} \\
&+ WXfcl_{il} + WYfcl_{il} + WZfcl_{il}) \\
&- (d_i + \mu_i + canc\_surv_4 + canc\_death_4)CDL2yr_{il} \\
&+ d_{i-1}CDL2yr_{(i-1)l}
\end{aligned} \tag{233}
$$

$$
\begin{aligned}
\frac{dCDL3yr_{1l}}{dt} &= canc\_surv_4 CDL2yr_{1l} \\
&- (d_1 + \mu_1 + canc\_surv_7 + canc\_death_7)CDL3yr_{1l}
\end{aligned} \tag{234}
$$

$$\begin{aligned}\frac{dCDL3yr_{il}}{dt} &= canc\_surv_4 CDL2yr_{il} \\ &- (d_i + \mu_i + canc\_surv_7 + canc\_death_7) CDL3yr_{il} \\ &+ d_{i-1} CDL3yr_{(i-1)l}\end{aligned} \tag{235}$$

$$\begin{aligned}\frac{dCDL4yr_{1l}}{dt} &= canc\_surv_7 CDL3yr_{1l} \\ &- (d_1 + \mu_1 + canc\_surv_{10} + canc\_death_{10}) CDL4yr_{1l}\end{aligned} \tag{236}$$

$$\begin{aligned}\frac{dCDL4yr_{il}}{dt} &= canc\_surv_7 CDL3yr_{il} \\ &- (d_i + \mu_i + canc\_surv_{10} + canc\_death_{10}) CDL4yr_{il} \\ &+ d_{i-1} CDL4yr_{(i-1)l}\end{aligned} \tag{237}$$

$$\begin{aligned}\frac{dCDL5yr_{1l}}{dt} &= canc\_surv_{10} CDL4yr_{1l} \\ &- (d_1 + \mu_1 + canc\_surv_{13} + canc\_death_{13}) CDL5yr_{1l}\end{aligned} \tag{238}$$

$$\begin{aligned}\frac{dCDL5yr_{il}}{dt} &= canc\_surv_{10} CDL4yr_{il} \\ &- (d_i + \mu_i + canc\_surv_{13} + canc\_death_{13}) CDL5yr_{il} \\ &+ d_{i-1} CDL5yr_{(i-1)l}\end{aligned} \tag{239}$$

$$\begin{aligned}\frac{dCDL6yr_{1l}}{dt} &= canc\_surv_{13} CDL5yr_{1l} \\ &- (d_1 + \mu_1 + canc\_surv_{16} + canc\_death_{16}) CDL6yr_{1l}\end{aligned} \tag{240}$$

$$\begin{aligned}\frac{dCDL6yr_{il}}{dt} &= canc\_surv_{13} CDL5yr_{il} \\ &- (d_i + \mu_i + canc\_surv_{16} + canc\_death_{16}) CDL6yr_{il} \\ &+ d_{i-1} CDL6yr_{(i-1)l}\end{aligned} \tag{241}$$

$$\begin{aligned}\frac{dCDL7yr_{1l}}{dt} &= canc\_surv_{16} CDL6yr_{1l} \\ &- (d_1 + \mu_1 + canc\_surv_{19} + canc\_death_{19}) CDL7yr_{1l}\end{aligned} \tag{242}$$

$$\begin{aligned}\frac{dCDL7yr_{il}}{dt} &= canc\_surv_{16}CDL6yr_{il} \\ &- (d_i + \mu_i + canc\_surv_{19} + canc\_death_{19})CDL7yr_{il} \\ &+ d_{i-1}CDL7yr_{(i-1)l}\end{aligned} \tag{243}$$

$$\begin{aligned}\frac{dCDL8yr_{1l}}{dt} &= canc\_surv_{19}CDL7yr_{1l} \\ &- (d_1 + \mu_1 + canc\_surv_{22} + canc\_death_{22})CDL8yr_{1l}\end{aligned} \tag{244}$$

$$\begin{aligned}\frac{dCDL8yr_{il}}{dt} &= canc\_surv_{19}CDL7yr_{il} \\ &- (d_i + \mu_i + canc\_surv_{22} + canc\_death_{22})CDL8yr_{il} \\ &+ d_{i-1}CDL8yr_{(i-1)l}\end{aligned} \tag{245}$$

$$\begin{aligned}\frac{dCDL9yr_{1l}}{dt} &= canc\_surv_{22}CDL8yr_{1l} \\ &- (d_1 + \mu_1 + canc\_surv_{25} + canc\_death_{25})CDL9yr_{1l}\end{aligned} \tag{246}$$

$$\begin{aligned}\frac{dCDL9yr_{il}}{dt} &= canc\_surv_{22}CDL8yr_{il} \\ &- (d_i + \mu_i + canc\_surv_{25} + canc\_death_{25})CDL9yr_{il} \\ &+ d_{i-1}CDL9yr_{(i-1)l}\end{aligned} \tag{247}$$

$$\begin{aligned}\frac{dCDL10yr_{1l}}{dt} &= canc\_surv_{25}CDL9yr_{1l} \\ &- (d_1 + \mu_1 + canc\_surv_{28} + canc\_death_{28})CDL10yr_{1l}\end{aligned} \tag{248}$$

$$\begin{aligned}\frac{dCDL10yr_{il}}{dt} &= canc\_surv_{25}CDL9yr_{il} \\ &- (d_i + \mu_i + canc\_surv_{28} + canc\_death_{28})CDL10yr_{il} \\ &+ d_{i-1}CDL10yr_{(i-1)l}\end{aligned} \tag{249}$$

$$\begin{aligned}\frac{dCDR2yr_{1l}}{dt} &= canc\_surv_2(XYfcr_{1l} + YXfcr_{1l} + YYfcr_{1l} \\ &+ YZfcr_{1l} + VYfcr_{1l} + WXfcr_{1l} \\ &+ WYfcr_{1l} + WZfcr_{1l} + ZYfcr_{1l}) \\ &- (d_1 + \mu_1 + canc\_surv_5 + canc\_death_5)CDR2yr_{1l}\end{aligned} \tag{250}$$

$$
\begin{aligned}
\frac{dCDR2yr_{il}}{dt} &= canc\_surv_2(XYfcr_{il} + YXfcr_{il} + YYfcr_{il} \\
&+ YZfcr_{il} + VYfcr_{il} + WXfcr_{il} \\
&+ WYfcr_{il} + WZfcr_{il} + ZYfcr_{il}) \\
&- (d_i + \mu_i + canc\_surv_5 + canc\_death_5)CDR2yr_{il} \\
&+ d_{i-1}CDR2yr_{(i-1)l}
\end{aligned} \tag{251}
$$

$$
\begin{aligned}
\frac{dCDR3yr_{1l}}{dt} &= canc\_surv_5 CDR2yr_{1l} \\
&- (d_1 + \mu_1 + canc\_surv_8 + canc\_death_8)CDR3yr_{1l}
\end{aligned} \tag{252}
$$

$$
\begin{aligned}
\frac{dCDR3yr_{il}}{dt} &= canc\_surv_5 CDR2yr_{il} \\
&- (d_i + \mu_i + canc\_surv_8 + canc\_death_8)CDR3yr_{il} \\
&+ d_{i-1}CDR3yr_{(i-1)l}
\end{aligned} \tag{253}
$$

$$
\begin{aligned}
\frac{dCDR4yr_{1l}}{dt} &= canc\_surv_8 CDR3yr_{1l} \\
&- (d_1 + \mu_1 + canc\_surv_{11} + canc\_death_{11})CDR4yr_{1l}
\end{aligned} \tag{254}
$$

$$
\begin{aligned}
\frac{dCDR4yr_{il}}{dt} &= canc\_surv_8 CDR3yr_{il} \\
&- (d_i + \mu_i + canc\_surv_{11} + canc\_death_{11})CDR4yr_{il} \\
&+ d_{i-1}CDR4yr_{(i-1)l}
\end{aligned} \tag{255}
$$

$$
\begin{aligned}
\frac{dCDR5yr_{1l}}{dt} &= canc\_surv_{11} CDR4yr_{1l} \\
&- (d_1 + \mu_1 + canc\_surv_{14} + canc\_death_{14})CDR5yr_{1l}
\end{aligned} \tag{256}
$$

$$
\begin{aligned}
\frac{dCDR5yr_{il}}{dt} &= canc\_surv_{11} CDR4yr_{il} \\
&- (d_i + \mu_i + canc\_surv_{14} + canc\_death_{14})CDR5yr_{il} \\
&+ d_{i-1}CDR5yr_{(i-1)l}
\end{aligned} \tag{257}
$$

$$
\begin{aligned}
\frac{dCDR6yr_{1l}}{dt} &= canc\_surv_{14} CDR5yr_{1l} \\
&- (d_1 + \mu_1 + canc\_surv_{17} + canc\_death_{17})CDR6yr_{1l}
\end{aligned} \tag{258}
$$

$$\begin{aligned}\frac{dCDR6yr_{il}}{dt} &= canc\_surv_{14}CDR5yr_{il} \\ &- (d_i + \mu_i + canc\_surv_{17} + canc\_death_{17})CDR6yr_{il} \\ &+ d_{i-1}CDR6yr_{(i-1)l}\end{aligned} \tag{259}$$

$$\begin{aligned}\frac{dCDR7yr_{1l}}{dt} &= canc\_surv_{17}CDR6yr_{1l} \\ &- (d_1 + \mu_1 + canc\_surv_{20} + canc\_death_{20})CDR7yr_{1l}\end{aligned} \tag{260}$$

$$\begin{aligned}\frac{dCDR7yr_{il}}{dt} &= canc\_surv_{17}CDR6yr_{il} \\ &- (d_i + \mu_i + canc\_surv_{20} + canc\_death_{20})CDR7yr_{il} \\ &+ d_{i-1}CDR7yr_{(i-1)l}\end{aligned} \tag{261}$$

$$\begin{aligned}\frac{dCDR8yr_{1l}}{dt} &= canc\_surv_{20}CDR7yr_{1l} \\ &- (d_1 + \mu_1 + canc\_surv_{23} + canc\_death_{23})CDR8yr_{1l}\end{aligned} \tag{262}$$

$$\begin{aligned}\frac{dCDR8yr_{il}}{dt} &= canc\_surv_{20}CDR7yr_{il} \\ &- (d_i + \mu_i + canc\_surv_{23} + canc\_death_{23})CDR8yr_{il} \\ &+ d_{i-1}CDR8yr_{(i-1)l}\end{aligned} \tag{263}$$

$$\begin{aligned}\frac{dCDR9yr_{1l}}{dt} &= canc\_surv_{23}CDR8yr_{1l} \\ &- (d_1 + \mu_1 + canc\_surv_{26} + canc\_death_{26})CDR9yr_{1l}\end{aligned} \tag{264}$$

$$\begin{aligned}\frac{dCDR9yr_{il}}{dt} &= canc\_surv_{23}CDR8yr_{il} \\ &- (d_i + \mu_i + canc\_surv_{26} + canc\_death_{26})CDR9yr_{il} \\ &+ d_{i-1}CDR9yr_{(i-1)l}\end{aligned} \tag{265}$$

$$\begin{aligned}\frac{dCDR10yr_{1l}}{dt} &= canc\_surv_{26}CDR9yr_{1l} \\ &- (d_1 + \mu_1 + canc\_surv_{29} + canc\_death_{29})CDR10yr_{1l}\end{aligned} \tag{266}$$

$$\begin{aligned}\frac{dCDR10yr_{il}}{dt} &= canc\_surv_{26}CDR9yr_{il} \\ &- (d_i + \mu_i + canc\_surv_{29} + canc\_death_{29})CDR10yr_{il} \\ &+ d_{i-1}CDR10yr_{(i-1)l}\end{aligned} \tag{267}$$

$$\begin{aligned}\frac{dCDD2yr_{1l}}{dt} &= canc\_surv_3(XYfcd_{1l} + YXfcd_{1l} + YYfcd_{1l} \\ &+ YZfcd_{1l} + VYfcd_{1l} + WXfcd_{1l} \\ &+ WYfcd_{1l} + WZfcd_{1l} + ZYfcd_{1l}) \\ &- (d_1 + \mu_1 + canc\_surv_6 + canc\_death_6)CDD2yr_{1l}\end{aligned} \tag{268}$$

$$\begin{aligned}\frac{dCDD2yr_{il}}{dt} &= canc\_surv_3(XYfcd_{il} + YXfcd_{il} + YYfcd_{il} \\ &+ YZfcd_{il} + VYfcd_{il} + WXfcd_{il} \\ &+ WYfcd_{il} + WZfcd_{il} + ZYfcd_{il}) \\ &- (d_i + \mu_i + canc\_surv_6 + canc\_death_6)CDD2yr_{il} \\ &+ d_{i-1}CDD2yr_{(i-1)l}\end{aligned} \tag{269}$$

$$\begin{aligned}\frac{dCDD3yr_{1l}}{dt} &= canc\_surv_6CDD2yr_{1l} \\ &- (d_1 + \mu_1 + canc\_surv_9 + canc\_death_9)CDD3yr_{1l}\end{aligned} \tag{270}$$

$$\begin{aligned}\frac{dCDD3yr_{il}}{dt} &= canc\_surv_6CDD2yr_{il} \\ &- (d_i + \mu_i + canc\_surv_9 + canc\_death_9)CDD3yr_{il} \\ &+ d_{i-1}CDD3yr_{(i-1)l}\end{aligned} \tag{271}$$

$$\begin{aligned}\frac{dCDD4yr_{1l}}{dt} &= canc\_surv_9CDD3yr_{1l} \\ &- (d_1 + \mu_1 + canc\_surv_{12} + canc\_death_{12})CDD4yr_{1l}\end{aligned} \tag{272}$$

$$\begin{aligned}\frac{dCDD4yr_{il}}{dt} &= canc\_surv_9CDD3yr_{il} \\ &- (d_i + \mu_i + canc\_surv_{12} + canc\_death_{12})CDD4yr_{il} \\ &+ d_{i-1}CDD4yr_{(i-1)l}\end{aligned} \tag{273}$$

$$
\begin{aligned}
\frac{dCDD5yr_{1l}}{dt} &= canc\_surv_{12} CDD4yr_{1l} \\
&- (d_1 + \mu_1 + canc\_surv_{15} + canc\_death_{15}) CDD5yr_{1l}
\end{aligned}
\tag{274}
$$

$$
\begin{aligned}
\frac{dCDD5yr_{il}}{dt} &= canc\_surv_{12} CDD4yr_{il} \\
&- (d_i + \mu_i + canc\_surv_{15} + canc\_death_{15}) CDD5yr_{il} \\
&+ d_{i-1} CDD5yr_{(i-1)l}
\end{aligned}
\tag{275}
$$

$$
\begin{aligned}
\frac{dCDD6yr_{1l}}{dt} &= canc\_surv_{15} CDD5yr_{1l} \\
&- (d_1 + \mu_1 + canc\_surv_{18} + canc\_death_{18}) CDD6yr_{1l}
\end{aligned}
\tag{276}
$$

$$
\begin{aligned}
\frac{dCDD6yr_{il}}{dt} &= canc\_surv_{15} CDD5yr_{il} \\
&- (d_i + \mu_i + canc\_surv_{18} + canc\_death_{18}) CDD6yr_{il} \\
&+ d_{i-1} CDD6yr_{(i-1)l}
\end{aligned}
\tag{277}
$$

$$
\begin{aligned}
\frac{dCDD7yr_{1l}}{dt} &= canc\_surv_{18} CDD6yr_{1l} \\
&- (d_1 + \mu_1 + canc\_surv_{21} + canc\_death_{21}) CDD7yr_{1l}
\end{aligned}
\tag{278}
$$

$$
\begin{aligned}
\frac{dCDD7yr_{il}}{dt} &= canc\_surv_{18} CDD6yr_{il} \\
&- (d_i + \mu_i + canc\_surv_{21} + canc\_death_{21}) CDD7yr_{il} \\
&+ d_{i-1} CDD7yr_{(i-1)l}
\end{aligned}
\tag{279}
$$

$$
\begin{aligned}
\frac{dCDD8yr_{1l}}{dt} &= canc\_surv_{21} CDD7yr_{1l} \\
&- (d_1 + \mu_1 + canc\_surv_{24} + canc\_death_{24}) CDD8yr_{1l}
\end{aligned}
\tag{280}
$$

$$
\begin{aligned}
\frac{dCDD8yr_{il}}{dt} &= canc\_surv_{21} CDD7yr_{il} \\
&- (d_i + \mu_i + canc\_surv_{24} + canc\_death_{23}) CDD8yr_{il} \\
&+ d_{i-1} CDD8yr_{(i-1)l}
\end{aligned}
\tag{281}
$$

$$
\begin{aligned}
\frac{dCDD9yr_{1l}}{dt} &= canc\_surv_{24} CDD8yr_{1l} \\
&- (d_1 + \mu_1 + canc\_surv_{27} + canc\_death_{27}) CDD9yr_{1l}
\end{aligned}
\tag{282}
$$

$$\frac{dCDD9yr_{il}}{dt} = canc\_surv_{24}CDD8yr_{il} - (d_i + \mu_i + canc\_surv_{27} + canc\_death_{27})CDD9yr_{il} + d_{i-1}CDD9yr_{(i-1)l} \tag{283}$$

$$\frac{dCDD10yr_{1l}}{dt} = canc\_surv_{27}CDD9yr_{1l} - (d_1 + \mu_1 + canc\_surv_{30} + canc\_death_{30})CDD10yr_{1l} \tag{284}$$

$$\frac{dCDD10yr_{il}}{dt} = canc\_surv_{27}CDD9yr_{il} - (d_i + \mu_i + canc\_surv_{30} + canc\_death_{30})CDD10yr_{il} + d_{i-1}CDD10yr_{(i-1)l} \tag{285}$$

$$\frac{dCSurv_{1l}}{dt} = canc\_surv_{28}CDL10yr_{1l} + canc\_surv_{29}CDR10yr_{1l} + canc\_surv_{30}CDD10yr_{1l} - (d_1 + \mu_1)CSurv_{1l} \tag{286}$$

$$\frac{dCSurv_{il}}{dt} = canc\_surv_{28}CDL10yr_{il} + canc\_surv_{29}CDR10yr_{il} + canc\_surv_{30}CDD10yr_{il} - (d_1 + \mu_1)CSurv_{il} + d_{i-1}CSurv_{(i-1)l} \tag{287}$$

[7] Center for Disease Control and Prevention. *National Survey of Family Growth (NSFG).* URL: https://www.cdc.gov/nchs/nsfg/index.htm.

[8] Center for Disease Control and Prevention. *Youth Risk Behavior Surveillance System (YRBSS).* URL: http://www.cdc.gov/healthyyouth/data/yrbs/index.htm.

[22] National Cancer Institute. Surveillance, Epidemiology, and End Results (SEER) Program. *Cancer Statistics.* Accessed at https://seer.cancer.gov/statistics/.

[29] University of California, Berkeley. *United States Mortality DataBase*. Available at usa.mortality.org (data downloaded on 02/10/22). 2023.

[30] Cosette M. Wheeler et al. "A population-based study of human papillomavirus genotype prevalence in the United States: baseline measures prior to mass human papillomavirus vaccination". In: *International journal of cancer* 132.1 (2013), pp. 198–207. DOI: 10.1002/ijc.27608.
